# Common and Rare Variant Contributions to Bradyarrhythmias from Multi-Ancestry Meta-Analyses

**DOI:** 10.1101/2023.09.24.23295485

**Authors:** Lu-Chen Weng, Joel T. Rämö, Sean J. Jurgens, Shaan Khurshid, Mark Chaffin, Amelia Weber Hall, Valerie N. Morrill, Victor Nauffal, Yan V. Sun, Dominik Beer, Simon Lee, Girish Nadkarni, ThuyVy Duong, Biqi Wang, Tomasz Czuba, Thomas R. Austin, Zachary T. Yoneda, Daniel J. Friedman, Anne Clayton, Matthew C. Hyman, Renae L. Judy, Allan C. Skanes, Kate M. Orland, Timothy M. Treu, Matthew T. Oetjens, Alvaro Alonso, Elsayed Z. Soliman, Honghuang Lin, Kathryn L. Lunetta, Jesper van der Pals, Tariq Z. Issa, Navid A. Nafissi, Heidi T. May, Peter Leong-Sit, Carolina Roselli, Seung Hoan Choi, FinnGen, Million Veteran Program, Regeneron Genetics Center, Habib R. Khan, Stacey Knight, Richard K. Linnér, Connie R. Bezzina, Samuli Ripatti, J. Michael Gaziano, Ruth Loos, Bruce M. Psaty, J. Gustav Smith, Emelia J. Benjamin, Dan E. Arking, Daniel Rader, Svati H. Shah, Dan M. Roden, Scott M. Damrauer, Lee L. Eckhardt, Jason D. Roberts, Michael J. Cutler, M. Benjamin Shoemaker, Christopher M. Haggerty, Kelly Cho, Aarno Palotie, Peter W.F. Wilson, Patrick T. Ellinor, Steven A. Lubitz

**Affiliations:** Cardiovascular Research Center, Massachusetts General Hospital, Boston, MA, USA; Cardiovascular Disease Initiative, The Broad Institute of MIT and Harvard, Cambridge, MA, USA; VA Boston Healthcare System, Boston, MA, USA; Institute for Molecular Medicine Finland (FIMM), Helsinki Institute of Life Science (HiLIFE), University of Helsinki, Helsinki, Finland; The Broad Institute of MIT and Harvard, Cambridge, MA, USA; Department of Experimental Cardiology, Amsterdam UMC, University of Amsterdam, Amsterdam, The Netherlands; Demoulas Center for Cardiac Arrhythmias, Massachusetts General Hospital, Boston, MA, USA; Gene Regulation Observatory, The Broad Institute of MIT and Harvard, Cambridge, MA, USA; Cardiovascular Medicine Division, Brigham and Women’s Hospital, Boston, MA, USA; VA Atlanta Healthcare System, Decatur, GA, USA; Department of Epidemiology, Emory University Rollins School of Public Health, Atlanta, GA, USA; Heart Institute, Geisinger, Danville, PA, USA; Icahn School of Medicine at Mount Sinai, New York, NY, USA; McKusick Nathans Institute, Department of Genetic Medicine, Johns Hopkins University School of Medicine, Baltimore, MD, USA; Department of Medicine, University of Massachusetts Chan Medical School; The Wallenberg Laboratory/Department of Molecular and Clinical Medicine, Institute of Medicine, Gothenburg University; Department of Cardiology, Sahlgrenska University Hospital, Gothenburg, Sweden; Cardiovascular Health Research Unit, University of Washington, Seattle, WA, USA; Department of Epidemiology, University of Washington, Seattle, WA, USA; Department of Medicine, Division of Cardiovascular Medicine, Vanderbilt University Medical Center, Nashville, TN, USA; Division of Cardiology, Department of Medicine, Duke University School of Medicine, Durham, NC, USA; Intermountain Heart Institute, Intermountain Medical Center, Murray, UT, USA; Division of Cardiac Electrophysiology, Hospital of the University of Pennsylvania, Philadelphia, PA, USA; Department of Surgery, Perelman School of Medicine, University of Pennsylvania, Philadelphia, PA, USA; Section of Cardiac Electrophysiology, Division of Cardiology, Department of Medicine, Western University, London, ON, Canada; Department of Medicine, Division of Cardiovascular Medicine, University of Wisconsin–Madison, Madison, WI, USA; Autism and Developmental Medicine Institute, Geisinger, Lewisburg, PA, USA; Department of Epidemiology, Rollins School of Public Health, Emory University, Atlanta, GA, USA; Epidemiological Cardiology Research Center, Section on Cardiovascular Medicine, Department of Medicine, Wake Forest School of Medicine, Winston-Salem, NC, USA; Department of Medicine, University of Massachusetts Chan Medical School, MA, USA; Department of Biostatistics, Boston University School of Public Health, MA, USA; Department of Cardiology, Clinical Sciences, Lund University and Skane University Hospital, Lund, Sweden; Feinberg School of Medicine, Northwestern University, Chicago, IL, USA; Department of Cardiology, University of Groningen, University Medical Center Groningen, The Netherlands; Section of Cardiac Electrophysiology, Western University, London, ON, Canada; Department of Medicine, University of Utah, Salt Lake City, UT; Department of Economics, Leiden Law School, Leiden University, Leiden, 2531ES, The Netherlands; Department of Experimental Cardiology, Amsterdam Cardiovascular Sciences, Heart Center, Amsterdam University Medical Centre, University of Amsterdam, Amsterdam, The Netherlands; Institute for Molecular Medicine Finland (FIMM), University of Helsinki, Helsinki, Finland; Harvard Medical School, Boston, MA, USA; Brigham and Women’s Hospital, Boston, MA, USA; Charles Bronfman Institute for Personalized Medicine, Icahn School of Medicine at Mount Sinai, New York, NY, USA; Novo Nordisk Foundation Center for Basic Metabolic Research, Department of Health and Medical Sciences, University of Copenhagen, Copenhagen, Denmark; Departments of Medicine, Epidemiology, and Health Services, University of Washington, Seattle, WA, USA; Wallenberg Center for Molecular Medicine and Lund University Diabetes Center, Lund University, Lund, Sweden; Department of Medicine, Boston University Chobanian & Avedisian School of Medicine; Department of Epidemiology, Boston University School of Public Health, MA, USA; NHLBI and BU’s Framingham Heart Study, MA, USA; Division of Cardiovascular Medicine, Department of Medicine, Perelman School of Medicine at the University of Pennsylvania, Philadelphia, PA, USA; Duke Molecular Physiology Institute, Duke University School of Medicine, Durham, NC, USA; Vanderbilt University Medical Center, Nashville, TN, USA; Department of Genetics and Cardiovascular Institute, Perelman School of Medicine, University of Pennsylvania, Philadelphia, PA, USA; Department of Translational Data Science and Informatics, Geisinger, Danville, PA, USA; The Stanley Center for Psychiatric Research and Program in Medical and Population Genetics, The Broad Institute of MIT and Harvard, Cambridge, MA, USA; Analytic and Translational Genetics Unit, Department of Medicine, Department of Neurology and Department of Psychiatry, Massachusetts General Hospital, Boston, MA, USA; Department of Medicine, Emory University School of Medicine, Atlanta, GA, USA

**Keywords:** arrhythmias, genetics

## Abstract

To broaden our understanding of bradyarrhythmias and diseases of the cardiac conduction system, we performed cross-sectional multi-ancestry genome-wide association study meta-analyses in up to 1.3 million individuals for sinus node dysfunction (SND), distal conduction disease (DCD), and pacemaker implantation (PM). We evaluated the biological relevance of bradyarrhythmia loci by analyses of transcriptomes, pleiotropy, and partitioned heritability based on cardiac single cell RNA sequencing data. Finally, we performed rare variant burden testing in 460,000 whole exome sequenced individuals from two biobanks. We identified 13, 28, and 21 common variant loci for SND, DCD, and PM, respectively. Four well-known common variant arrhythmia loci (*SCN5A/SCN10A*, *CCDC141, TBX20*, and *CAMK2D)* were shared for SND and DCD, while other loci were more specific for either SND or DCD. Cardiomyocyte-expressed genes were strongly enriched for contributions to DCD heritability, while SND and PM were more heterogeneous. Rare variant analyses implicated *LMNA* for all bradyarrhythmia subtypes; *SMAD6* and *SCN5A* for DCD; and *TTN*, *MYBPC3*, and *SCN5A* for PM. The genetic architectures of SND and DCD are both overlapping and distinct. Multiple genetic mechanisms involving ion channels, sarcomeric components, cellular homeostasis, and cardiac development may influence the development of bradyarrhythmias.

## Introduction

Bradyarrhythmias are cardiac rhythm abnormalities characterized by pathological conduction slowing and/or slow heart rates, which collectively represent a major public health problem. Bradyarrhythmias may be caused by abnormalities of cardiac impulse formation or conduction. While milder presentations cause symptoms such as fatigue and dizziness, more severe bradyarrhythmias can lead to syncope, heart failure, or sudden cardiac death. Currently, the only established therapy for clinically significant and non-reversible bradyarrhythmias is the implantation of a pacemaker, which is itself associated with an increased risk of complications and morbidity.

The prevalence of bradyarrhythmias has been increasing over time largely due to an aging population, with the incidence of pacemaker implantation (PM) in the United States increasing from 46.7 per 100,000 persons in 1993 to 61.6 per 100,000 persons in 2009.^1^ Based on recent estimates from a large community-based prospective study in the United Kingdom, the prevalence of bradyarrhythmias was 0.5% in the overall population and up to 1.26% among individuals aged ≥65 years.^2^

Bradyarrhythmias have largely been considered a disease of aging, with fibrosis of the conduction system implicated as a major precipitant in prior histopathological samples. However, the molecular causes of bradyarrhythmias are poorly understood.^3^ Furthermore, a subset of conduction system disorders manifest early in life or aggregate in families, with mutations primarily in ion channel or ion channel-associated genes implicated by candidate gene approaches.^4–6^

Past studies that focused on very broad bradyarrhythmia definitions such as the need for a PM,^3^ have had relatively limited power to detect statistically robust associations. One previous genome-wide association study (GWAS) of sinus node dysfunction (SND)—the inability of the sinoatrial node to generate electrical impulses that result in a physiologically adequate atrial rate—identified six genetic loci including a rare susceptibility variant in *MYH6*, a gene encoding a myosin heavy chain isoform.^7^ In contrast, there have been no published GWAS to date for distal conduction disease (DCD), which reflects disease distal to the sinus node, such as atrioventricular node or bundle branch blocks.^8^ The recent availability of genetic data from multiple large-scale studies, along with meta-analyses across cohorts to increase sample size, may enable the detection of common and rare variation underlying bradyarrhythmias with greater physiologic specificity.

In the current study, we performed multi-ancestry meta-analyses of GWAS and rare variant burden tests for SND, DCD, and either entity resulting in the need for a pacemaker, with the goal of elucidating the genetic variation underlying these distinct and clinically relevant conditions.

## Results

**Figure 1** summarizes the study design and depicts the typical anatomical regions in which conduction tissue affected by SND and DCD are localized in the heart. We first began by performing common variant analyses including a total of 1.3M participants for SND and DCD from 10 studies. We conducted additional sensitivity analyses by examining restrictive definitions of SND or DCD (early-onset cases with PM, defined as onset at age ≤75 years) and a combined outcome of PM for SND or DCD. Approximately 90% of individuals were of European ancestry. Baseline characteristics are shown in **Supplemental Tables 1-5**. Manhattan and Miami plots are shown in **Figures 2** and **Supplemental Figures 1-2**. Q-Q plots did not suggest any systematic test statistic inflation (λ<1.10; **Supplemental Figure 3**).

**Figure 1.**
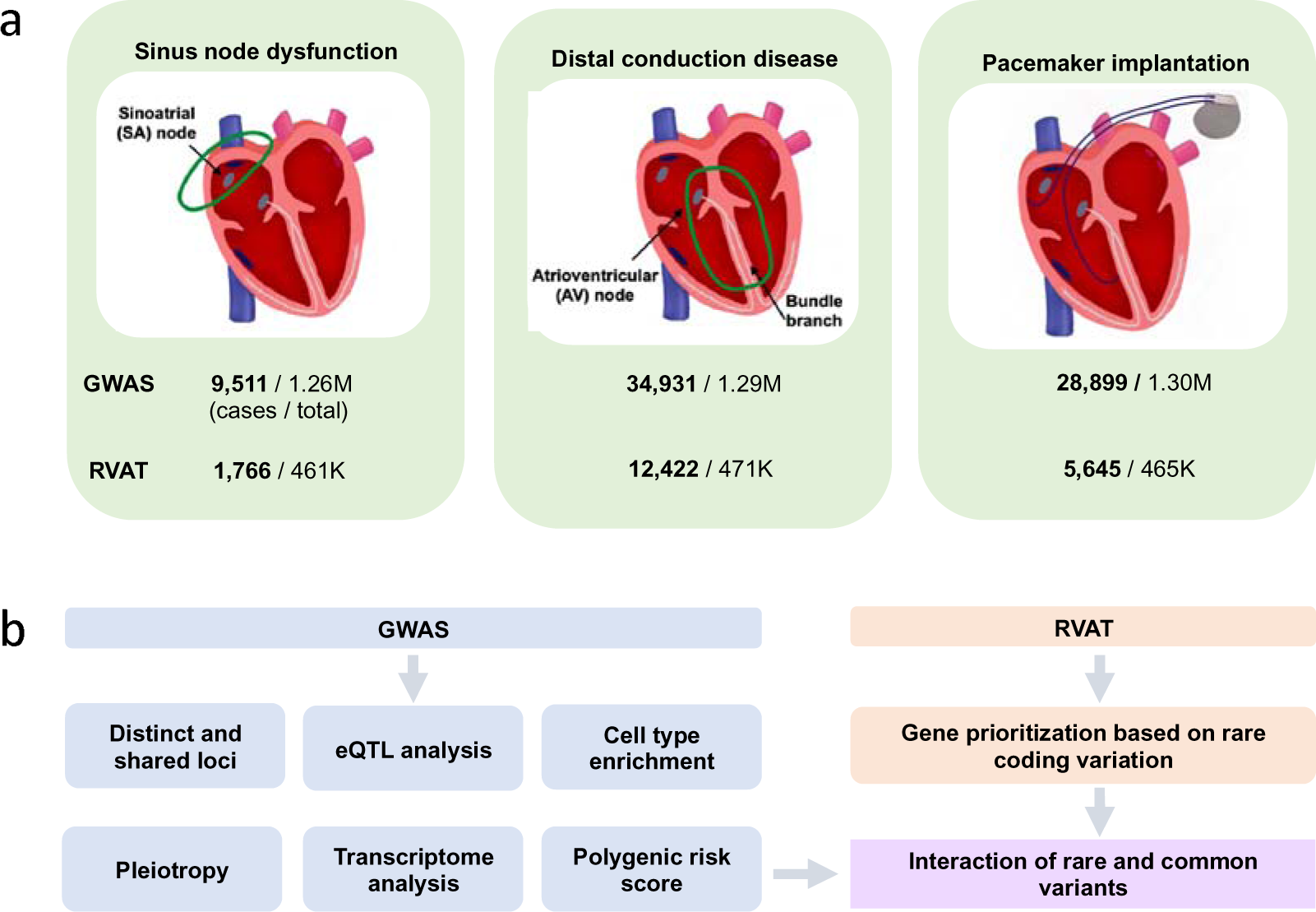
Study design. Panel (a) shows the typical anatomical regions in which conduction tissue affected by sinus node dysfunction and distal conduction disease are localized in the heart. A biventricular pacemaker is also demonstrated. Sample sizes are shown for all three outcomes. Panel (b) shows an overview of common and rare variant analyses for for sinus node dysfunction, distal conduction disease and pacemaker implantation. Common-variant genome-wide association studies (GWAS) were performed in 10 collaborating studies with genotyped and imputed data, and rare variant burden testing was performed for the same phenotypes in 2 studies with whole exome sequencing. The results were combined in meta-analyses including up to 1.3 million individuals for GWAS and 471K individuals for rare variant association testing. For common variant loci reaching genome-wide significance, follow-up evaluations included analyses of cardiac gene expression profiles, pleiotropic associations, predicted transcriptomes, genetic correlations, and polygenic scores. The interaction of rare variants and polygenic risk was further evaluated.

**Figure 2.**
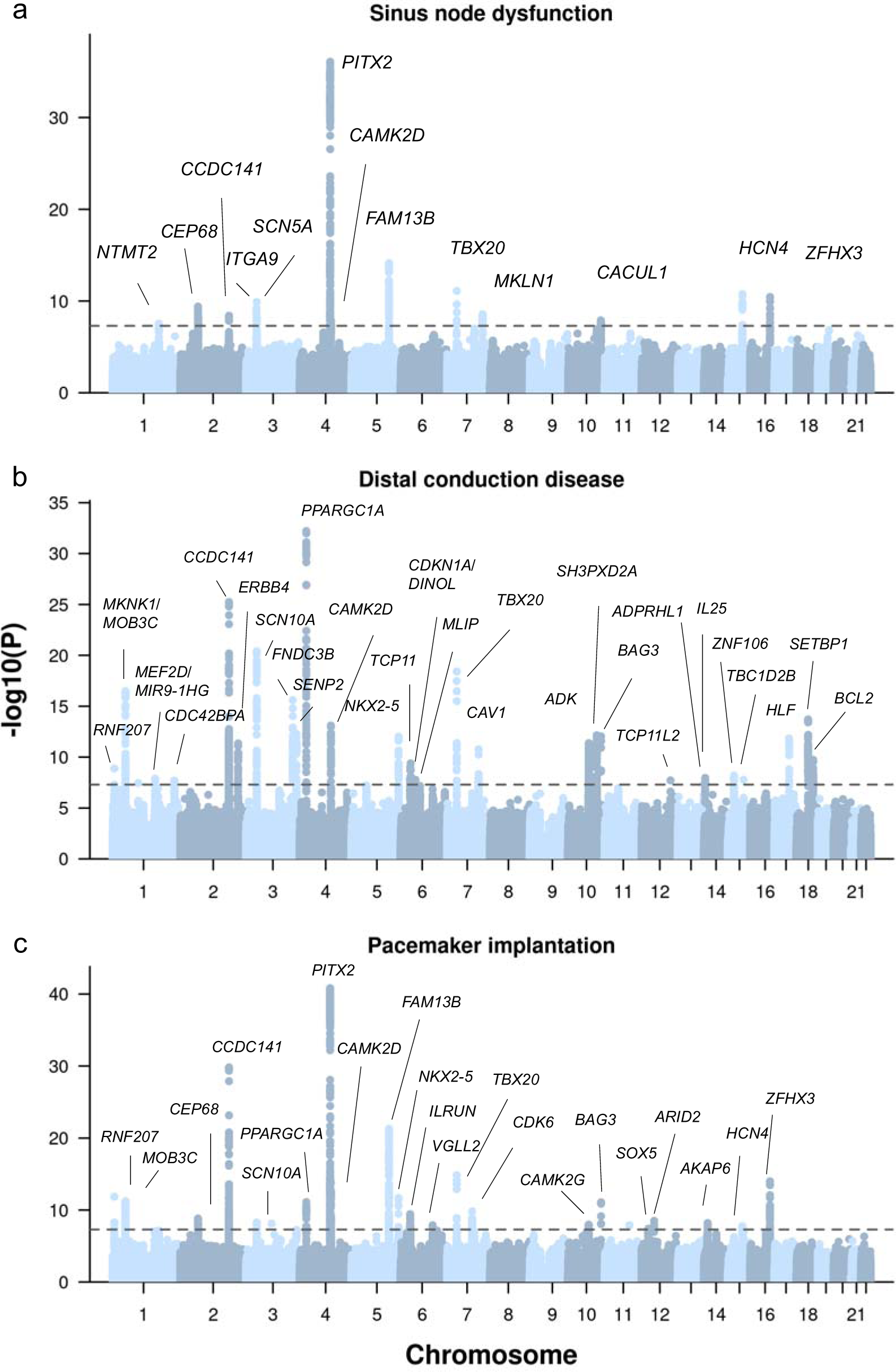
Manhattan plot for bradyarrhythmias. Genome-wide association study results are shown separately for sinus node dysfunction (a), distal conduction disease (b) and pacemaker implantation (c). P-values (on -log10 scale) for each association test between variants and bradyarrhythmias from fixed effect meta-analyses of multi-ancestry individuals are shown on the y-axis. Genome-wide significant association loci (P-value < 5×10^-8^; dashed line) are annotated with the name of the gene closest to the index variant.

### Common variation and sinus node dysfunction

We identified thirteen genome-wide significant loci (P-value <5×10^-8^) associated with SND in 9,511 cases and 1,249,043 controls (**Figure 2**; **Supplemental Table 6**; **Supplemental Figure 1**). When conditioning for the index variants, we observed a secondary signal at two loci (*PITX2* and *SCN5A*/*SCN10A*) (**Supplemental Table 7**). At the *SCN5A*/*SCN10A* locus, both the index variant and the independent variant in the conditional analysis were intronic within *SCN5A*.

A sensitivity analysis using a more restrictive definition of early-onset and PM-dependent SND (4,940 cases) yielded seven of the above genome-wide significant loci and an additional locus close to *MTHFSD* (**Figure 2; Supplemental Table 6; Supplemental Figure 1**). In general, significant index variants were shared or in high LD (r^2^> 0.6 in 1000 Genomes European samples) between the inclusive and restrictive definitions, with larger effect sizes for the restrictive definition (**Supplemental Table 6**). Inclusive and restrictive SND definitions were also highly genetically correlated (r_g_ = 1.03, P-value = 1.49×10^-221^; **Supplemental Table 8**).

Among thirteen SND loci, four (*CCDC141*, *PITX2, ZFHX3*, and *SCN5A*/*SCN10A*) have been reported in a previous GWAS of SND.^9^ Index variants between the two studies were in moderate to high LD (r^2^=0.60-1.00) for three loci but were distinct for the *SCN5A*/*SCN10A* locus (r^2^=0.006). In three loci, index variants or their proxies have been also associated with heart rate or heart rate recovery from exercise (*CCDC141*, *FAM13B/KLHL3* and *HCN4*).^10^ Seven loci have been previously associated with atrial fibrillation (AF),^11–13^ with stronger effects for AF at the *PITX2 and ZFHX3* loci, and stronger or similar effects for SND at 5 loci **(Supplemental Table 9**). We additionally report shared co-associations between SND and prior GWAS of other electrocardiographic and cardiovascular traits,^10^ including pulse and systolic blood pressure *(CEP68* and *MKLN1),* PR interval (*CAMK2D*, *MLKN1* and *HCN4*), electrocardiogram morphology (*PITX2*, *CAMK2D*, *MLKN1*, and *HCN4*), heart rate or heart rate recovery from exercise (*CCDC141*, *FAM13B/KLHL3* and *HCN4*), heart failure (*PITX2*, *ZFHX4*), and stroke (*PITX2*, *CAMK2D*, *FAM13B/KLHL3,* and *HCN4*, *ZFHX3)* **(Supplemental Table 10).**

Significant expression quantitative trait loci (eQTL) are presented in **Supplemental Table 11**. In genetic colocalization analyses, we observed likely shared causal variants (posterior probabilities [PP] >0.80) for SND and the eQTL of cardiac expression at *CEP68*, *CAMK2D*, *FAM13B*, and *CACUL1* **Supplemental Table 11**). Reduced predicted cardiac expression of *FAM13B*, *RP11-325L7.2*, *SRRT*, *GIGYF1*, *CACUL1*, *ZKSCAN1*, *PRRX1*, and *SCN10A* was associated with increased risk of SND, whereas higher predicted cardiac expression of *CEP68* was associated with increased risk of SND (**Supplemental Table 12**).

### Common variation and distal conduction disease

We identified 28 genome-wide significant loci associated with DCD in 34,931 cases and 1,253,984 controls (**Figure 2; Supplemental Table 6**; **Supplemental Figure 2)**. In analyses conditioned on the index variants, we identified additional signals at the *PLEKHA3* (33k bp downstream of *TTN)*, *SCN5A*/*SCN10A*, *TBX20*, and *SH3PXD2A* loci (**Supplemental Table 7**). In contrast to SND, the index variant for DCD at the *SCN5A*/*SCN10A* locus was intronic within *SCN10A*, and we observed a weaker independent signal for a synonymous variant of *SCN5A*.

In sensitivity analyses including 8,629 individuals with restrictive (early-onset and PM-dependent) DCD, we identified six significant loci. Index variants were identical or in moderate LD (r^2^> 0.5) between the inclusive and restrictive DCD definitions, with larger effect sizes for the restrictive definition (**Figure 2; Supplemental Figure 2)**. Inclusive and restrictive DCD definitions were also highly genetically correlated (r_g_ = 0.90, P-value = 3.93×10^-113^; **Supplemental Table 8**).

Next, we evaluated shared co-associations between DCD and prior GWAS of electrocardiographic traits. In total, 10 and 8 of the DCD index variants or their proxies have been previously associated with the PR interval or QRS duration, respectively (**Supplemental Table 10**). Index variants or their proxies at several loci have also been reported to be associated with AF (five loci including three with discordant effect directions for DCD and AF), Brugada syndrome, and other cardiovascular diseases (hypertrophic cardiomyopathy, heart failure, and stroke) in prior GWAS (**Supplemental Tables 9 and 10)**.

In genetic colocalization analyses, we observed likely shared causal variants (PP>0.80) for DCD and the cardiac expression of 10 genes (**Supplemental Table 11**). Higher predicted cardiac gene expression levels of *FOLH1*, *FKBP7*, *STRN*, *MOB3C*, and *C6orf106* were associated with increased risk of DCD, whereas reduced predicted gene expression levels of *COQ8A*, *BCL2*, *MYOZ1, SCN10A, MEF2D*, and *PLEC* were associated with higher risk of DCD (**Supplemental Table 12**).

### Common variation and pacemaker implantation

In addition to disease-specific analyses, we performed a GWAS meta-analysis combining all 28,899 cases with PM for any bradyarrhythmia and identified 21 genome-wide significant loci. Among these loci, 9 overlapped with SND, 9 overlapped with DCD, and 7 were not discovered in the subtype specific analyses (**Figure 2; Supplemental Figure 4; Supplemental Table 6**). We identified secondary signals in four loci (*PLEKHA3*, *PITX2*, *ROS1*, and *TBX20*) in analyses conditioned on the index variants (**Supplemental Table 7**).

In 14 of 21 PM loci, index variants or their proxies have been previously associated with at least one electrocardiographic parameter (including 6 with heart rate, 6 with the PR interval, and 3 with QRS duration) (**Supplemental Table 10)**. In addition, index variants or their proxies in 10 of 21 loci have been previously associated with atrial fibrillation, and three loci (*CCDC141*, *PITX2* and *ZFHX3*) have been associated with sick sinus syndrome.

In genetic colocalization analyses of PM and eQTL of cardiac expression, we observed likely shared causal variants (PP>0.80) at the *CEP68*, *CAMK2D*, *FAM13B*, *and C6orf106* loci (**Supplemental Table 11**). Higher and reduced predicted cardiac expression levels of 7 and 9 genes were associated with increased risk of PM, respectively (**Supplemental Table 12**).

### Common variant genetic correlation analyses

Despite differences in the patterns of genome-wide significant association loci, we observed relatively high overall genetic correlation between SND and DCD (r_g_ = 0.64, P-value = 3.48×10^-25^; **Supplemental Table 8**). This correlation was even higher for the restrictive SND and DCD definitions (r_g_ = 0.85, P-value = 1.18×10^-25^).

We also evaluated the genome-wide correlations between bradyarrhythmias and electrocardiographic intervals. The results differed based on bradyarrhythmia subtypes and were generally consistent with established clinical associations. SND (r_g_ = 0.57, P-value = 4.34×10^-6^) and PM (r_g_ = 0.42, P-value = 1.10×10^-5^) were genetically correlated with resting heart rate, whereas DCD was genetically correlated with conduction times including P-wave duration (r_g_ = 0.34, P-value = 5.22×10^-4^), the PR interval (r_g_ = 0.39, P-value = 1.83 ×10^-6^), and QRS duration (r_g_ = 0.55, P-value = 7.33×10^-8^) (**Supplemental Table 8**). In contrast, corrected QT time was not genetically correlated with any bradyarrhythmia subtype.

### Common variant polygenic score analyses

Next, we evaluated the utility of polygenic scores for bradyarrhythmias. Due to the high genome-wide correlations of bradyarrhythmia subtypes but distinct locus architecture, we limited PRS to genome-wide significant variants in each GWAS meta-analysis to reduce pleiotropic associations. First, we evaluated the associations of each PRS with incident PM during a mean of 11.1 (SD 1.0) years of follow-up after excluding prevalent cases at study baseline and observed that all three PRSs were associated with incident PM (ANOVA P-value in range 4.3×10^-5^ – 4.5×10^-10^). Compared with participants in the bottom tertile of the SND, DCD, and PM PRS, those in the top tertile of each had HRs for PM of 1.24 [1.12–1.37], 1.37 [1.24–1.53], and 1.36 [1.23–1.51], respectively (**Figure 3a**; **Supplemental Table 13**).

**Figure 3.**
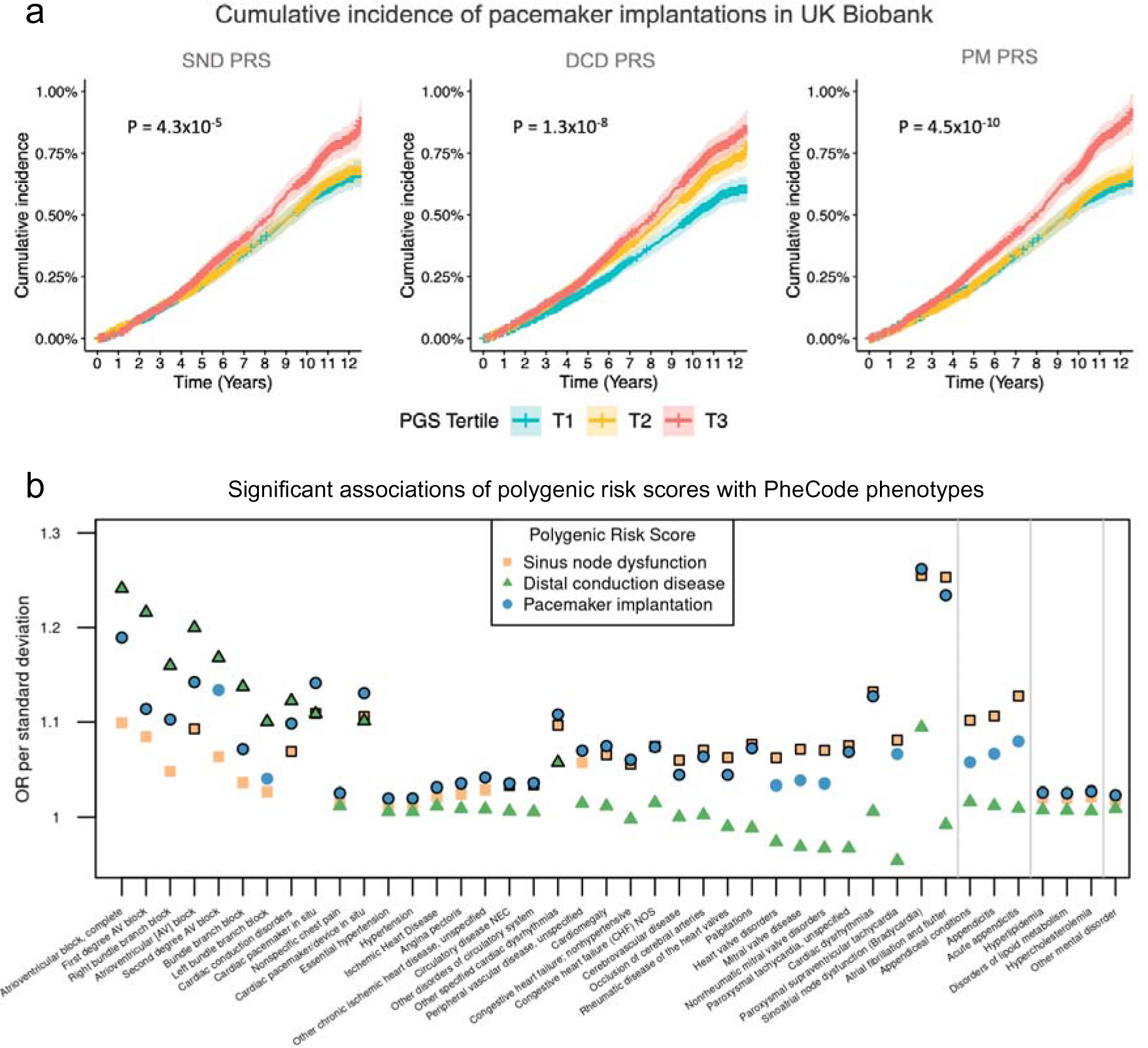
Associations of polygenic scores for bradyarrhythmias with outcomes in unrelated UK Biobank participants. Panel (a) shows the cumulative incidence of pacemaker implantations in UK Biobank participants stratified by polygenic scores for sinus node dysfunction, distal conduction disease and pacemaker implantation. PRSs were constructed using the clumping and thresholding method separately for each phenotype (n_variants for SND PRS_=34; n_variants for DCD_=61; n_variants for PM_=51). A total of 327,707 unrelated participants without pacemakers at study enrollment were included in the analyses. Participants were stratified into three groups based on polygenic score tertiles. P-values for each PRS were derived based on analysis of variance comparing a Cox proportional hazards model with only sex and age as covariates and a Cox proportional hazards model with the polygenic score as an additional covariate. Panel (b) shows the associations of polygenic scores for bradyarrhythmias with a wider set of outcomes based on the PheCode system in 350,877 unrelated individuals in the UK Biobank. Only outcomes with at least one significant association after Bonferroni correction are shown, and significant associations are highlighted with black borders. A total of 65 outcomes were significantly associated with at least one bradyarrhythmia-related polygenic risk score.

We then examined a wider range of cardiovascular outcomes using phecodes from 1329 traits. A PRS for SND was associated with multiple cardiovascular traits, including AF, SND, mitral valve disease, valve disorder, cerebrovascular disease, non-hypertensive heart failure, and hypercholesterolemia (**Figure 3b; Supplemental Table 14**). Furthermore, a PRS for PM was associated with a largely overlapping set of conditions. In contrast, despite a higher number of loci, a PRS for DCD was only associated with atrioventricular block and bundle branch block. All three PRSs were significantly associated with PM.

### Common variation and cell type enrichment in the human heart

To identify relevant cell types contributing to bradyarrhythmias, we used stratified LD Score regression (s-LDSC) with single-nucleus RNA-sequencing data from adult human myocardial samples.^14^ We observed enriched GWAS heritability near cardiomyocyte specific genes for DCD, consistent with findings that the distal conduction system arises from a cardiomyocyte lineage.^15^ (**Extended Data Figure 1; Supplemental Table 15**). In contrast, no individual cell type reached significance for SND or PM, which may be due to limited statistical power or more complex involvement of different cell types.

### Rare variant burden test for bradyarrhythmias

Following the evaluation of common variant contributions to bradyarrhythmias, we next assessed the role of rare protein-disrupting variation using rare variant (MAF<0.1%) burden testing in 460,000 whole exome sequenced individuals from UK Biobank (UKBB) and Mass General Brigham Biobank (MGB) (**Figure 1**). We combined the results in a meta-analysis including a total of 1,766 SND cases, 12,422 DCD cases, 5,645 PM cases, and 459,047 referents. Various rare variant masks were assessed and were combined in a layered approach using the Cauchy distribution test (**Methods**). While the meta-analyses showed some test statistic inflation at the 95th percentile of the test statistic distribution for SND (λ95% = 1.25) and PM (λ95% = 1.14), cohort-specific test statistics were well-calibrated **(Supplemental Figure 5**).

We observed an exome-wide significant (Cauchy P-value < 2.7×10^-6^) burden of rare protein-disrupting variants in *LMNA* for SND and in three genes for DCD (*LMNA*, *SMAD6*, and *SCN5A*) (**Figure 4; Supplemental Table 16a**). Participants with PM carried a higher burden of rare protein-disrupting variants in *LMNA*, *TTN*, *MYBPC3*, and *SCN5A*.

**Figure 4.**
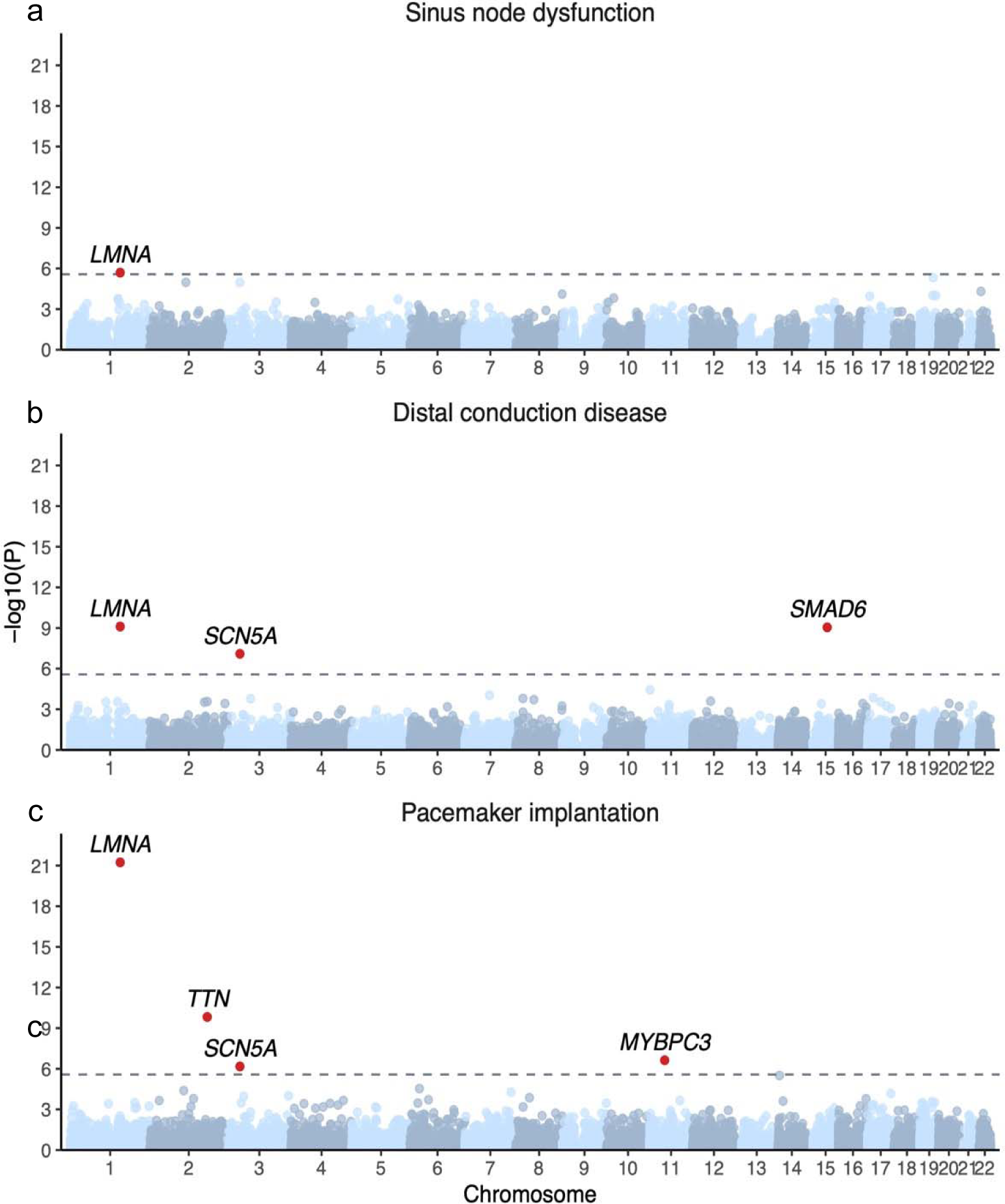
Rare variant association tests. Gene-level results from rare variant burden tests are shown separately for sinus node dysfunction (a), conduction disease (b), and pacemaker implantation (c). P-values (on - log10 scale) for each gene are shown on the y-axis. Dashed lines indicate exome-wide significance thresholds (P-value < 2.7×10-6), and significant genes are highlighted in red.

We conducted additional sensitivity analyses examining individual variation classes (**Supplemental Table 16b)**. In *LMNA*, only missense variants separately and the combination of missense and LOF variants were significantly enriched for all phenotypes (P-values in range of 0.08 – 1.4×10^-6^ for SND, 5.9×10^-5^ 2.4×10^-10^ for DCD; 6.5×10^-4^ – 1.6×10^-22^ for PM), although loss-of-function (LOF) variants were also nominally enriched when examining all exons in patients with DCD or PM (P-value= 0.02 and 0.006, respectively) or canonical transcripts in PM patients (P=0.03; **Supplemental Figure 6).**

For other exome-wide significant genes (*TTN, MYBPC3, SCN5A,* and *SMAD6*), LOF variants had larger effect sizes for the examined bradyarrhythmia phenotypes compared with missense variants (ORs for LOF variants in canonical transcripts in range of 2.10-19.30; ORs for missense variants in canonical transcripts in range of 0.81 – 10.80; **Supplemental Figure 7**). In *SCN5A* and *SMAD6*, however, we observed additional significant or suggestive enrichment of missense variants (Cauchy P-value for missense variants in canonical transcript: P-values in range of 1.4×10^-4^-7.8×10^-4^ for *SCN5A* and P=1.2×10^-5^ for *SMAD6* in DCD). In addition, for every exome-wide significant gene, the effect estimates of missense variants increased commensurate with the proportion of bioinformatics tools predicting a damaging/deleterious effect **(Supplemental Figure 7).** The finding suggests that true damaging missense variants in these genes are associated with bradyarrhythmias. The findings were generally consistent across all 8 tested tissue-specific masks, with the examined variant consequences predicted to affect 80% or more of transcripts in 5–8 tissues per gene.

Because *SMAD6* has been previously associated with congenital cardiovascular malformations that may necessitate invasive cardiac procedures,^16,17^ we performed additional sensitivity analyses to evaluate whether the association with DCD could reflect post-procedural conduction blocks. Supporting a more direct role in DCD, among UK Biobank participants, loss-of-function variants in *SMAD6* were similarly associated with DCD before (OR = 3.4, P-value = 1.2×10^-6^) and after excluding 24,683 participants with prevalent or incident congenital heart disease, cardiac surgery, or stenosis or regurgitation of the aortic, tricuspid or mitral valves (OR = 3.5, P-value = 3.5×10^-6^).

Lastly, we evaluated the proportion of participants with PM during study enrollment or follow-up in unrelated UK Biobank participants who were carriers for either a rare loss-of-function or damaging missense variant in any of the five genes (**Figure 5a, Supplementary Table 17**). A total of 2.11% (95% CI 1.76–2.54%) of rare variant carriers had prevalent or incident PM compared with 0.98% (0.94–1.01%) among non-carriers. Among rare variant carriers, the proportion with PM was highest in those with *LMNA* mutations (6.59%, 4.09–10.36) and lowest in those with *TTN* mutations (1.48%, 1.05–2.06%). When restricting the analyses to loss-of-function variants, the proportion of participants with a PM was greater for a subset of the genes, albeit with much larger confidence intervals (**Supplementary Table 17; Supplemental Figure 8**).

**Figure 5.**
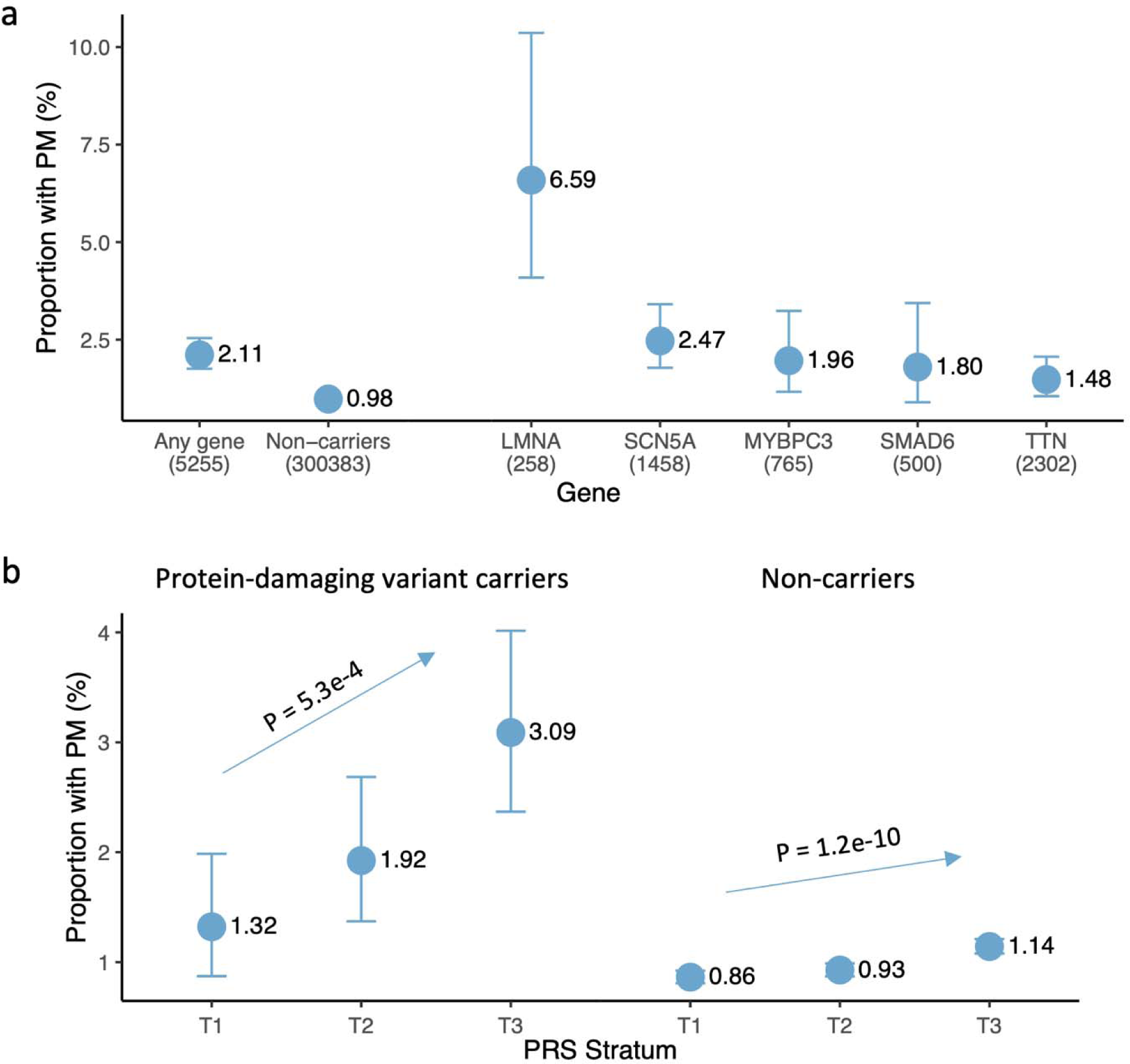
Pacemaker implantations in carriers of protein-damaging variants among 327,707 unrelated UK Biobank participants. Panel (a) shows the proportion of unrelated UK Biobank participants with pacemaker implantations at study enrollment or during follow-up in participants who were carriers of a protein-damaging variant (a loss-of-function variant or a missense variant predicted to be pathogenic by at least 80% of bioinformatics tools) in any of the five genes (*LMNA*, *SCN5A*, *MYBPC3*, *SMAD6*, and *TTN*) that were significantly associated with at least one bradyarrhythmia phenotype. Results are also shown separately for each gene. The Agresti-Coull method was used to calculate binomial 95% confidence intervals. Panel (b) shows the proportion of participants with pacemakers among carriers of a protein-damaging variants (left) and non-carriers (right), stratified by the tertiles of a polygenic score for pacemaker implantation. P-values for the polygenic score were derived with logistic regression using pacemaker implantation as the outcome and the polygenic score tertiles, sex, and age at study enrollment as predictors.

### Overlap and interaction between rare and common variation associated with bradyarrhythmias

Among genes with significant burden of rare protein-disrupting variants in bradyarrhythmia patients, three (*SCN5A*, *LMNA*, and *TTN*) overlapped with GWAS loci. In contrast, no genome-wide significant common variant signal was observed in the loci containing *MYBPC3* or *SMAD6*.

We then evaluated the utility of rare variant burden tests in prioritizing causative genes in GWAS loci (regions within ±1Mb of index variants). Excepting the exome-wide significant *SCN5A*, *LMNA*, and *TTN*, we observed no rare variant signals at a suggestive P-value threshold (**Supplemental Table 18**). However, the well-known cardiomyopathy genes *MYH7* (Cauchy P-values = 6.6×10^-3^ for DCD, 0.28 for SND, and 3.1×10^-6^ for PM) and *NKX2-5* (Cauchy P-values = 1.6×10^-3^ for DCD, 0.51 for SND and 0.07 for PM) were still most strongly prioritized in their respective DCD GWAS loci. In contrast, we observed minimal rare variant signals in some well-established arrhythmia loci such as locus containing *PITX2* (Cauchy P-values in range of 0.07–0.56 for SND, DCD and PM).

Finally, we evaluated the interaction of rare and common variation in unrelated UK Biobank participants. Among 5,255 carriers of a protein-damaging variant (loss-of function or predicted pathogenic missense variant), tertiles of a PRS for PM were associated with an increased likelihood of PM during study enrollment or follow-up (P = 4.1×10^-4^), with those in the top PRS tertile having 2.3-fold risk of PM compared with those in the bottom PRS tertile (3.09% vs 1.32%) (**Figure 5b, Supplementary Table 19**).

## Discussion

We report results from large-scale cross-sectional meta-analyses of bradyarrhythmias in over 1.3 million individuals and 30,000 cases from 10 studies across multiple continents. In total, we identified 13 common variant loci for SND, 28 loci for DCD, and 21 loci for PM. Rare variant association testing in 460,000 participants uncovered five distinct genes influencing susceptibility to bradyarrhythmias. Most of the associations we identified are novel and, consistent with expectations, point to the involvement of ion channel function, cellular homeostasis, and cardiac development in the pathogenesis of bradyarrhythmias. Cardiomyocyte specific genes contributed significantly to DCD heritability, whereas cell specific enrichments were less evident for SND and PM.

Our findings support and extend prior analyses by expanding sample sizes, increasing the specificity of clinically relevant subtypes of bradyarrhythmias, and exploring the shared genetics in cardiac arrhythmias. Although rare familial forms of isolated conduction system disease have been described, only a minority (roughly 5%) of patients have an identifiable mutation,^18^ with the most common variants involving cardiac ion channels (e.g., *SCN5A*,^19^ *TRPM4*,^20^ *HCN4*^6^). Familial conduction diseases can also be associated with cardiomyopathies, in which case mutations are more commonly found in cardiac transcription factors^21^ or structural genes (e.g., *DES*, *LMNA, MYH7,* and *MYBPC3*^18^). Here, we observed robust associations between distinct bradyarrhythmias and rare variants in *SCN5A, LMNA, MYH7* and *MYBPC3.* Moreover, we identified an association between protein-disrupting variants in *SMAD6* and DCD. Smad6 is a structurally distinct member of the Smad family of proteins within the transforming growth factor beta pathway and a preferential inhibitor of bone morphogenic protein responses.^22^ Protein-disrupting mutations in *SMAD6* have previously been linked with congenital cardiac malformations including valve and outflow tract abnormalities.^16,17^ The association with DCD was robust to exclusion of participants with congenital or structural heart disease, suggesting that functioning *SMAD6* is also required for normal development or maintenance of the distal conduction system.

In contrast to rare variation, the relationships between common genetic variation and bradyarrhythmias are less well understood, with no published GWAS of DCD to date and only six loci reported in a previous study of SND.^9^ In the current study, we identified association signals in known monogenic bradyarrhythmia loci (*HCN4* for SND and *SCN5A*/*SCN10A* for SND and DCD), replicated three previously reported loci for SND (*CCDC141*, *PITX2*, and *ZFHX3*), and built upon prior findings by identifying multiple novel and partly distinct common variant associations for SND and DCD. While we observed partial convergence of rare and common variant signals, the GWAS loci are much more numerous. In many loci this likely reflects relative statistical power, but the lack of rare variant association signals may also be due to underrepresentation of individuals with severe bradyarrhythmias in the study populations, or depletion of damaging variants in the general population due to premature mortality or embryonic lethality, such as has been reported for the essential transcription factor *PITX2*.^23^ Thus, common variant analyses, further aided by much larger sample sizes, may facilitate broader biological insights.

Although the loci we identified span a wide range of cardiac biology, our results generally highlight a critical role of cardiac ion channels in the development of bradyarrhythmias. We identified several associations implicating genes related to cardiac ion channels, including *SCN5A, SCN10A*, *HCN4*, *CAMK2D*, and *RNF207*. At the *SCN5A*/*SCN10A* locus, we observe spatially distinct localization of primary signals within *SCN5A* for SND and within *SCN10A* for DCD. Mutations in *SCN5A* are known causes of familial sinus node dysfunction, Long QT syndrome type 3, and Brugada Syndrome.^19^ Non-coding enhancers within SCN10A have been shown to regulate SCN5A expression.^24^ *HCN4*, a gene which encodes an ion channel responsible for spontaneous sinus pacemaker activity and is previously reported for familial sinus bradycardia,^6^ was associated with SND. The ion channel-related proteins *RNF207*, a delayed rectifier and voltage-gated potassium channel regulator, and *CAMK2D*, a kinase regulating myocyte calcium homeostasis and excitation-contraction coupling, do not appear to have been previously associated with bradyarrhythmias, but the locus containing *RNF207* is associated with the QT interval,^25,26^ and knockdown of *RNF207* in zebrafish has been reported to lead to reduced conduction velocity and occasional 2:1 atrioventricular block.^27^

Our results also suggest that common variation in genes relevant to cardiac development and cellular homeostasis appears to broadly influence risk of bradyarrhythmias. Prior analyses have implicated *CCDC141*, which encodes a protein involved in centrosomal function and neural migration, in SND. We expand the number of loci involved in cellular function and bradyarrhythmias. These additional loci include *CEP68* (associated with both SND and DCD), which codes for a protein involved in centrosomal cohesion. Other association loci implicate genes involved in diverse processes such as cell-cell and cell-matrix adhesion (*ITGA9*), and insulin-like growth factor 1 signaling (*GIGYF1*). Protein-damaging mutations in *GIGYF1* have been recently implicated in type 2 diabetes, adverse metabolic health, and clonal mosaicism.^28,29^ Our finding that higher predicted expression of *GIGYF1* is associated with reduced risk of SND is directionally concordant with these reports. Further work is warranted to assess pathways by which alterations in cellular function may lead to bradyarrhythmias. Broadly, our findings suggest that genes involved in cellular function and maintenance appear to influence bradyarrhythmia risk at multiple levels of the cardiac conduction system.

Our results also suggest that differences in the activation or repression of cardiac development and cell fate programs, which have been reported for tachyarrhythmias such as AF and supraventricular tachycardia, are also important for bradyarrhythmias. For example, both *NKX2-5* and *TBX20* encode transcription factors important for the appropriate development of the cardiac septum, which houses critical components of the conduction system. We also observed an association between SND and *PITX2*, a well-known AF susceptibility locus critically involved in promotion of correct left-right differentiation in the developing heart and specification of pulmonary vein myocardial sleeves.^30^ Several other novel associations, such as *ERBB4, BAG3,* and *MLIP*, primarily with DCD and PM, implicate potentially abnormal cardiac developmental programs in risk for bradyarrhythmias.

Although we identified shared loci and genetic correlation across bradyarrhythmias, our analyses also highlight distinct genetic mechanisms underlying bradyarrhythmias originating from the sinus node versus the distal conduction system. Of the 13 significant loci identified for SND and 28 loci identified for DCD, only four (*CAMK2D*, *CCDC141, SCN5A/SCN10A, and TBX20*) were shared between both phenotypes. Genome-wide correlations with electrocardiographic profiles also appeared distinct, as we only observed genetic correlation between SND and resting heart rate, and between DCD and cardiac conduction times. Moreover, we observed greater overlap between SND and AF loci than between DCD and AF loci. Several loci specific to DCD appear to be involved in diverse processes such as cellular apoptosis (e.g., *BCL2*), cellular metabolism (*e.g., PPARGC1A*), and inflammation (e.g., *IL25*), associations which merit further study to define potential mechanisms.

Our results should be interpreted in the context of the study design. First, to maximize statistical power for the relatively rare diseases, we utilized all samples for discovery. Generally consistent variant effect sizes across multiple datasets support the validity of our findings, although future replication in even larger external studies is warranted. Second, participants of some cohorts such as the UK Biobank may be healthier than the population on average. Third, the study participants were predominantly of European ancestry, which may limit the generalizability of our results. We anticipate that larger multi-ancestry biorepositories will enable assessment of the generalizability of our findings. Lastly, we utilized diagnostic and procedural codes to define SND and DCD cases, an approach which is subject to misclassification. We submit that PM (i.e., a procedural outcome) is less subject to misclassification, and therefore the largely consistent results in our restrictive analyses (i.e., those with SND or DCD requiring a PM) are reassuring against major misclassification affecting study results.

In summary, we performed common variant genetic association testing in ten study populations across two continents, in total representing over 1.3 million individuals and 30,000 cases representing either SND, DCD, or PM. In total, we identified 13 genome-wide significant loci for SND, 28 loci for DCD, and 21 loci for PM, and most of the associations identified are novel. Rare variant burden testing identified five genes, including the novel association of *SMAD6* with DCD. Our findings prioritize genes associated with ion channel function, cellular homeostasis, and cardiac development as important potential effectors of risk for bradyarrhythmias and suggest the presence of distinct genetic mechanisms predisposing to sinus node and distal conduction system diseases.

## Online Methods

### Study sample of common variant GWAS

We meta-analyzed common variant GWAS with at least 100 cases from 10 participating studies, and we present methodological details of data collection, genotyping, imputation, and quality controls in the **Supplemental Methods** and **Supplemental Table 20**. All participants provided verbal or written consents, and participating studies were approved by their respective ethics committees or institutional review boards.

### Phenotype definitions

All phenotypes were determined using relevant International Classification of Diseases 9th or 10th revision (ICD-9/10) codes, presence of relevant procedural codes, medical history, or standard 12-lead electrocardiogram data in a study-specific manner as described in the **Supplemental Methods** and **Supplemental Table 21**. SND was defined as the presence of a diagnosis of sinoatrial node dysfunction or sick sinus syndrome. DCD was defined as atrioventricular nodal disease or more distal conduction disease including first degree AV block, second degree AV block, third degree AV block, bundle branch block or combinations of those conditions. PM included codes corresponding to pacemaker implantation, replacement, removal, or interrogation. In secondary analyses, we defined more restrictive early-onset SND and DCD definitions focusing on cases with disease onset age prior to 75 years and a history of PM implantation. We excluded individuals with valvular heart disease, cardiac surgery, or myocardial infarction at or prior to time of bradyarrhythmia diagnosis since these conditions may secondarily cause conduction disease.

### Common variant GWAS

Ancestry-specific common variant GWAS were performed separately in each participating study site. Common variant genetic association testing assumed an additive genetic model, and study-specific statistical models are described in **Supplemental Table 20**.

Post-GWAS quality controls were performed centrally using EasyQC v11.4.^31^ We removed variants with invalid or mismatched alleles from the reference file (1000 Genomes p1v3 EUR/AFR/AMR samples), duplicates, variants with poor imputation quality (INFO < 0.3), rare variants (minor allele count ≤6 or minor allele frequency [MAF] x number of cases x INFO < 10), or variants with invalid summary statistics. To ensure the variant position was consistent across studies, we used LiftOver^32^ to align each variant to Genome Reference Consortium Human Build 37 positions prior to meta-analysis.

### Meta-analysis of common variant GWAS

We performed separate meta-analyses of 6 studies of SND (5 SND restrictive), 8 studies of DCD (5 DCD restrictive), and 9 studies of PM using a fixed-effects approach implemented in METAL.^33^ To control for inflation due to population structure, we applied genomic control to all studies. We removed variants present in only a single study and insertion-deletion variants to avoid mismatch across studies. We set GWAS significant threshold at P-value = 5×10^-8^, and we only report the top (index) variants within a ±1 megabase-pairs [Mb] range or a peak region (if appropriate) with at least 1 supportive variant nearby (P-value < 1×10^-6^). We used the *CMplot*^34^ and *qqman*^35^ packages in R v4.0 to generate Manhattan and Q-Q plots.

We applied the conditional and joint analysis approach in GCTA-cojo to the summary statistics to identify independent signals at identified loci.^36^ Based on the suggestions from GCTA developers (i.e. minimum reference sample size > 4000 individuals), we specifically conditioned on all index variants with linkage disequilibrium (LD) information from 323,061 unrelated European individuals in the UKBB who contributed to all examined phenotypes in this analysis. We included all bi-allelic hard-call transformed (probability>0.8) common single nucleotide polymorphisms with a MAF ≥ 0.01.

### Effect on gene expression and pleiotropic associations

We assessed the association of index variants and their proxies (r^2^≥0.6 in 1000 Genomes p3v5 European participants in a 1000 Kb window) with gene expression in two heart tissues (right atrial appendage and left ventricle) from Genotype-Tissue Expression (GTEx) v8.^37^ We reported all significant eQTL with q-value < 0.05, and we assessed the PP of shared causal variants between GWAS and eQTL results, using the coloc R package.^38^ The testing region comprised all variants in both the eQTL (minimum to maximum position of significant eQTL of the gene) and GWAS (minimum to maximum position of the top GWAS significant variant within ±250 kilobase-pairs [Kb]) regions, and an additional ±5 Kb.

We also performed transcriptome-wide analyses to test associations between predicted gene expression in aforementioned two heart tissues and each phenotype using the elastic-net model in S-Predixcan.^39^ We considered expressed genes significant based on a P-value threshold of 0.05 divided by the total number of tested genes.

We accessed the NHGRI-EBI GWAS Catalog (accessed on Jan 11^th^, 2023) to explore whether index variants or their proxies were previously reported for other cardiovascular disorders.^10^ Additionally, we compared associations for bradyarrhythmias and AF based on summary-level data from large AF meta-analyses.^11–13^ To examine overall genetic links among bradyarrhythmias and with electrocardiographic endophenotypes as reported in prior GWAS,^40^ we performed genetic correlation analyses, using LD score regression with European LD scores from 1000 Genomes provided by LDSC package.^41,42^ To further clarify the genetic association between bradyarrhythmias and other traits, we performed a meta-analysis without individuals from UKBB and derived polygenic risk scores (PRS) for SND, DCD, and PM without ambiguous variants (A/T or C/G) or variant only available in one study, using clumping and thresholding method (P-value cut-off=5×10^-8^, r^2^=0.5 in 1000 Genome p3v5 European participants, window size=2 Mb) in Plink. We calculated PRS by summing the product of effect sizes and allele dosages in the top loci and evaluated the associations of the PRS with incident PM implantations using Cox proportional hazards models with time from study enrollment to diagnosis or censoring as the outcome and sex and age as additional covariates. We removed close relatives, prioritizing the inclusion of PM cases. In addition to the exclusion criteria applied to our main analysis (**Supplemental Table 21**), participants with prevalent SND, DCD, or PM at study enrollment were excluded from incident disease analyses, leaving a total of 327,707 unrelated UKBB European individuals (2,183 with PM) for analyses, with an average 11 years of follow-up. We additionally performed logistic regression on disease outcomes (with at least 100 cases) to estimate the associations from 350,877 unrelated UKBB European individuals, who have complete information of genetic and disease status. The model was adjusted for age at enrollment, genotyping array, male, and first 5 principal components. Disease status was ascertained by the definition from PheCode.^43,44^ Significant P-value thresholds for genetic correlation and PRS-association tests were accounted for multiple testing, using Bonferroni correction.

### Cell type enrichment with stratified LD score regression (s-LDSC)

To identify relevant cell types for bradyarrhythmia GWAS, we used s-LDSC^45^ as described in Finucane et al, 2018^46^. Using single-nucleus RNA-sequencing (snRNAseq) data from Tucker et al, 2020,^14^ we defined cell type specific gene expression profiles by collapsing nuclei into 9 major cell types from the human heart. We tested for differentially expressed genes in each cell type compared to all other cell types by summing gene expression counts for each combination of individual, cell type, and chamber across all nuclei to create a pseudo-bulk expression profile. If a given combination of individual, cell type, and chamber had less than 20 nuclei, it was omitted. Lowly expressed genes were removed using the function filterByExpr() in edgeR.^47^ After DESeq2 normalization,^48^ differential expression testing was performed using the limma-voom framework^49^ with a design of ∼0 + cell_type + individual + chamber and extracting an explicit contrast comparing expression in each cell type to all other cell types. For each cell type, we defined the cell type specific profile as the top 10% most upregulated genes based on the t-statistic from the differential expression test.

We then annotated SNPs near cell type specific genes by building a 100 Kb window on either side of the transcribed region of each gene annotated to a particular cell type, as in Finucane et al, 2018.^46^ All gene coordinates were based on the GRCh38 gene reference used in the snRNAseq data analysis. To test for enriched heritability in regions near cell type specific genes, we mapped GWAS summary statistics to GRCh38 using LiftOver^32^ and ran s-LDSC with our cell type specific annotations along with the baseline model,^45^ using the previously derived 1000 Genomes European ancestry LD reference. To account for the 9 cell types tested for each GWAS trait, we applied a Bonferroni significance cutoff by setting significance at 0.05/9=0.0056.

### Rare variant association tests

We performed exome-wide rare variant burden tests from whole exome sequencing data for SND, DCD, and PM in UKBB and MGB (N_SND cases, UKBB_=803; N_SND cases, MGB_=963; N_DCD cases, UKBB_=9,379; N_DCD cases, MGB_=3,043; N_PM cases, UKBB_=4,091; N_PM cases, MGB_=1,554; N_controls, UKBB_=414,360; N_controls, MGB_=44,687). Details of the datasets, quality controls, and variant annotations are described in **Supplemental Methods.** Within both datasets, we used REGENIE (v3.1.3)^50^ to perform burden tests across various rare variant masks. The REGENIE null-model (step 1) was fit using genome-wide autosomal common variants from genotype array data, adjusting for sex, age, age^2^, sequencing batch, ancestral principal components 1 to 4, as well as any component from 5 to 20 if associated with any of the outcomes. In step 2, a logistic regression model was used to test for the association between rare variant burdens and the outcomes; an approximate Firth’s regression – with back-computation of standard errors – was used for associations reaching nominal *P*<0.05.^50^ The same covariates were applied for step 2, additionally adding the REGENIE PRS as a fixed-effect.^50,51^ Study-specific results were subsequently meta-analyzed using a fixed-effects inverse-variance weighted meta-analysis approach. Prior to meta-analysis, any results with <10 alternative allele carriers were removed, while after meta-analysis mask results with <20 alternative allele carriers were removed; this was done to avoid spurious results from low allele count.

For each gene, up to 198 different masks were tested for association with a given outcome. The different masks were based on two filters for MAF (<0.1% and <0.001%), 11 filters based on variant annotation (LOF, missense with different predicted-deleteriousness cutoffs,^28^ LOF+missense), and 9 filters based on affected transcripts (canonical transcript, all exons, 7 tissue-specific transcripts as determined by pext values). We used a layered approach to combine the many mask-phenotype *P-value*s into a single gene-phenotype *P-value* using the Cauchy distribution test^52^. The Cauchy distribution test allows for valid aggregation of multiple, potentially correlated, test statistics into a single omnibus test statistic. Details on this pipeline, including details on the various filters and Cauchy combination layers, are presented in **Extended Data Figure 2**.

We utilized two approaches to identify bradyarrhythmia-related genes. First, we identified exome-wide significant genes with Cauchy-p value less than 2.7×10^-6^ for each of the phenotypes. For all exome-wide significant genes, we further explored the most relevant variation classes contributing to the Cauchy test (ie, the variant class with the lowest nominal P-value). Second, we evaluated the intersection of rare and common variation by examining the Cauchy P-values of all genes within ±1 Mb of index variants in all GWAS loci. Suggestive significance thresholds for these genes were set by correcting P-values for the number of tested genes across GWAS loci for each phenotype.

We performed additional sensitivity analyses for loss-of-function variation in *SMAD6*, evaluating their association with DCD among UK Biobank participants using logistic regression and identical covariates compared with the discovery analyses. Analyses were performed separately in all whole exome sequenced UK Biobank participants and in participants without prevalent or incident congenital heart disease, cardiac surgery, or stenosis or regurgitation of the aortic, tricuspid or mitral valves.

Finally, we calculated estimates of the proportion of unrelated UKBB participants who had prevalent or incident PM. We derived estimates separately for participants with and without protein-damaging variants (defined as loss-of-function variants or missense variants predicted to be damaging/deleterious by over 80% of bioinformatics tools), and further stratified analyses by tertiles of a polygenic score for PM. We used the Agresti-Coull method to calculate binomial 95% confidence intervals.

## Acknowledgments

The authors thank all participants, study staffs, and participating study centers in this project. Acknowledgements of each participating study are listed in the **Supplemental Data**. The Genotype-Tissue Expression (GTEx) Project was supported by the Common Fund of the Office of the Director of the National Institutes of Health, and by NCI, NHGRI, NHLBI, NIDA, NIMH, and NINDS. The data used for the analyses described in this manuscript were obtained from: Single-Tissue cis-QTL Data from Datasets at the GTEx Portal on 12/17/21. This research has been conducted using the UK Biobank Resource under Application number 17488.

## Sources of Funding

SAL is a full-time employee of Novartis Institutes for BioMedical Research as of July 18, 2022. This work was supported by grants from the National Institutes of Health: R01HL139731 (PTE, SAL, LCW), R01HL157635 (PTE, SAL), RO1HL092577 (KLL, EJB, PTE); from the American Heart Association Strategically Focused Research Networks: 18SFRN34250007 (SAL), 18SFRN34230127 (PTE), 18SFRN34110082 (LCW, EJB); from the American Heart Association: 75N92019D00031 (EJB); and from the European Union: MAESTRIA 965286 (PTE). SJJ is supported by the Junior Clinical Scientist Fellowship from the Dutch Heart foundation (Hartstichting; grant no 03-007-2022-0035). CMH is supported by NIH grants R01HL141901 and R01HL157635. NAN is supported by NIH grant T32HL007101. JTR was supported by a research fellowship from the Sigrid Jusélius foundation. AP was supported by Academy of Finland Centre of Excellence in Complex Disease Genetics (grant no. 312074 and 336824). SMD is supported by IK2-CX001780. MCH is supported by Fondation Leducq (TNE FANTASY 19CV03). This publication does not represent the views of the Department of Veterans Affairs or the United States Government. JGS was supported by grants from the Swedish Heart-Lung Foundation (2022-0344, 2022-0345), the Swedish Research Council (2021-02273), the European Research Council (ERC-STG-2015-679242), Gothenburg University, Skane University Hospital, the Scania county, governmental funding of clinical research within the Swedish National Health Service, a generous donation from the Knut and Alice Wallenberg foundation to the Wallenberg Center for Molecular Medicine in Lund, and funding from the Swedish Research Council (Linnaeus grant Dnr 349-2006-237, Strategic Research Area Exodiab Dnr 2009-1039) and Swedish Foundation for Strategic Research (Dnr IRC15-0067) to the Lund University Diabetes Center. SMD is supported by the US Department of Veterans Affairs Clinical Research and Development award IK2-CX001780. This publication does not represent the views of the Department of Veterans Affairs or the United States Government. Funding of each participating study are listed in the **Supplemental Data**.

## Disclosures

SAL is a full-time employee of Novartis Institutes for BioMedical Research as of July 18, 2022. Previously SAL received sponsored research support from Bristol Myers Squibb / Pfizer, Bayer AG, Boehringer Ingelheim, Fitbit, IBM, Medtronic, and Premier Inc., and consulted for Bristol Myers Squibb / Pfizer, Bayer AG, Blackstone Life Sciences, and Invitae. PTE has received sponsored research support from Bayer AG, IBM Research, Bristol Myers Squibb, Pfizer and Novo Nordisk; he has also served on advisory boards or consulted for Bayer AG and MyoKardia. SMD receives research support for RenalytixAI and has consulted for Calico Labs. The FinnGen project is funded by two grants from Business Finland (HUS 4685/31/2016 and UH 4386/31/2016) and the following industry partners: AbbVie Inc., AstraZeneca UK Ltd, Biogen MA Inc., Bristol Myers Squibb (and Celgene Corporation & Celgene International II Sàrl), Genentech Inc., Merck Sharp & Dohme LCC, Pfizer Inc., GlaxoSmithKline Intellectual Property Development Ltd., Sanofi US Services Inc., Maze Therapeutics Inc., Janssen Biotech Inc, Novartis AG, and Boehringer Ingelheim International GmbH. BMP serves on the Steering Committee of the Yale Open Data Access Project funded by Johnson & Johnson. SMD receives research support from RenalytixAI and Novo Nordisk, outside the scope of the current research. The remaining authors report no disclosures.

## Data availability statement

The GWAS summary statistics are available on the Cardiovascular Disease Knowledge Portal at https://cvd.hugeamp.org/.

## Non-standard Abbreviations and Acronyms

AF: atrial fibrillation
DCD: distal conduction disease
eQTL: expression quantitative trait loci
GTEx: Genotype-Tissue Expression
GWAS: Genome-wide association studies
Kb: kilobase-pairs
LD: linkage disequilibrium
LOF: loss-of-function
MAF: minor allele frequency
Mb: megabase-pairs
MGB: Mass General Brigham Biobank
PM: pacemaker implantation
PRS: polygenic risk scores
RVAT: rare variant association tests
s-LDSC: stratified LD-Score regression
SND: sinus node dysfunction
snRNAseq: single-nucleus RNA-sequencing
UKBB: UK Biobank

## Notes

### Funding Statement

LCW is supported by NIH 1R01HL139731 and American Heart Association 18SFRN34110082. SAL is a full-time employee of Novartis Institutes for BioMedical Research as of July 18, 2022. During this work he was supported by NIH grants R01HL139731, R01HL157635 and American Heart Association 18SFRN34250007. PTE is supported by NIH 1R01HL092577 and K24HL105780, American Heart Association 18SFRN34110082, and the Foundation Leducq 14CVD01. SJJ is supported by the Junior Clinical Scientist Fellowship from the Dutch Heart foundation (Hartstichting; grant no 03-007-2022-0035). CMH is supported by NIH grants R01HL141901 and R01HL157635. NAN is supported by NIH grant T32HL007101. JTR was supported by a research fellowship from the Sigrid Juselius foundation. AP was supported by Academy of Finland Centre of Excellence in Complex Disease Genetics (grant no. 312074 and 336824). SMD is supported by IK2-CX001780. MCH is supported by Fondation Leducq (TNE FANTASY 19CV03). This publication does not represent the views of the Department of Veterans Affairs or the United States Government. JGS was supported by grants from the Swedish Heart-Lung Foundation (2022-0344, 2022-0345), the Swedish Research Council (2021-02273), the European Research Council (ERC-STG-2015-679242), Gothenburg University, Skane University Hospital, the Scania county, governmental funding of clinical research within the Swedish National Health Service, a generous donation from the Knut and Alice Wallenberg foundation to the Wallenberg Center for Molecular Medicine in Lund, and funding from the Swedish Research Council (Linnaeus grant Dnr 349-2006-237, Strategic Research Area Exodiab Dnr 2009-1039) and Swedish Foundation for Strategic Research (Dnr IRC15-0067) to the Lund University Diabetes Center. KLL was supported by NIH 1R01HL092577. EJB was supported by R01HL092577; American Heart Association AF AHA_18SFRN34110082, 75N92019D00031. SMD is supported by the US Department of Veterans Affairs Clinical Reserach and Development award IK2-CX001780. This publication does not represent the views of the Department of Veterans Affairs or the United States Government. Funding of each participating study are listed in the Supplemental Data.

### Author Declarations

The Coordinating Ethics Committee of the Hospital District of Helsinki and Uusimaa (HUS), the UK Biobank Research Ethics Committee, the Mass General Brigham Institutional Review Board, the Human Research Committee of MGB, the Intermountain Health IRB, and the MyCode Community Health Initiative gave ethical approval for this work.

## References

1. Greenspon, A.J. et al. Trends in permanent pacemaker implantation in the United States from 1993 to 2009: increasing complexity of patients and procedures. J Am Coll Cardiol 60, 1540–5 (2012).

2. Khurshid, S. et al. Frequency of Cardiac Rhythm Abnormalities in a Half Million Adults. Circ Arrhythm Electrophysiol 11, e006273 (2018).

3. Celestino-Soper, P.B. et al. Evaluation of the Genetic Basis of Familial Aggregation of Pacemaker Implantation by a Large Next Generation Sequencing Panel. PLoS One 10, e0143588 (2015).

4. Benson, D.W. et al. Congenital sick sinus syndrome caused by recessive mutations in the cardiac sodium channel gene (SCN5A). J Clin Invest 112, 1019–28 (2003).

5. Veldkamp, M.W. et al. Contribution of sodium channel mutations to bradycardia and sinus node dysfunction in LQT3 families. Circ Res 92, 976–83 (2003).

6. Milanesi, R., Baruscotti, M., Gnecchi-Ruscone, T. & DiFrancesco, D. Familial sinus bradycardia associated with a mutation in the cardiac pacemaker channel. N Engl J Med 354, 151–7 (2006).

7. Holm, H. et al. A rare variant in MYH6 is associated with high risk of sick sinus syndrome. Nat Genet 43, 316–20 (2011).

8. Park, D.S. & Fishman, G.I. The cardiac conduction system. Circulation 123, 904–15 (2011).

9. Thorolfsdottir, R.B. et al. Genetic insight into sick sinus syndrome. Eur Heart J 42, 1959–1971 (2021).

10. Buniello, A. et al. The NHGRI-EBI GWAS Catalog of published genome-wide association studies, targeted arrays and summary statistics 2019. Nucleic Acids Res 47, D1005–D1012 (2019).

11. Roselli, C. et al. Multi-ethnic genome-wide association study for atrial fibrillation. Nat Genet 50, 1225–1233 (2018).

12. Nielsen, J.B. et al. Biobank-driven genomic discovery yields new insight into atrial fibrillation biology. Nat Genet 50, 1234–1239 (2018).

13. Christophersen, I.E. et al. Large-scale analyses of common and rare variants identify 12 new loci associated with atrial fibrillation. Nat Genet 49, 946–952 (2017).

14. Tucker, N.R. et al. Transcriptional and Cellular Diversity of the Human Heart. Circulation 142, 466–482 (2020).

15. Mohan, R.A., Boukens, B.J. & Christoffels, V.M. Developmental Origin of the Cardiac Conduction System: Insight from Lineage Tracing. Pediatr Cardiol 39, 1107–1114 (2018).

16. Jin, S.C. et al. Contribution of rare inherited and de novo variants in 2,871 congenital heart disease probands. Nat Genet 49, 1593–1601 (2017).

17. Tan, H.L. et al. Nonsynonymous variants in the SMAD6 gene predispose to congenital cardiovascular malformation. Hum Mutat 33, 720–7 (2012).

18. Ackerman, M.J. et al. HRS/EHRA expert consensus statement on the state of genetic testing for the channelopathies and cardiomyopathies this document was developed as a partnership between the Heart Rhythm Society (HRS) and the European Heart Rhythm Association (EHRA). Heart Rhythm 8, 1308–39 (2011).

19. Priori, S.G. et al. HRS/EHRA/APHRS expert consensus statement on the diagnosis and management of patients with inherited primary arrhythmia syndromes: document endorsed by HRS, EHRA, and APHRS in May 2013 and by ACCF, AHA, PACES, and AEPC in June 2013. Heart Rhythm 10, 1932–63 (2013).

20. Liu, H. et al. Gain-of-function mutations in TRPM4 cause autosomal dominant isolated cardiac conduction disease. Circ Cardiovasc Genet 3, 374–85 (2010).

21. Schott, J.J. et al. Congenital heart disease caused by mutations in the transcription factor NKX2-5. Science 281, 108–11 (1998).

22. Imamura, T. et al. Smad6 inhibits signalling by the TGF-beta superfamily. Nature 389, 622–6 (1997).

23. Gage, P.J., Suh, H. & Camper, S.A. Dosage requirement of Pitx2 for development of multiple organs. Development 126, 4643–51 (1999).

24. van den Boogaard, M., et al. A common genetic variant within SCN10A modulates cardiac SCN5A expression. J Clin Invest 124, 1844–52 (2014).

25. Newton-Cheh, C. et al. Common variants at ten loci influence QT interval duration in the QTGEN Study. Nat Genet 41, 399–406 (2009).

26. Pfeufer, A. et al. Common variants at ten loci modulate the QT interval duration in the QTSCD Study. Nat Genet 41, 407–14 (2009).

27. Roder, K. et al. RING finger protein RNF207, a novel regulator of cardiac excitation. J Biol Chem 289, 33730–40 (2014).

28. Jurgens, S.J. et al. Analysis of rare genetic variation underlying cardiometabolic diseases and traits among 200,000 individuals in the UK Biobank. Nat Genet 54, 240–250 (2022).

29. Zhao, Y. et al. GIGYF1 loss of function is associated with clonal mosaicism and adverse metabolic health. Nat Commun 12, 4178 (2021).

30. Syeda, F., Kirchhof, P. & Fabritz, L. PITX2-dependent gene regulation in atrial fibrillation and rhythm control. J Physiol 595, 4019–4026 (2017).

31. Winkler, T.W. et al. Quality control and conduct of genome-wide association meta-analyses. Nat Protoc 9, 1192–212 (2014).

32. Hinrichs, A.S. et al. The UCSC Genome Browser Database: update 2006. Nucleic Acids Res 34, D590–8 (2006).

33. Willer, C.J., Li, Y. & Abecasis, G.R. METAL: fast and efficient meta-analysis of genomewide association scans. Bioinformatics 26, 2190–1 (2010).

34. Yin, L. et al. rMVP: A Memory-efficient, Visualization-enhanced, and Parallel-accelerated Tool for Genome-wide Association Study. Genomics Proteomics Bioinformatics 19, 619–628 (2021).

35. Turner, S.D. qqman: an R package for visualizing GWAS results using Q-Q and manhattan plots. bioRxiv, 005165 (2014).

36. Yang, J. et al. Conditional and joint multiple-SNP analysis of GWAS summary statistics identifies additional variants influencing complex traits. Nat Genet 44, 369–75, S1-3 (2012).

37. Carithers, L.J. et al. A Novel Approach to High-Quality Postmortem Tissue Procurement: The GTEx Project. Biopreserv Biobank 13, 311–9 (2015).

38. Giambartolomei, C. et al. Bayesian test for colocalisation between pairs of genetic association studies using summary statistics. PLoS Genet 10, e1004383 (2014).

39. Barbeira, A.N. et al. Exploring the phenotypic consequences of tissue specific gene expression variation inferred from GWAS summary statistics. Nat Commun 9, 1825 (2018).

40. Choi, S.H., et al. Rare Coding Variants Associated With Electrocardiographic Intervals Identify Monogenic Arrhythmia Susceptibility Genes: A Multi-Ancestry Analysis. Circ Genom Precis Med 14, e003300 (2021).

41. Bulik-Sullivan, B.K. et al. LD Score regression distinguishes confounding from polygenicity in genome-wide association studies. Nat Genet 47, 291–5 (2015).

42. Bulik-Sullivan, B. et al. An atlas of genetic correlations across human diseases and traits. Nat Genet 47, 1236–41 (2015).

43. Wu, P. et al. Mapping ICD-10 and ICD-10-CM Codes to Phecodes: Workflow Development and Initial Evaluation. JMIR Med Inform 7, e14325 (2019).

44. Wei, W.Q. et al. Evaluating phecodes, clinical classification software, and ICD-9-CM codes for phenome-wide association studies in the electronic health record. PLoS One 12, e0175508 (2017).

45. Finucane, H.K. et al. Partitioning heritability by functional annotation using genome-wide association summary statistics. Nat Genet 47, 1228–35 (2015).

46. Finucane, H.K. et al. Heritability enrichment of specifically expressed genes identifies disease-relevant tissues and cell types. Nat Genet 50, 621–629 (2018).

47. Robinson, M.D., McCarthy, D.J. & Smyth, G.K. edgeR: a Bioconductor package for differential expression analysis of digital gene expression data. Bioinformatics 26, 139–40 (2010).

48. Love, M.I., Huber, W. & Anders, S. Moderated estimation of fold change and dispersion for RNA-seq data with DESeq2. Genome Biol 15, 550 (2014).

49. Law, C.W., Chen, Y., Shi, W. & Smyth, G.K. voom: Precision weights unlock linear model analysis tools for RNA-seq read counts. Genome Biol 15, R29 (2014).

50. Mbatchou, J. et al. Computationally efficient whole-genome regression for quantitative and binary traits. Nat Genet 53, 1097–1103 (2021).

51. Jurgens, S.J. et al. Adjusting for common variant polygenic scores improves yield in rare variant association analyses. Nat Genet 55, 544–548 (2023).

52. Liu, Y. et al. ACAT: A Fast and Powerful p Value Combination Method for Rare-Variant Analysis in Sequencing Studies. Am J Hum Genet 104, 410–421 (2019).

