## Supplement for "Common and Rare Variant Contributions to Bradyarrhythmias from Multi-Ancestry Meta-Analyses"

### SUPPLEMENTARY METHODS

#### Study descriptions

Sample selection for AGENT (including BioVu, Duke, Intermountain, Penn, Western Ontario, and Wisconsin), BIOME, FINNGEN, Geisinger MyCode, Million Veteran Program (MVP), and UK Biobank (UKBB) are described previously.[1] Disease status was obtained from electronic health record and ascertained using ICD and CPT code provided in **Supplemental table 2**. Individuals who have at least two records of selected ICD code or at least one selected CPT code were defined as cases, unless otherwise mentioned.

#### AGENT (Intermountain)

Intermountain Health patients were recruited prospectively to provide blood as part of the INSPIRE biological registry, which is an ongoing DNA and plasma biorepository. The recruitment was mainly through the cardiac catheterization labs and cardiology clinics. Patients were recruited using written informed consent approved for the INSPIRE biological registry by the Intermountain Health IRB. For the current study, electronic medical records were queried to identify patients in the INSPIRE registry with a diagnosis of sinus node disease and distal conduction disease using ICD-9 or ICD-10 codes in combination with EKG reports of first degree AV block, second degree AV block, third degree AV block, bundle branch block, or fascicular block.

**ARIC**

ARIC is a prospective community-based study of cardiovascular disease and its risk factors. At baseline (1987-1989), 15,792 men and women aged 45 to 64 were recruited from four US communities (Washington County, Maryland; Forsyth County, North Carolina; Jackson, Mississippi; Minneapolis suburbs, Minnesota) and underwent extensive clinic examinations. Presence of distal conduction disease was defined as the presence of 2 or more ICD-9-CM or ICD-10-CM codes for first degree AV block, second degree AV block (any level), third degree AV block, bundle branch block, fascicular block, or combinations of these conditions in any hospitalization. Events occurring in those with valvular heart disease, cardiac surgery, or myocardial infarction at or prior to time of phenotype diagnosis were excluded.

Hypertension was defined as having a systolic blood pressure ≥ 140 mmHg or diastolic blood pressure ≥ 90 mmHg as measured during the exam (following cohort protocol) or participants indicated medication was used within the two weeks prior to examination. Individuals were noted as being diabetic if fasting glucose levels were at least 126 mg/dL or non-fasting glucose levels were at least 200 mg/dL. Additionally, self-reported physician diagnosis of diabetes or diabetic medication use were used to for the definition. Heart failure was defined based on the Gothenburg Criteria and medication usage. Atrial fibrillation diagnoses were ascertained by exam ECG.

**CHS**

The Cardiovascular Health Study (CHS) is a population-based longitudinal study of risk factors, progression, and outcomes of CVD in 5888 community participants aged 65 years and older (42% male, 85% white) recruited from four US communities: Forsyth County, North Carolina; Sacramento County, California; Washington County, Maryland; and Pittsburgh, Pennsylvania. Pacemaker implantation was based on the presence of the following ICD9 codes in any hospitalization: 37.8 (insertion, placement and revision of pacemaker), V45.01 (status post-pacemaker implantation), or V53.31 (fitting and adjustment of cardiac pacemaker).

**FHS**

The Framingham Heart Study (FHS) is a prospective community-based study designed to examine the risk factors and outcomes associated with cardiovascular diseases. There were three generations of participants: the original cohort (initiated in 1948), the offspring (initiated in 1971), and the third generation (initiated in 2002). Clinical examination included standard interviews, physical exams, ECG, and laboratory evaluations. Standard 12-lead ECGs have been conducted on FHS participants and reviewed by FHS cardiologists as well as outside medical records. The distal conduction disorder was defined as an atrioventricular nodal disease or more distal disease including first-degree AV block, second-degree AV block (any level), third-degree AV block, and some ventricular conduction abnormalities such as IV blocks, pattern, complete, hemiblock abnormalities by the 12-lead ECG in each examination. The pacemaker was defined as the insertion of a permanent pacemaker for sinus node dysfunction or atrioventricular block or distal conduction disease by the cardiovascular diseases procedures records.

**Geisinger MyCode**

The MyCode Community Health Initiative is an IRB-approved research study of the Geisinger health system in central and northeastern Pennsylvania. Started in 2007, the study is open to any Geisinger patient through opt-in informed consent. Through the DiscovEHR collaboration with Regeneron Genetics Center, data from exome sequencing and genome-wide genotyping are available and have been linked with health information from the Geisinger electronic health record. The genotype data used for this analysis were derived from filtering and QC performed on the first 145K genotyped individuals in the cohort. Genotyping was performed on either Illumina HumanOmniExpress or Global Screening Array platforms. Pre-imputation variant filters included MAF > 0.01, HWE p > 1e-15, and site-level missingness < 0.01. Iterative principal component analysis, with 1000 Genomes phase 3 version 5 data as reference panel, classified genetic ancestry and identified a homogenous subset of 122,370 samples that clustered tightly with the CEU, TSI, GBR populations in 1000 Genomes (and later passed the pre-GWAS QC, described next). Data were imputed, by platform, for the European-ancestry subset using the Human Reference Consortium panel via the Michigan Imputation Server. Post-imputation, data from the two platforms were filtered to include non-duplicated bi-allelic SNPS with MAF ≥0.001, INFO ≥0.2, and average maximum posterior call ≥0.90. Additional pre-GWAS QC filters included sample missingness, PC-adjusted heterozygosity, and sex mismatch. Lastly, per-chromosome files were merged across the two platforms (all down-stream analyses were adjusted with a dummy variable). The genetic variance component of the linear mixed models was modelled with a lightly LD-pruned (--indep-pairwise 1000 80 0.7) set of 447,896

QC'd SNPs (MAF > 0.01, HWE P > 5e–6, site-level missingness <0.05). To capture subtle population structure, the principal components used in association analysis were re-estimated and evaluated within the homogenous EUR sample (N = 122,370).

Phenotypes were defined based on the presence of ICD codes on two or more patient encounters from the Geisinger electronic health record.

**MDCS**

MDCS is a community-based prospective cohort of middle-aged individuals from Southern Sweden. In total, 30,447 subjects attended a baseline exam in 1991-1996 when they filled out a questionnaire, underwent anthropometric measurements and donated peripheral venous blood samples (PMID 19936945). The study was approved by the local ethics committee and all participants provided written informed consent. Prevalent or incident cases of bradyarrhythmia were ascertained from nation-wide hospital registers based on ICD-9 or ICD-10 codes (PMID 21070922). Genome-wide genotyping of single nucleotide variants in the full cohort was by the Regeneron Genetics Center performed using the Illumina Human Omni Express Exome BeadChip kit.

**FinnGen**

FinnGen is a collection of prospective Finnish epidemiological and disease-based cohorts and hospital biobank samples, aiming to collect genotype samples from 500,000 individuals by 2023. The FinnGen data used in this study comprise 377,277 individuals from FinnGen Data Freeze 9 (https://www.finngen.fi/en). The data were linked by unique national personal identification numbers to the national hospital discharge registry (available from 1968), the cause of death registry (1969-), medication purchase (1995-) and reimbursement (1964-) registries, the specialist outpatient registry (1998-), and the registry of primary health care visits (2011-). Cases with sinus node dysfunction, distal conduction disease or pacemakers were identified according to the harmonized meta-analysis protocol based on ICD-9 codes, ICD-10 codes and NOMESCO codes corresponding to surgical procedures. For each phenotype, a minimum of two ICD or NOMESCO code based instances were required to determine case status. Individuals not meeting any of these case criteria were designated as controls for all analyses. Participants with at least two instances of ICD or NOMESCO codes corresponding to valvular heart disease, cardiac surgery or myocardial infarction at or prior to the index events were additionally excluded from cases (if such events occurred during or before the case index event) and from controls.

The FinnGen samples were genotyped using Illumina and Affymetrix arrays (Illumina Inc., San Diego, and Thermo Fisher Scientific, Santa Clara, CA, USA). Genotype imputation was performed using a population-specific SISu v4 imputation reference panel comprised of 8,557 whole genome sequences. The genotype imputation protocol is available at: https://dx.doi.org/10.17504/protocols.io.xbgfijw.

Patients and control subjects in FinnGen provided informed consent for biobank research, based on the Finnish Biobank Act. Alternatively, separate research cohorts, collected prior the Finnish Biobank Act came into effect (in September 2013) and start of FinnGen (August 2017), were collected based on study-specific consents and later transferred to the Finnish biobanks after approval by Fimea (Finnish Medicines

Agency), the National Supervisory Authority for Welfare and Health. Recruitment protocols followed the biobank protocols approved by Fimea. The Coordinating Ethics Committee of the Hospital District of Helsinki and Uusimaa (HUS) statement number for the FinnGen study is Nr HUS/990/2017. The FinnGen study is approved by Finnish Institute for Health and Welfare (permit numbers: THL/2031/6.02.00/2017, THL/1101/5.05.00/2017, THL/341/6.02.00/2018, THL/2222/6.02.00/2018, THL/283/6.02.00/2019, THL/1721/5.05.00/2019 and THL/1524/5.05.00/2020), Digital and population data service agency (permit numbers: VRK43431/2017-3, VRK/6909/2018-3, VRK/4415/2019-3), the Social Insurance Institution (permit numbers: KELA 58/522/2017, KELA 131/522/2018, KELA 70/522/2019, KELA 98/522/2019, KELA 134/522/2019, KELA 138/522/2019, KELA 2/522/2020, KELA 16/522/2020), Findata permit numbers THL/2364/14.02/2020, THL/4055/14.06.00/2020,,THL/3433/14.06.00/2020, THL/4432/14.06/2020, THL/5189/14.06/2020, THL/5894/14.06.00/2020, THL/6619/14.06.00/2020, THL/209/14.06.00/2021, THL/688/14.06.00/2021, THL/1284/14.06.00/2021, THL/1965/14.06.00/2021, THL/5546/14.02.00/2020, THL/2658/14.06.00/2021, THL/4235/14.06.00/202, Statistics Finland (permit numbers: TK-53-1041-17 and TK/143/07.03.00/2020 (earlier TK-53-90-20) TK/1735/07.03.00/2021, TK/3112/07.03.00/2021) and Finnish Registry for Kidney Diseases permission/extract from the meeting minutes on 4th July 2019.

The Biobank Access Decisions for FinnGen samples and data utilized in FinnGen Data Freeze 9 include: THL Biobank BB2017_55, BB2017_111, BB2018_19, BB_2018_34, BB_2018_67, BB2018_71, BB2019_7, BB2019_8, BB2019_26, BB2020_1, Finnish Red

Cross Blood Service Biobank 7.12.2017, Helsinki Biobank HUS/359/2017, HUS/248/2020, Auria Biobank AB17-5154 and amendment #1 (August 17 2020), AB20-5926 and amendment #1 (April 23 2020) and it´s modification (Sep 22 2021), Biobank Borealis of Northern Finland_2017_1013, Biobank of Eastern Finland 1186/2018 and amendment 22 § /2020, Finnish Clinical Biobank Tampere MH0004 and amendments (21.02.2020 & 06.10.2020), Central Finland Biobank 1-2017, and Terveystalo Biobank STB 2018001 and amendment 25$^{th}$ Aug 2020.

**UKBB**

Disease status in UK Biobank was ascertained by the selected ICD and CPT code. Individuals who have one or more ICD/CPT code are defined as cases.

**Statistical analysis**

We performed the primary GWAS on the AGENT, the UK Biobank, and MVP, using logistic regression in PLINK2.0.[2] GWAS of other participating studies were performed at each participating study site with study-specific model and covariates. Details of data collection, genotyping and imputation, and data analysis are described in prior studies[1] and **Supplemental Table 20**. We removed 2$^{nd}$ degree or closer relatives and prioritized retention of cases over controls when excluding related individuals. The regression models were stratified by race and adjusted for age (standardized in AGENT and MVP), sex (standardized in AGENT), and first 5 principal components (standardized in AGENT) to account for population structure.

### Rare variant study datasets

*UK Biobank*

The UK Biobank (UKB) is a large population-based prospective cohort study from the United Kingdom that included over 500,000 individuals with deep phenotypic data, including medical interviews, electronic health record linkage and death registry linkage.[3,4] Participants were recruited between 2006 and 2010 at ages of 40-69 years.[4] Relevant genomic data currently includes exome sequencing on over 450,000 samples funded through industry partnerships.[5,6] Exomes were captured using the revised version of the IDT xGen Exome Research Panel v1.0 on Illumina NovaSeq 6000 machines (https://www.ukbiobank.ac.uk/media/najcnoaz/access_064-uk-biobank-exome-release-faq_v11-1_final-002.pdf). Alignment using BWA-MEM, calling using DeepVariant, and joint genotyping using GLNexus have been described in detail elsewhere (https://biobank.ndph.ox.ac.uk/showcase/ukb/docs/UKB_WES_Protocol.pdf). In the present study, we utilized the OQFE exome call set and closely followed a previously published pipeline to perform stringent quality-control (QC) of the exome sequencing data, including genotype QC, variant QC and sample QC.[7] After quality-control, we were left with 454,162 samples with WES data who could be linked to their phenotypic data. The UK Biobank resource was approved by the UK Biobank Research Ethics Committee and all participants provided written informed consent to participate. Use of UKB data was performed under application number 17488 and was approved by the Mass General Brigham Institutional Review Board.

*Mass General Brigham Biobank*

The Mass General Brigham Biobank (MGB; formerly known as Partners Biobank) is an ongoing observational research project enrolling participants from a multicenter health system in Eastern Massachusetts.[8] Participants are enrolled with broad-based consent collected by local research coordinators, either as part of a collaborative research study or electronically through a patient portal.[9] Demographic data, blood samples and surveys are collected at baseline and linked to electronic health record data. All adult patients provided informed consent to participate. A small number of children were enrolled with IRB-approved assent forms; upon reaching 18 years of age all enrolled children had to provide consent or were removed from the study. The Human Research Committee of MGB approved the Biobank protocol (2009P002312). Exome sequencing, as part of NHGRI's *Center for Common Disease Genomics*, has currently been completed for over 53,000 MGB participants. Samples were sequenced on Illumina NovaSeq machines with a custom exome panel (TWIST Human Core Exome), with a target of at least 20X coverage at >85% of target sites. Alignment, processing and joint-calling of variants were performed using the Genome Analysis ToolKit (GATK v4.1) following GATK best practices, after which we applied a stringent QC pipeline on the sequencing data, closely following the pipeline applied in the UK Biobank. After stringent QC, we were left with 51,815 samples that could be matched to their electronic health records.

**Variant annotation**

In each dataset, variants were annotated using dbNSFP (v.4.2a for MGB and v.4.3a for UKB)[10] and the Loss-of-Function Transcript Effect Estimator (LOFTEE[11]) plug-in implemented in the Variant Effect Predictor (VEP; v.105)[12] (https://github.com/konradjk/loftee). VEP was used to ascertain the most severe consequences for all annotated gene transcripts. LOFTEE was implemented to identify high-confidence LOF variants, which include frameshift indels, stop-gain variants and splice site disrupting variants. LOFs flagged by LOFTEE as dubious were removed. Missense variants were assigned a missense score representing the proportion of bioinformatics tools predicting a damaging effect, following previously published methods[6]. In short, we used information from 30 tools included in the dbNSFP database to score each missense variant by the number of tools predicting a damaging/deleterious effect, and divided this value by the number of tools that gave a prediction. Missense variants with <7 predictions were removed. For instance, if 14 tools predicted a damaging effect and 28 total tools gave a prediction, then the missense score would equal 0.5 (14/28). Finally, variants were annotated with the highest continental allele frequency from gnomAD v2 exomes (extracting frequencies for EUR, EAS, SAS, AFR and AMR super-populations) denoted as 'gnomAD popmax'[10].

We used the *tx_annotation* tool (https://github.com/macarthur-lab/tx_annotation) to annotate variants with their tissue-specific 'proportion expression across transcripts' (pext) values.[13] Pext values represent the proportion of gene transcripts predicted to be affected by a given variant consequence, based on isoform expression, in a specific tissue.[13] We utilized isoform expression data from the GTEx version 8 RNAseq dataset[14] (reported as transcripts per million mapped reads from the RSEM v1.3.0 software), and applied the *get_gtex_summary()* function from *tx_annotation* to compute median isoform expression for each GTEx tissue; the maximum gene expression for each tissue was computed by feeding the median isoform expression values into the *get_gene_expression()* function. Finally, pext values were extracted for the most severe consequence of each variant using the *tx_annotate_mt()* function, specifying the "proportion" flag. In the current manuscript, we used a tissue-specific pext cutoff of >=0.8 to assign variants as gene-affecting. In other words, if a given variant consequence was predicted to affect 80% or more of gene transcripts in a tissue, we declared the variant as affecting the gene in that tissue. Here, we focused on a few tissues of interest: left ventricle, atrial appendage, aorta, coronary artery, tibial artery, whole blood, and the mean across all GTEx tissues.

**Study-specific acknowledgements**

**ARIC**

The Atherosclerosis Risk in Communities study has been funded in whole or in part with Federal funds from the National Heart, Lung, and Blood Institute, National Institutes of Health, Department of Health and Human Services, under Contract nos. (75N92022D00001, 75N92022D00002, 75N92022D00003, 75N92022D00004, 75N92022D00005). Funding was also supported by R01HL087641 and R01HL086694; National Human Genome Research Institute contract U01HG004402; and National Institutes of Health contract HHSN268200625226C. Infrastructure was partly supported by Grant Number UL1RR025005, a component of the National Institutes of Health and

NIH Roadmap for Medical Research. The authors thank the staff and participants of the ARIC study for their important contributions.

**FinnGen**

We want to acknowledge the participants and investigators of FinnGen study. The FinnGen project is funded by two grants from Business Finland (HUS 4685/31/2016 and UH 4386/31/2016) and the following industry partners: AbbVie Inc., AstraZeneca UK Ltd, Biogen MA Inc., Bristol Myers Squibb (and Celgene Corporation & Celgene International II Sàrl), Genentech Inc., Merck Sharp & Dohme LCC, Pfizer Inc., GlaxoSmithKline Intellectual Property Development Ltd., Sanofi US Services Inc., Maze Therapeutics Inc., Janssen Biotech Inc, Novartis AG, and Boehringer Ingelheim International GmbH. Following biobanks are acknowledged for delivering biobank samples to FinnGen: Auria Biobank (www.auria.fi/biopankki), THL Biobank (www.thl.fi/biobank), Helsinki Biobank (www.helsinginbiopankki.fi), Biobank Borealis of Northern Finland (https://www.ppshp.fi/Tutkimus-ja-opetus/Biopankki/Pages/Biobank-Borealis-briefly-in-English.aspx), Finnish Clinical Biobank Tampere (www.tays.fi/en-US/Research_and_development/Finnish_Clinical_Biobank_Tampere), Biobank of Eastern Finland (www.ita-suomenbiopankki.fi/en), Central Finland Biobank (www.ksshp.fi/fi-FI/Potilaalle/Biopankki), Finnish Red Cross Blood Service Biobank (www.veripalvelu.fi/verenluovutus/biopankkitoiminta), Terveystalo Biobank (www.terveystalo.com/fi/Yritystietoa/Terveystalo-Biopankki/Biopankki/) and Arctic Biobank (https://www.oulu.fi/en/university/faculties-and-units/faculty-medicine/northern-finland-birth-cohorts-and-arctic-biobank). All Finnish Biobanks are members of BBMRI.fi infrastructure (www.bbmri.fi). Finnish Biobank Cooperative -FINBB (https://finbb.fi/) is the coordinator of BBMRI-ERIC operations in Finland. The Finnish biobank data can be accessed through the Fingenious® services (https://site.fingenious.fi/en/) managed by FINBB.

**Study specific funding**

**AGENT (BioVU)**

Vanderbilt University Medical Center's BioVU projects are supported by numerous sources: institutional funding, private agencies, and federal grants. These include NIH funded Shared Instrumentation Grant S10OD017985, S10RR025141, and S10OD025092; CTSA grants UL1TR002243, UL1TR000445, and UL1RR024975. Genomic data are also supported by investigator-led projects that include U01HG004798, R01NS032830, RC2GM092618, P50GM115305, U01HG006378, U19HL065962, R01HD074711; and the additional funding sources listed at https://victr.vumc.org/biovu-funding/.

**AGENT (Duke)**

Duke's CATHGEN cohort was supported by National Heart, Lung, and Blood Institute grant R01-HL095987.

**AGENT (Intermountain)**

AGENT (Intermountain) is supported by internal funding and the Dell Loy Hansen Heart Foundation.

**AGENT (Western Ontario)**

The UWO Genetic Investigation of Supraventricular Tachycardias study acknowledges support from the Marianne Barrie Philanthropic Fund, the Canadian Institutes of Health Research, and the Cardiac Arrhythmic Network of Canada (CANet).

**AGENT (Wisconsin)**

AGENT (Wisconsin) is supported by NHLBI 1R01HL139738-01A1 (LLE) and 1R01HL128598-01 (LLE).

**CHS**

This research was supported by contracts HHSN268201200036C, HHSN268200800007C, HHSN268201800001C, N01HC55222, N01HC85079, N01HC85080, N01HC85081, N01HC85082, N01HC85083, N01HC85086, 75N92021D00006, and grants R01HL105756, U01HL080295 and U01HL130114 from the National Heart, Lung, and Blood Institute (NHLBI), with additional contribution from the National Institute of Neurological Disorders and Stroke (NINDS). Additional support was provided by R01AG023629 from the National Institute on Aging (NIA). A full list of principal CHS investigators and institutions can be found at CHS-NHLBI.org. The provision of genotyping data was supported in part by the National Center for Advancing Translational Sciences, CTSI grant UL1TR001881, and the National Institute of

Diabetes and Digestive and Kidney Disease Diabetes Research Center (DRC) grant DK063491 to the Southern California Diabetes Endocrinology Research Center. The content is solely the responsibility of the authors and does not necessarily represent the official views of the National Institutes of Health.

**FinnGen**

The FinnGen project is funded by two grants from Business Finland (HUS 4685/31/2016 and UH 4386/31/2016) and the following industry partners: AbbVie Inc., AstraZeneca UK Ltd, Biogen MA Inc., Bristol Myers Squibb (and Celgene Corporation & Celgene International II Sàrl), Genentech Inc., Merck Sharp & Dohme LCC, Pfizer Inc., GlaxoSmithKline Intellectual Property Development Ltd., Sanofi US Services Inc., Maze Therapeutics Inc., Janssen Biotech Inc, Novartis AG, and Boehringer Ingelheim International GmbH.

**FHS**

The Framingham Heart Study was supported by 75N92019D00031.

**Geisinger MyCode**

MyCode/DiscovEHR was supported by Geisinger and the Regeneron Genetics Center. The authors also wish to acknowledge assistance from Dr. Eric Carruth in compiling patient demographic data.

**MVP**

The MVP research is based on data from the Million Veteran Program, Office of Research and Development, Veterans Health Administration, and was supported by award # I01-BX004821. This publication does not represent the views of the Department of Veteran Affairs or the United States Government.

**Supplemental Figure 1.** Miami plot for sinus node dysfunction

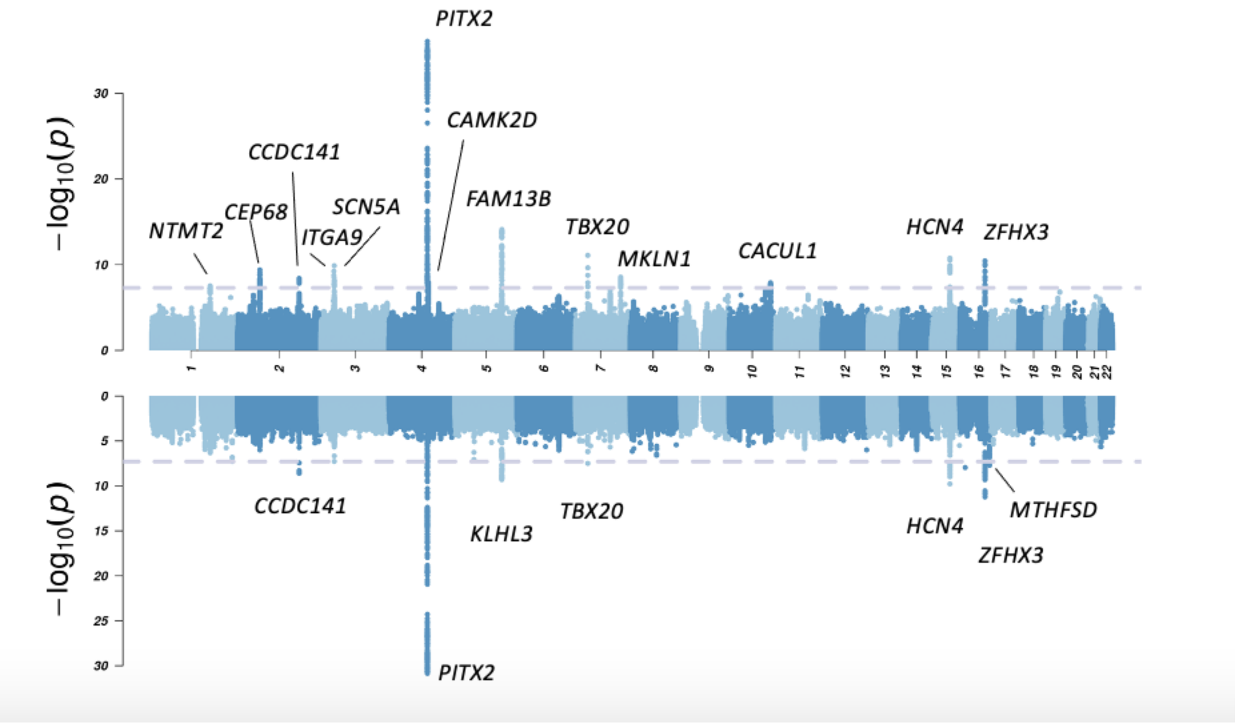

P-value is presented in -log10 scale for each association test between SNP and sinus node dysfunction (upper) or sinus node dysfunction – restrictive (lower) from fixed effect meta-analysis of multi-ancestry individuals. The grey dashed line indicated genome-wide significance at p-value equaled to $5x10^{-8}$.

**Supplemental Figure 2.** Miami plot for distal conduction disease

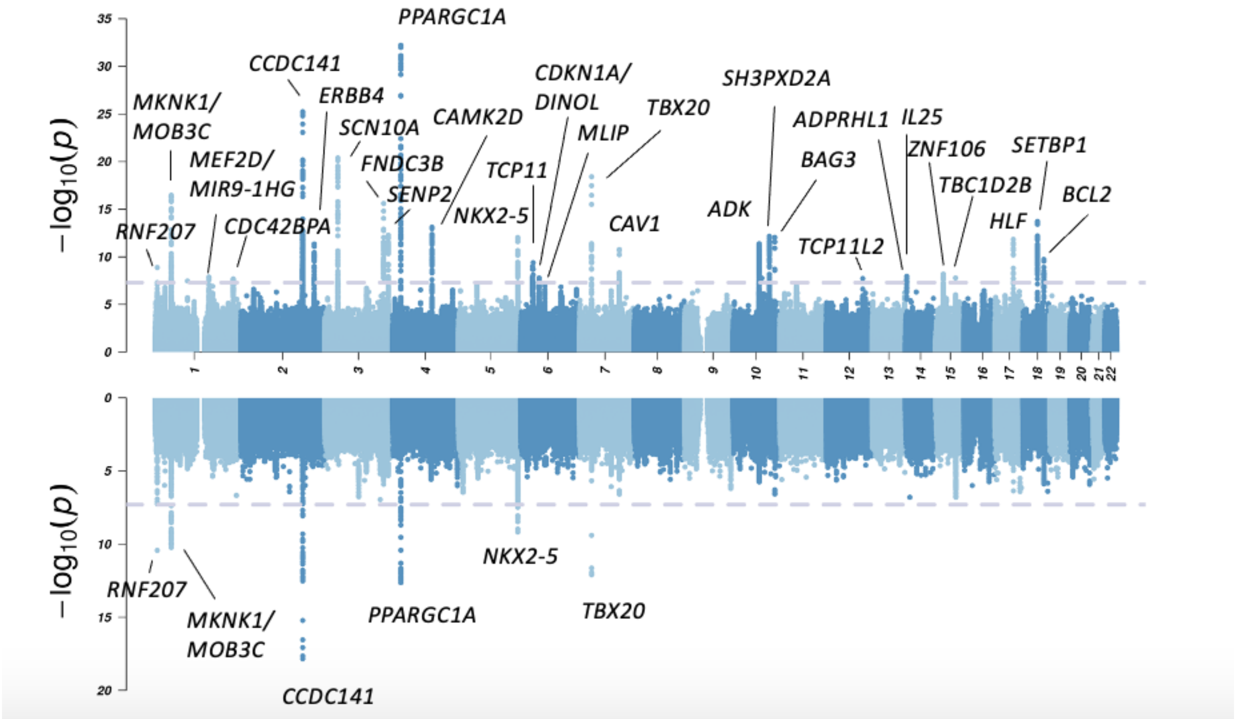

P-value is presented in -log10 scale for each association test between SNP and distal conduction disease (upper) or distal conduction disease – restrictive (lower) from fixed effect meta-analysis of multi-ancestry individuals. The grey dashed line indicated genome-wide significance at p-value equaled to $5x10^{-8}$.

**Supplemental Figure 3.** Q-Q plot for bradyarrhythmias

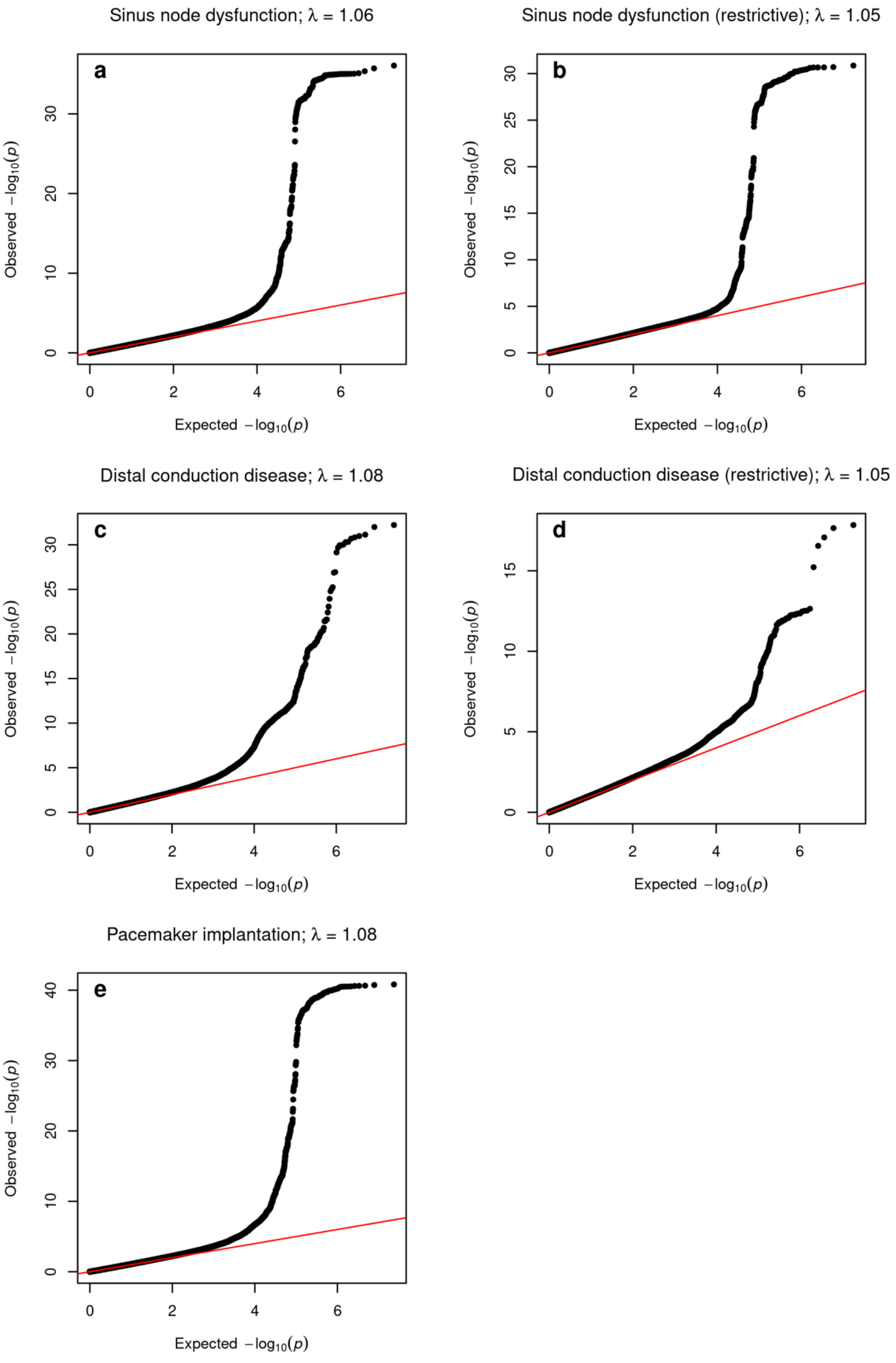

**Supplemental Figure 4.** Euler Diagram of genetic loci associated with bradyarrhythmia subtypes and pacemaker implantation

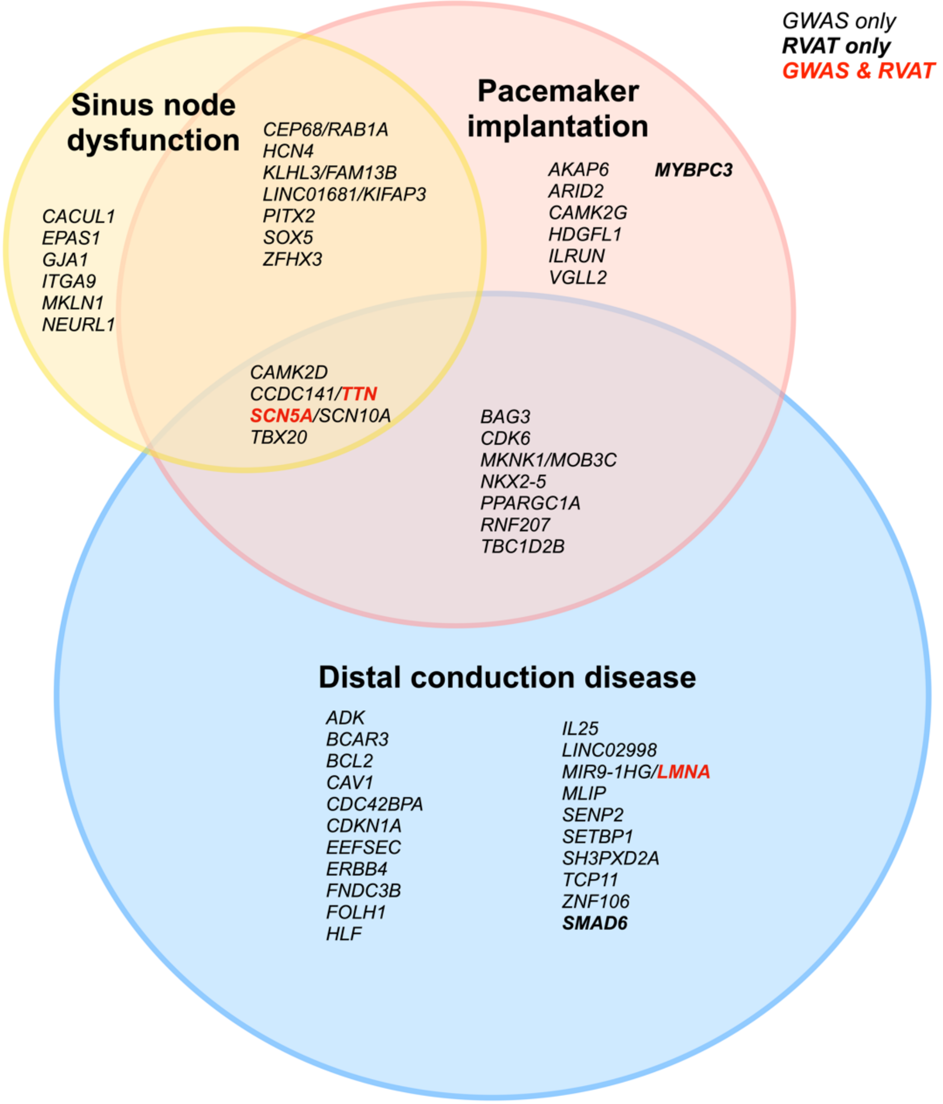

Figure shows the overlapped gene regions reported for bradyarrhythmias. We determined independent GWAS signals for bradyarrhythmias based on the distance-based approach (±1 Mb). We removed loci if they are within a peak region of other top loci or without at least 1 additional supportive variant nearby ($p<1x10^{-6}$). For the association signals close to or overlapping with more than one gene, we labeled both genes to avoid underestimating the shared genetic components across multiple Bradyarrhythmia traits. Genes which were identified in both GWAS and rare variant association tests (RVAT) are shown in bold red font. Genes which were only identified with RVAT are shown with bold black font.

**Supplemental Figure 5.** QQ plots of rare variant analysis

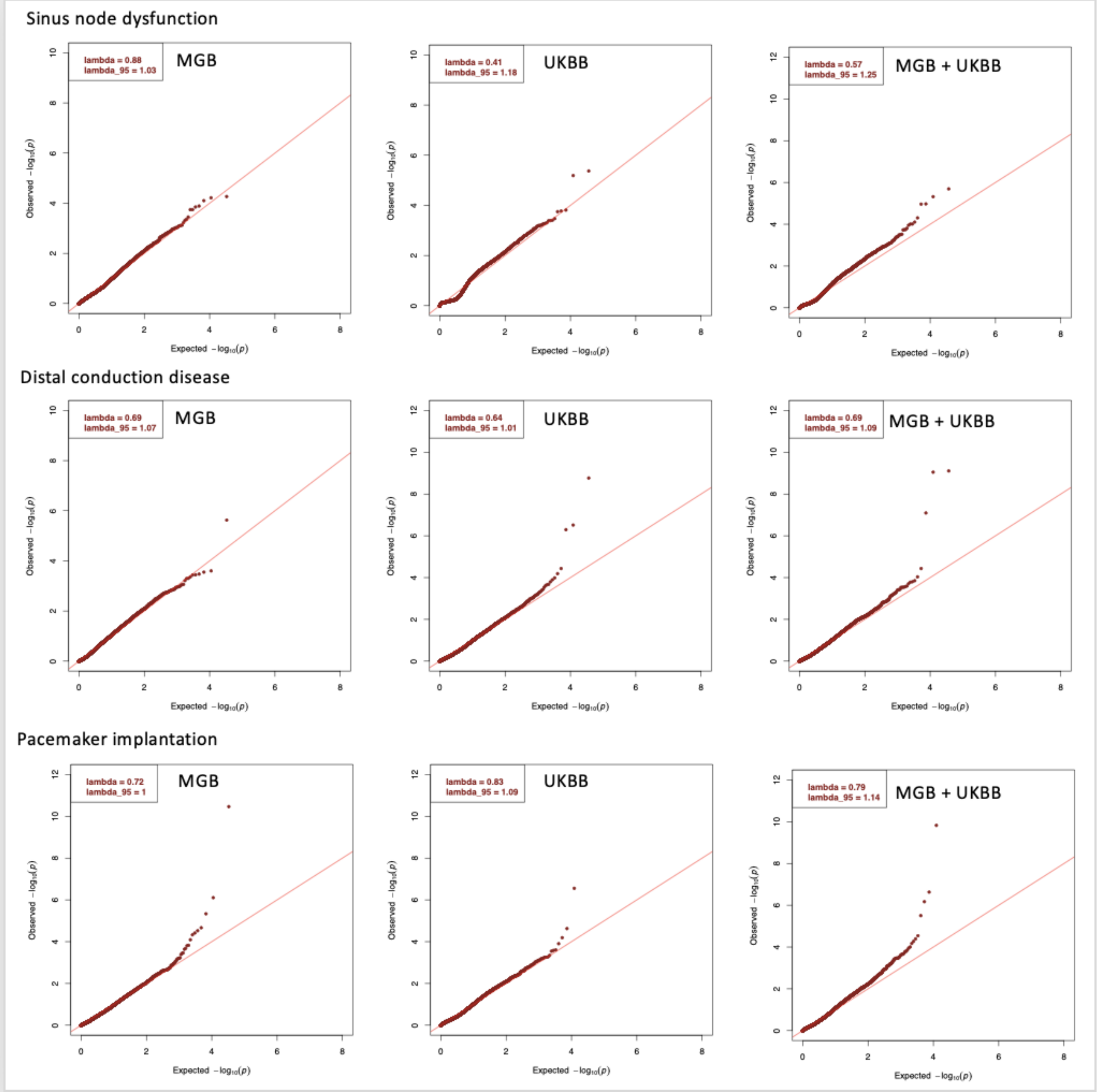

**Supplemental Figure 6.** Cauchy p-values of individual variation classes in exome-wide significant genes

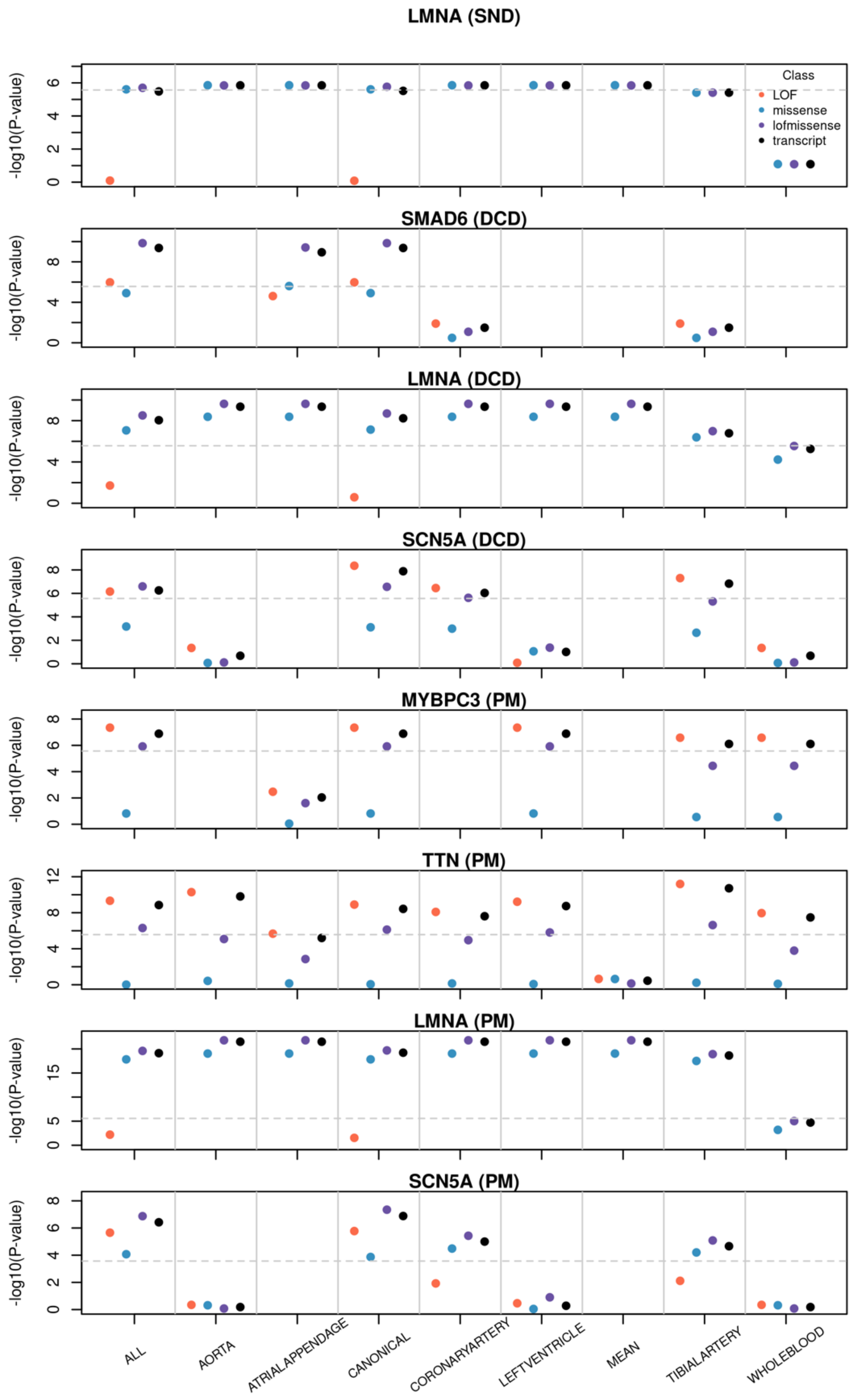

**Supplemental Figure 7.** Effect size of each association tests in exome-wide significant genes from various frequency and annotation filters in 9 categories

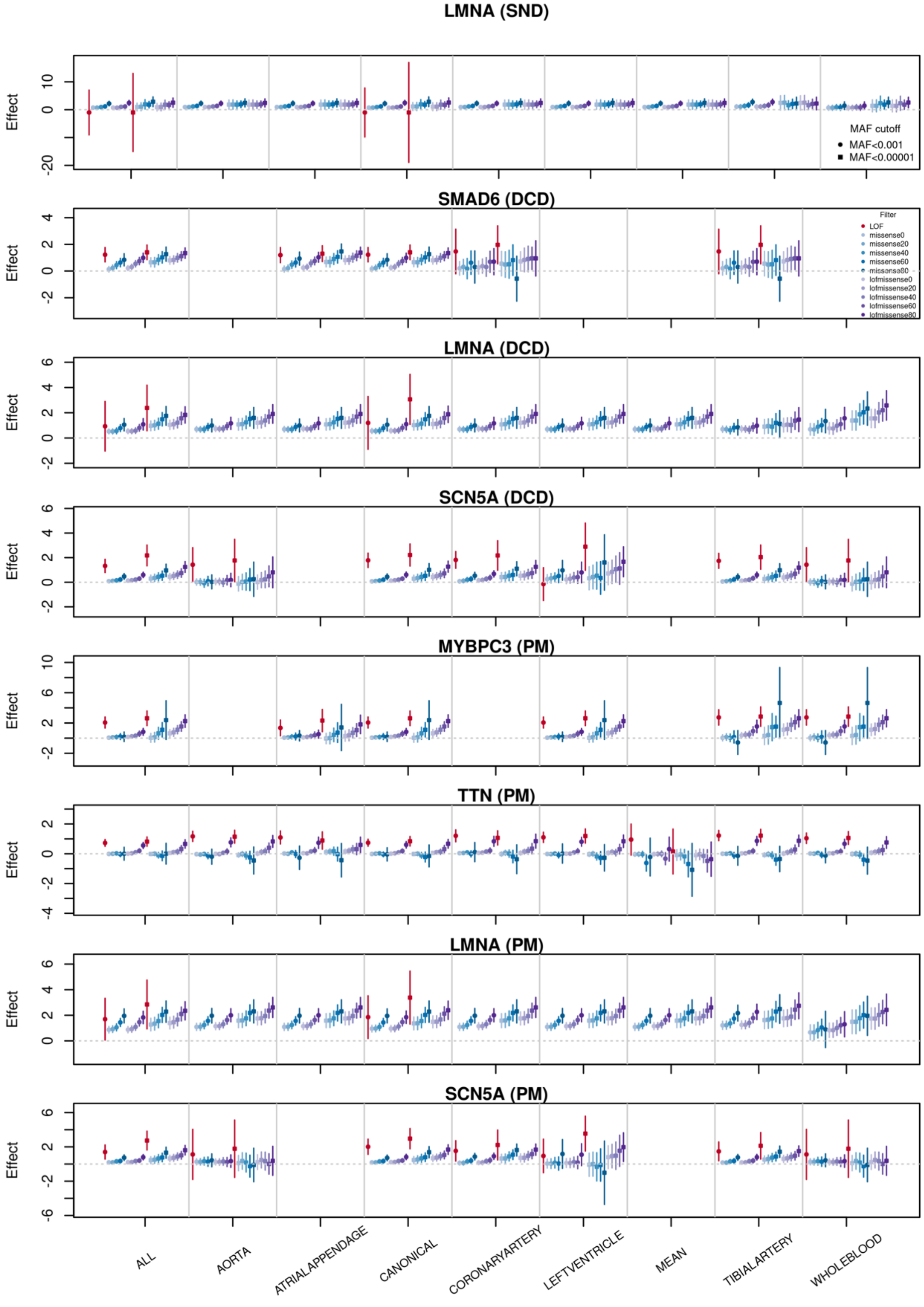

**Supplemental Figure 8** Pacemaker implantations in carriers of loss-of-function variants among 327,707 unrelated UK Biobank participants

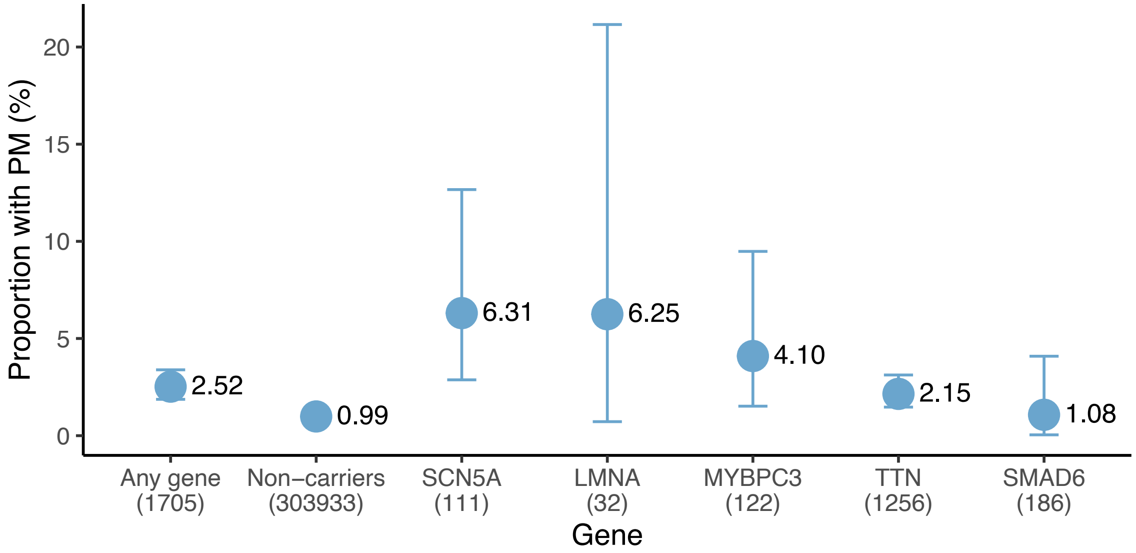

**Extended Data Figure 1.** Heritability enrichment for bradyarrhythmias in 9 major cell types from human heart

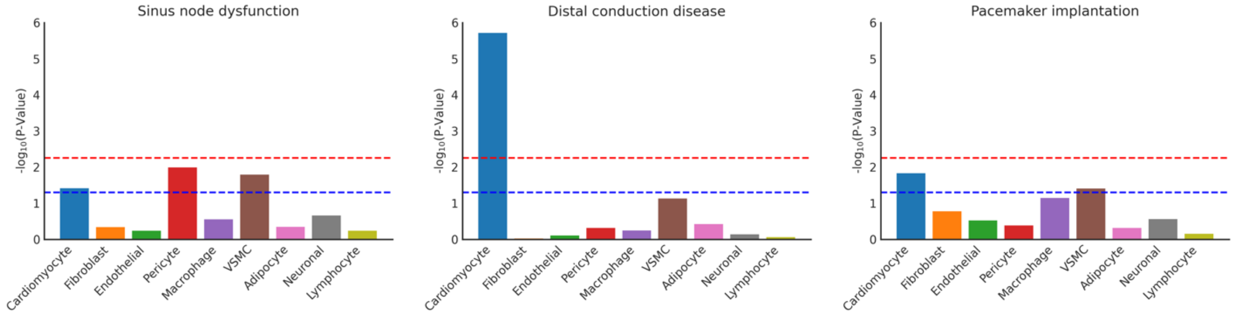

Results of stratified LD-Score regression (s-LDSC) on the combined major cell types in heart, where the p-values were derived from tests comparing GWAS heritability near cell type-specific genes to the background of all genes l. Dashed lines show statistical (red; with Bonferroni correction) and nominal (blue; p-value = 0.05) significant p-value thresholds.

**Extended Data Figure 2** The layered Cauchy combination pipeline in rare variant studies of bradyarrhythmia, as applied for each gene.

**a**

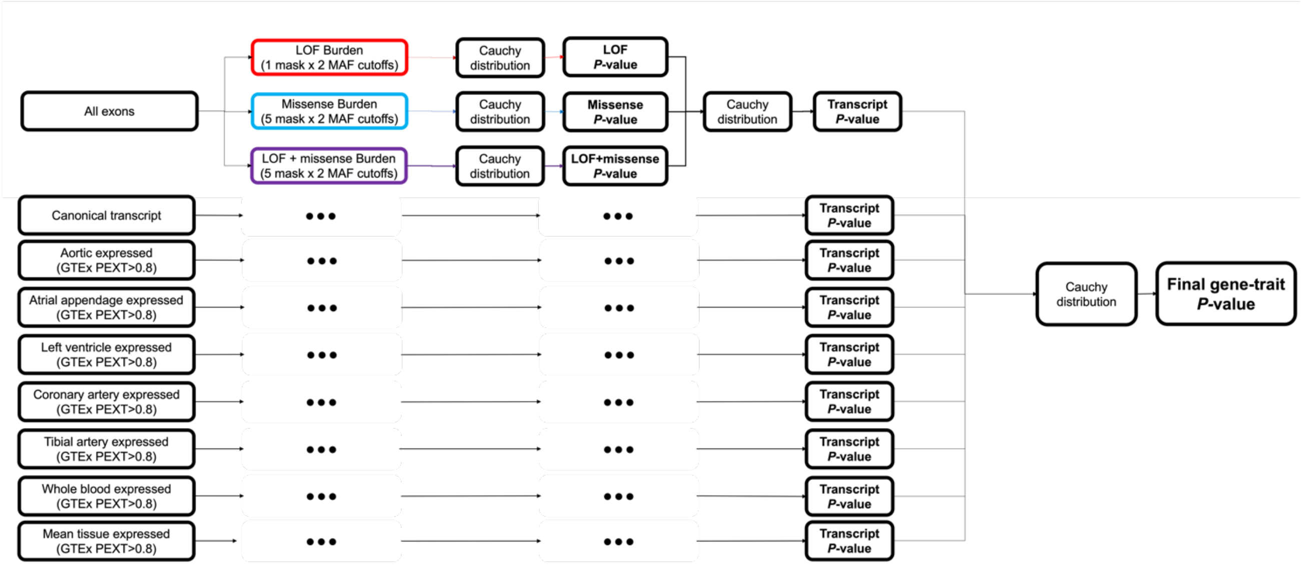

**b**

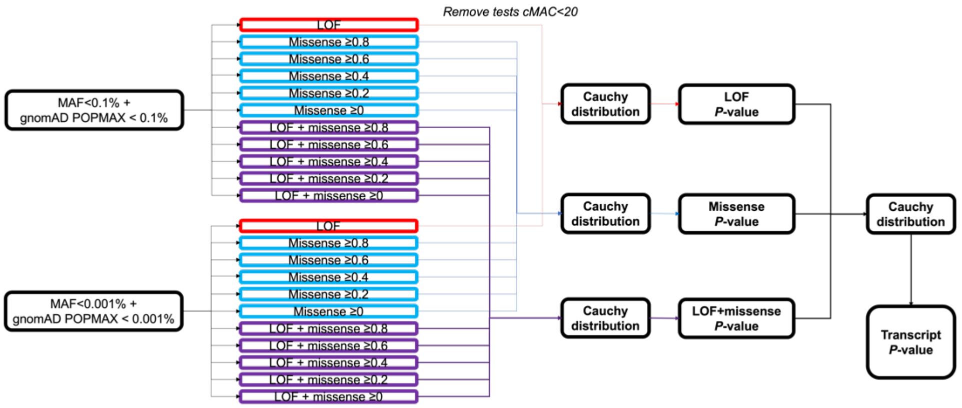

Panel **a** shows the birds-eye view of the pipeline as applied to each transcript of a given gene. As shown on the far left, we defined 9 'transcripts' that were defined as 1) all exons, 2) canonical gene transcripts as determined by Ensemble, 3) aortic expressed transcripts, formed by variants with aortic-specific pext values >=0.8, 4) atrial appendage-expressed transcripts, 5) left ventricle-expressed transcripts, 6) coronary artery-expressed transcripts, 7) tibial artery-expressed transcripts, 8) whole blood-expressed transcripts, and 9) mean transcript expression across GTEx tissues. For each of the defined gene transcripts, burden testing *P*-values based on LOF variant masks were combined into a single *P*-value using the Cauchy distribution test;[15] burden testing *P*-values based on various missense masks were combined into a single *P*-value using the Cauchy distribution test; and all LOF+missense burden *P*-values were combined into a single *P*-value using the Cauchy distribution test. Then the LOF, missense, and LOF+missense *P*-values were combined into a single transcript *P*-value using the Cauchy distribution test. This approach is repeated across the different transcripts, after which the various transcript *P*-values are finally combined into a single *P*-value for the gene-phenotype association using the Cauchy distribution test. Panel **b** shows, in more detail, the various frequency and annotation filters used within the pipeline for a given transcript. Mainly, two frequency filters are applied (MAF<0.1% and MAF<0.001%), and 11 different annotation filters are used (1 annotation for LOF variants; 5 for missense variants). The cutoffs for missense variants are based on the proportion of bio-informatic tools that predict a deleterious effect for the given missense variant.

**Reference**

1. Weng, L.-C. *et al.* Multi-Ancestry Genome-Wide Association Study Reveals Genetic Mechanisms of Supraventricular Arrhythmias. *Circulation: Genomic and Precision Medicine*, (under review).
2. Chang, C.C. *et al.* Second-generation PLINK: rising to the challenge of larger and richer datasets. *Gigascience* **4**, 7 (2015).
3. Bycroft, C. *et al.* The UK Biobank resource with deep phenotyping and genomic data. *Nature* **562**, 203-209 (2018).
4. Sudlow, C. *et al.* UK biobank: an open access resource for identifying the causes of a wide range of complex diseases of middle and old age. *PLoS Med* **12**, e1001779 (2015).
5. Backman, J.D. *et al.* Exome sequencing and analysis of 454,787 UK Biobank participants. *Nature* **599**, 628-634 (2021).
6. Szustakowski, J.D. *et al.* Advancing human genetics research and drug discovery through exome sequencing of the UK Biobank. *Nat Genet* **53**, 942–948 (2021).
7. Jurgens, S.J. *et al.* Analysis of rare genetic variation underlying cardiometabolic diseases and traits among 200,000 individuals in the UK Biobank. *Nat Genet* **54**, 240-250 (2022).
8. Karlson, E.W., Boutin, N.T., Hoffnagle, A.G. & Allen, N.L. Building the Partners HealthCare Biobank at Partners Personalized Medicine: Informed Consent, Return of Research Results, Recruitment Lessons and Operational Considerations. *J Pers Med* **6**(2016).
9. Boutin, N.T. *et al.* Implementation of Electronic Consent at a Biobank: An Opportunity for Precision Medicine Research. *J Pers Med* **6**(2016).
10. Liu, X., Wu, C., Li, C. & Boerwinkle, E. dbNSFP v3.0: A One-Stop Database of Functional Predictions and Annotations for Human Nonsynonymous and Splice-Site SNVs. *Hum Mutat* **37**, 235-41 (2016).
11. Karczewski, K.J. *et al.* The mutational constraint spectrum quantified from variation in 141,456 humans. *Nature* **581**, 434-443 (2020).
12. McLaren, W. *et al.* The Ensembl Variant Effect Predictor. *Genome Biol* **17**, 122 (2016).
13. Cummings, B.B. *et al.* Transcript expression-aware annotation improves rare variant interpretation. *Nature* **581**, 452-458 (2020).
14. GTEx Consortium. The GTEx Consortium atlas of genetic regulatory effects across human tissues. *Science* **369**, 1318-1330 (2020).
15. Liu, Y. *et al.* ACAT: A Fast and Powerful p Value Combination Method for Rare-Variant Analysis in Sequencing Studies. *Am J Hum Genet* **104**, 410-421 (2019).

### BANNER AUTHOR

#### Study-specific Banner Authors

#### FinnGen

| Full Name | Affiliation | Role 1 | Role 2 |
|---|---|---|---|
| Aarno Palotie | Institute for Molecular Medicine Finland (FIMM), HiLIFE, University of Helsinki, Helsinki, Finland; Broad Institute of MIT and Harvard; Massachusetts General Hospital | **Steering Committee** | **Steering Committee** |
| Mark Daly | Institute for Molecular Medicine Finland (FIMM), HiLIFE, University of Helsinki, Helsinki, Finland; Broad Institute of MIT and Harvard; Massachusetts General Hospital | **Steering Committee** | **Steering Committee** |
| Bridget Riley-Gills | Abbvie, Chicago, IL, United States | **Steering Committee** | **Pharmaceutical companies** |
| Howard Jacob | Abbvie, Chicago, IL, United States | **Steering Committee** | **Pharmaceutical companies** |
| Dirk Paul | Astra Zeneca, Cambridge, United Kingdom | **Steering Committee** | **Pharmaceutical companies** |
| Athena Matakidou | Astra Zeneca, Cambridge, United Kingdom | **Steering Committee** | **Pharmaceutical companies** |
| Adam Platt | Astra Zeneca, Cambridge, United Kingdom | **Steering Committee** | **Pharmaceutical companies** |
| Heiko Runz | Biogen, Cambridge, MA, United States | **Steering Committee** | **Pharmaceutical companies** |
| Sally John | Biogen, Cambridge, MA, United States | **Steering Committee** | **Pharmaceutical companies** |
| George Okafo | Boehringer Ingelheim, Ingelheim am Rhein, Germany | **Steering Committee** | **Pharmaceutical companies** |
| Nathan Lawless | Boehringer Ingelheim, Ingelheim am Rhein, Germany | **Steering Committee** | **Pharmaceutical companies** |

| | | | |
|---|---|---|---|
| Heli Salminen-Mankonen | Boehringer Ingelheim, Ingelheim am Rhein, Germany | **Steering Committee** | **Pharmaceutical companies** |
| Robert Plenge | Bristol Myers Squibb, New York, NY, United States | **Steering Committee** | **Pharmaceutical companies** |
| Joseph Maranville | Bristol Myers Squibb, New York, NY, United States | **Steering Committee** | **Pharmaceutical companies** |
| Mark McCarthy | Genentech, San Francisco, CA, United States | **Steering Committee** | **Pharmaceutical companies** |
| Julie Hunkapiller | Genentech, San Francisco, CA, United States | **Steering Committee** | **Pharmaceutical companies** |
| Margaret G. Ehm | GlaxoSmithKline, Collegeville, PA, United States | **Steering Committee** | **Pharmaceutical companies** |
| Kirsi Auro | GlaxoSmithKline, Espoo, Finland | **Steering Committee** | **Pharmaceutical companies** |
| Simonne Longerich | Merck, Kenilworth, NJ, United States | **Steering Committee** | **Pharmaceutical companies** |
| Caroline Fox | Merck, Kenilworth, NJ, United States | **Steering Committee** | **Pharmaceutical companies** |
| Anders Mälarstig | Pfizer, New York, NY, United States | **Steering Committee** | **Pharmaceutical companies** |
| Katherine Klinger | Translational Sciences, Sanofi R&D, Framingham, MA, USA | **Steering Committee** | **Pharmaceutical companies** |
| Deepak Raipal | Translational Sciences, Sanofi R&D, Framingham, MA, USA | **Steering Committee** | **Pharmaceutical companies** |
| Eric Green | Maze Therapeutics, San Francisco, CA, United States | **Steering Committee** | **Pharmaceutical companies** |
| Robert Graham | Maze Therapeutics, San Francisco, CA, United States | **Steering Committee** | **Pharmaceutical companies** |
| Robert Yang | Janssen Biotech, Beerse, Belgium | **Steering Committee** | **Pharmaceutical companies** |
| Chris O´Donnell | Novartis Institutes for BioMedical Research, Cambridge, MA, United States | **Steering Committee** | **Pharmaceutical companies** |

| | | | |
|---|---|---|---|
| Tomi P. Mäkelä | HiLIFE, University of Helsinki, Finland, Finland | **Steering Committee** | **University of Helsinki & Biobanks** |
| Jaakko Kaprio | Institute for Molecular Medicine Finland (FIMM), HiLIFE, University of Helsinki, Helsinki, Finland | **Steering Committee** | **University of Helsinki & Biobanks** |
| Petri Virolainen | Auria Biobank / University of Turku / Hospital District of Southwest Finland, Turku, Finland | **Steering Committee** | **University of Helsinki & Biobanks** |
| Antti Hakanen | Auria Biobank / University of Turku / Hospital District of Southwest Finland, Turku, Finland | **Steering Committee** | **University of Helsinki & Biobanks** |
| Terhi Kilpi | THL Biobank / Finnish Institute for Health and Welfare (THL), Helsinki, Finland | **Steering Committee** | **University of Helsinki & Biobanks** |
| Markus Perola | THL Biobank / Finnish Institute for Health and Welfare (THL), Helsinki, Finland | **Steering Committee** | **University of Helsinki & Biobanks** |
| Jukka Partanen | Finnish Red Cross Blood Service / Finnish Hematology Registry and Clinical Biobank, Helsinki, Finland | **Steering Committee** | **University of Helsinki & Biobanks** |
| Anne Pitkäranta | Helsinki Biobank / Helsinki University and Hospital District of Helsinki and Uusimaa, Helsinki | **Steering Committee** | **University of Helsinki & Biobanks** |
| Taneli Raivio | Helsinki Biobank / Helsinki University and Hospital District of Helsinki and Uusimaa, Helsinki | **Steering Committee** | **University of Helsinki & Biobanks** |
| Raisa Serpi | Northern Finland Biobank Borealis / University of Oulu / Northern Ostrobothnia Hospital District, Oulu, Finland | **Steering Committee** | **University of Helsinki & Biobanks** |
| Tarja Laitinen | Finnish Clinical Biobank Tampere / University of Tampere / Pirkanmaa Hospital District, Tampere, Finland | **Steering Committee** | **University of Helsinki & Biobanks** |
| Veli-Matti Kosma | Biobank of Eastern Finland / University of Eastern Finland / Northern Savo Hospital District, Kuopio, Finland | **Steering Committee** | **University of Helsinki & Biobanks** |

| | | | |
|---|---|---|---|
| Jari Laukkanen | Central Finland Biobank / University of Jyväskylä / Central Finland Health Care District, Jyväskylä, Finland | **Steering Committee** | **University of Helsinki & Biobanks** |
| Marco Hautalahti | FINBB - Finnish biobank cooperative | **Steering Committee** | **University of Helsinki & Biobanks** |
| Outi Tuovila | Business Finland, Helsinki, Finland | **Steering Committee** | **Other Experts/ Non-Voting Members** |
| Raimo Pakkanen | Business Finland, Helsinki, Finland | **Steering Committee** | **Other Experts/ Non-Voting Members** |
| Jeffrey Waring | Abbvie, Chicago, IL, United States | **Scientific Committee** | **Pharmaceutical companies** |
| Bridget Riley-Gillis | Abbvie, Chicago, IL, United States | **Scientific Committee** | **Pharmaceutical companies** |
| Fedik Rahimov | Abbvie, Chicago, IL, United States | **Scientific Committee** | **Pharmaceutical companies** |
| Ioanna Tachmazidou | Astra Zeneca, Cambridge, United Kingdom | **Scientific Committee** | **Pharmaceutical companies** |
| Chia-Yen Chen | Biogen, Cambridge, MA, United States | **Scientific Committee** | **Pharmaceutical companies** |
| Heiko Runz | Biogen, Cambridge, MA, United States | **Scientific Committee** | **Pharmaceutical companies** |
| Zhihao Ding | Boehringer Ingelheim, Ingelheim am Rhein, Germany | **Scientific Committee** | **Pharmaceutical companies** |
| Marc Jung | Boehringer Ingelheim, Ingelheim am Rhein, Germany | **Scientific Committee** | **Pharmaceutical companies** |
| Shameek Biswas | Bristol Myers Squibb, New York, NY, United States | **Scientific Committee** | **Pharmaceutical companies** |
| Rion Pendergrass | Genentech, San Francisco, CA, United States | **Scientific Committee** | **Pharmaceutical companies** |
| Julie Hunkapiller | Genentech, San Francisco, CA, United States | **Scientific Committee** | **Pharmaceutical companies** |
| Margaret G. Ehm | GlaxoSmithKline, Collegeville, PA, United States | **Scientific Committee** | **Pharmaceutical companies** |

| | | | |
|---|---|---|---|
| David Pulford | GlaxoSmithKline, Stevenage, United Kingdom | **Scientific Committee** | **Pharmaceutical companies** |
| Neha Raghavan | Merck, Kenilworth, NJ, United States | **Scientific Committee** | **Pharmaceutical companies** |
| Adriana Huertas-Vazquez | Merck, Kenilworth, NJ, United States | **Scientific Committee** | **Pharmaceutical companies** |
| Jae-Hoon Sul | Merck, Kenilworth, NJ, United States | **Scientific Committee** | **Pharmaceutical companies** |
| Anders Mälarstig | Pfizer, New York, NY, United States | **Scientific Committee** | **Pharmaceutical companies** |
| Xinli Hu | Pfizer, New York, NY, United States | **Scientific Committee** | **Pharmaceutical companies** |
| Katherine Klinger | Translational Sciences, Sanofi R&D, Framingham, MA, USA | **Scientific Committee** | **Pharmaceutical companies** |
| Robert Graham | Maze Therapeutics, San Francisco, CA, United States | **Scientific Committee** | **Pharmaceutical companies** |
| Eric Green | Maze Therapeutics, San Francisco, CA, United States | **Scientific Committee** | **Pharmaceutical companies** |
| Sahar Mozaffari | Maze Therapeutics, San Francisco, CA, United States | **Scientific Committee** | **Pharmaceutical companies** |
| Dawn Waterworth | Janssen Research & Development, LLC, Spring House, PA, United States | **Scientific Committee** | **Pharmaceutical companies** |
| Nicole Renaud | Novartis Institutes for BioMedical Research, Cambridge, MA, United States | **Scientific Committee** | **Pharmaceutical companies** |
| Ma´en Obeidat | Novartis Institutes for BioMedical Research, Cambridge, MA, United States | **Scientific Committee** | **Pharmaceutical companies** |
| Samuli Ripatti | Institute for Molecular Medicine Finland (FIMM), HiLIFE, University of Helsinki, Helsinki, Finland | **Scientific Committee** | **University of Helsinki & Biobanks** |
| Johanna Schleutker | Auria Biobank / Univ. of Turku / Hospital District of Southwest Finland, Turku, Finland | **Scientific Committee** | **University of Helsinki & Biobanks** |
| Markus Perola | THL Biobank / Finnish Institute for Health and Welfare (THL), Helsinki, Finland | **Scientific Committee** | **University of Helsinki & Biobanks** |

| | | | |
|---|---|---|---|
| Mikko Arvas | Finnish Red Cross Blood Service / Finnish Hematology Registry and Clinical Biobank, Helsinki, Finland | **Scientific Committee** | **University of Helsinki & Biobanks** |
| Olli Carpén | Helsinki Biobank / Helsinki University and Hospital District of Helsinki and Uusimaa, Helsinki | **Scientific Committee** | **University of Helsinki & Biobanks** |
| Reetta Hinttala | Northern Finland Biobank Borealis / University of Oulu / Northern Ostrobothnia Hospital District, Oulu, Finland | **Scientific Committee** | **University of Helsinki & Biobanks** |
| Johannes Kettunen | Northern Finland Biobank Borealis / University of Oulu / Northern Ostrobothnia Hospital District, Oulu, Finland | **Scientific Committee** | **University of Helsinki & Biobanks** |
| Arto Mannermaa | Biobank of Eastern Finland / University of Eastern Finland / Northern Savo Hospital District, Kuopio, Finland | **Scientific Committee** | **University of Helsinki & Biobanks** |
| Katriina Aalto-Setälä | Faculty of Medicine and Health Technology, Tampere University, Tampere, Finland | **Scientific Committee** | **University of Helsinki & Biobanks** |
| Mika Kähönen | Finnish Clinical Biobank Tampere / University of Tampere / Pirkanmaa Hospital District, Tampere, Finland | **Scientific Committee** | **University of Helsinki & Biobanks** |
| Jari Laukkanen | Central Finland Biobank / University of Jyväskylä / Central Finland Health Care District, Jyväskylä, Finland | **Scientific Committee** | **University of Helsinki & Biobanks** |
| Johanna Mäkelä | FINBB - Finnish biobank cooperative | **Scientific Committee** | **University of Helsinki & Biobanks** |
| Reetta Kälviäinen | Northern Savo Hospital District, Kuopio, Finland | **Clinical Groups** | **Neurology Group** |
| Valtteri Julkunen | Northern Savo Hospital District, Kuopio, Finland | **Clinical Groups** | **Neurology Group** |
| Hilkka Soininen | Northern Savo Hospital District, Kuopio, Finland | **Clinical Groups** | **Neurology Group** |
| Anne Remes | Northern Ostrobothnia Hospital District, Oulu, Finland | **Clinical Groups** | **Neurology Group** |

| | | | |
|---|---|---|---|
| Mikko Hiltunen | University of Eastern Finland, Kuopio, Finland | **Clinical Groups** | **Neurology Group** |
| Jukka Peltola | Pirkanmaa Hospital District, Tampere, Finland | **Clinical Groups** | **Neurology Group** |
| Minna Raivio | Hospital District of Helsinki and Uusimaa, Helsinki, Finland | **Clinical Groups** | **Neurology Group** |
| Pentti Tienari | Hospital District of Helsinki and Uusimaa, Helsinki, Finland | **Clinical Groups** | **Neurology Group** |
| Juha Rinne | Hospital District of Southwest Finland, Turku, Finland | **Clinical Groups** | **Neurology Group** |
| Roosa Kallionpää | Hospital District of Southwest Finland, Turku, Finland | **Clinical Groups** | **Neurology Group** |
| Juulia Partanen | Institute for Molecular Medicine Finland, HiLIFE, University of Helsinki, Finland | **Clinical Groups** | **Neurology Group** |
| Ali Abbasi | Abbvie, Chicago, IL, United States | **Clinical Groups** | **Neurology Group** |
| Adam Ziemann | Abbvie, Chicago, IL, United States | **Clinical Groups** | **Neurology Group** |
| Nizar Smaoui | Abbvie, Chicago, IL, United States | **Clinical Groups** | **Neurology Group** |
| Anne Lehtonen | Abbvie, Chicago, IL, United States | **Clinical Groups** | **Neurology Group** |
| Susan Eaton | Biogen, Cambridge, MA, United States | **Clinical Groups** | **Neurology Group** |
| Heiko Runz | Biogen, Cambridge, MA, United States | **Clinical Groups** | **Neurology Group** |
| Sanni Lahdenperä | Biogen, Cambridge, MA, United States | **Clinical Groups** | **Neurology Group** |
| Shameek Biswas | Bristol Myers Squibb, New York, NY, United States | **Clinical Groups** | **Neurology Group** |
| Julie Hunkapiller | Genentech, San Francisco, CA, United States | **Clinical Groups** | **Neurology Group** |

| | | | |
|---|---|---|---|
| Natalie Bowers | Genentech, San Francisco, CA, United States | **Clinical Groups** | **Neurology Group** |
| Edmond Teng | Genentech, San Francisco, CA, United States | **Clinical Groups** | **Neurology Group** |
| Rion Pendergrass | Genentech, San Francisco, CA, United States | **Clinical Groups** | **Neurology Group** |
| Fanli Xu | GlaxoSmithKline, Brentford, United Kingdom | **Clinical Groups** | **Neurology Group** |
| David Pulford | GlaxoSmithKline, Stevenage, United Kingdom | **Clinical Groups** | **Neurology Group** |
| Kirsi Auro | GlaxoSmithKline, Espoo, Finland | **Clinical Groups** | **Neurology Group** |
| Laura Addis | GlaxoSmithKline, Brentford, United Kingdom | **Clinical Groups** | **Neurology Group** |
| John Eicher | GlaxoSmithKline, Brentford, United Kingdom | **Clinical Groups** | **Neurology Group** |
| Qingqin S Li | Janssen Research & Development, LLC, Titusville, NJ 08560, United States | **Clinical Groups** | **Neurology Group** |
| Karen He | Janssen Research & Development, LLC, Spring House, PA, United States | **Clinical Groups** | **Neurology Group** |
| Ekaterina Khramtsova | Janssen Research & Development, LLC, Spring House, PA, United States | **Clinical Groups** | **Neurology Group** |
| Neha Raghavan | Merck, Kenilworth, NJ, United States | **Clinical Groups** | **Neurology Group** |
| Martti Färkkilä | Hospital District of Helsinki and Uusimaa, Helsinki, Finland | **Clinical Groups** | **Gastroenterology Group** |
| Jukka Koskela | Hospital District of Helsinki and Uusimaa, Helsinki, Finland | **Clinical Groups** | **Gastroenterology Group** |
| Sampsa Pikkarainen | Hospital District of Helsinki and Uusimaa, Helsinki, Finland | **Clinical Groups** | **Gastroenterology Group** |
| Airi Jussila | Pirkanmaa Hospital District, Tampere, Finland | **Clinical Groups** | **Gastroenterology Group** |

| | | | |
|---|---|---|---|
| Katri Kaukinen | Pirkanmaa Hospital District, Tampere, Finland | **Clinical Groups** | **Gastroenterology Group** |
| Timo Blomster | Northern Ostrobothnia Hospital District, Oulu, Finland | **Clinical Groups** | **Gastroenterology Group** |
| Mikko Kiviniemi | Northern Savo Hospital District, Kuopio, Finland | **Clinical Groups** | **Gastroenterology Group** |
| Markku Voutilainen | Hospital District of Southwest Finland, Turku, Finland | **Clinical Groups** | **Gastroenterology Group** |
| Mark Daly | Institute for Molecular Medicine, Finland (FIMM), HiLIFE, University of Helsinki, Helsinki, Finland; Broad Institute of MIT and Harvard; Massachusetts General Hospital | **Clinical Groups** | **Gastroenterology Group** |
| Ali Abbasi | Abbvie, Chicago, IL, United States | **Clinical Groups** | **Gastroenterology Group** |
| Jeffrey Waring | Abbvie, Chicago, IL, United States | **Clinical Groups** | **Gastroenterology Group** |
| Nizar Smaoui | Abbvie, Chicago, IL, United States | **Clinical Groups** | **Gastroenterology Group** |
| Fedik Rahimov | Abbvie, Chicago, IL, United States | **Clinical Groups** | **Gastroenterology Group** |
| Anne Lehtonen | Abbvie, Chicago, IL, United States | **Clinical Groups** | **Gastroenterology Group** |
| Tim Lu | Genentech, San Francisco, CA, United States | **Clinical Groups** | **Gastroenterology Group** |
| Natalie Bowers | Genentech, San Francisco, CA, United States | **Clinical Groups** | **Gastroenterology Group** |
| Rion Pendergrass | Genentech, San Francisco, CA, United States | **Clinical Groups** | **Gastroenterology Group** |
| Linda McCarthy | GlaxoSmithKline, Brentford, United Kingdom | **Clinical Groups** | **Gastroenterology Group** |
| Amy Hart | Janssen Research & Development, LLC, Spring House, PA, United States | **Clinical Groups** | **Gastroenterology Group** |

| | | | |
|---|---|---|---|
| Meijian Guan | Janssen Research & Development, LLC, Spring House, PA, United States | **Clinical Groups** | **Gastroenterology Group** |
| Jason Miller | Merck, Kenilworth, NJ, United States | **Clinical Groups** | **Gastroenterology Group** |
| Kirsi Kalpala | Pfizer, New York, NY, United States | **Clinical Groups** | **Gastroenterology Group** |
| Melissa Miller | Pfizer, New York, NY, United States | **Clinical Groups** | **Gastroenterology Group** |
| Xinli Hu | Pfizer, New York, NY, United States | **Clinical Groups** | **Gastroenterology Group** |
| Kari Eklund | Hospital District of Helsinki and Uusimaa, Helsinki, Finland | **Clinical Groups** | **Rheumatology Group** |
| Antti Palomäki | Hospital District of Southwest Finland, Turku, Finland | **Clinical Groups** | **Rheumatology Group** |
| Pia Isomäki | Pirkanmaa Hospital District, Tampere, Finland | **Clinical Groups** | **Rheumatology Group** |
| Laura Pirilä | Hospital District of Southwest Finland, Turku, Finland | **Clinical Groups** | **Rheumatology Group** |
| Oili Kaipiainen-Seppänen | Northern Savo Hospital District, Kuopio, Finland | **Clinical Groups** | **Rheumatology Group** |
| Johanna Huhtakangas | Northern Ostrobothnia Hospital District, Oulu, Finland | **Clinical Groups** | **Rheumatology Group** |
| Nina Mars | Institute for Molecular Medicine Finland (FIMM), HiLIFE, University of Helsinki, Helsinki, Finland | **Clinical Groups** | **Rheumatology Group** |
| Ali Abbasi | Abbvie, Chicago, IL, United States | **Clinical Groups** | **Rheumatology Group** |
| Jeffrey Waring | Abbvie, Chicago, IL, United States | **Clinical Groups** | **Rheumatology Group** |
| Fedik Rahimov | Abbvie, Chicago, IL, United States | **Clinical Groups** | **Rheumatology Group** |
| Apinya Lertratanakul | Abbvie, Chicago, IL, United States | **Clinical Groups** | **Rheumatology Group** |

| | | | |
|---|---|---|---|
| Nizar Smaoui | Abbvie, Chicago, IL, United States | **Clinical Groups** | **Rheumatology Group** |
| Anne Lehtonen | Abbvie, Chicago, IL, United States | **Clinical Groups** | **Rheumatology Group** |
| Marla Hochfeld | Bristol Myers Squibb, New York, NY, United States | **Clinical Groups** | **Rheumatology Group** |
| Natalie Bowers | Genentech, San Francisco, CA, United States | **Clinical Groups** | **Rheumatology Group** |
| Rion Pendergrass | Genentech, San Francisco, CA, United States | **Clinical Groups** | **Rheumatology Group** |
| Jorge Esparza Gordillo | GlaxoSmithKline, Brentford, United Kingdom | **Clinical Groups** | **Rheumatology Group** |
| Kirsi Auro | GlaxoSmithKline, Espoo, Finland | **Clinical Groups** | **Rheumatology Group** |
| Dawn Waterworth | Janssen Research & Development, LLC, Spring House, PA, United States | **Clinical Groups** | **Rheumatology Group** |
| Fabiana Farias | Merck, Kenilworth, NJ, United States | **Clinical Groups** | **Rheumatology Group** |
| Kirsi Kalpala | Pfizer, New York, NY, United States | **Clinical Groups** | **Rheumatology Group** |
| Nan Bing | Pfizer, New York, NY, United States | **Clinical Groups** | **Rheumatology Group** |
| Xinli Hu | Pfizer, New York, NY, United States | **Clinical Groups** | **Rheumatology Group** |
| Tarja Laitinen | Pirkanmaa Hospital District, Tampere, Finland | **Clinical Groups** | **Pulmonology Group** |
| Margit Pelkonen | Northern Savo Hospital District, Kuopio, Finland | **Clinical Groups** | **Pulmonology Group** |
| Paula Kauppi | Hospital District of Helsinki and Uusimaa, Helsinki, Finland | **Clinical Groups** | **Pulmonology Group** |
| Hannu Kankaanranta | University of Gothenburg, Gothenburg, Sweden/ Seinäjoki Central Hospital, Seinäjoki, | **Clinical Groups** | **Pulmonology Group** |

| | | | |
|---|---|---|---|
| | Finland/ Tampere University, Tampere, Finland | | |
| Terttu Harju | Northern Ostrobothnia Hospital District, Oulu, Finland | **Clinical Groups** | **Pulmonology Group** |
| Riitta Lahesmaa | Hospital District of Southwest Finland, Turku, Finland | **Clinical Groups** | **Pulmonology Group** |
| Nizar Smaoui | Abbvie, Chicago, IL, United States | **Clinical Groups** | **Pulmonology Group** |
| Glenda Lassi | Astra Zeneca, Cambridge, United Kingdom | **Clinical Groups** | **Pulmonology Group** |
| Susan Eaton | Biogen, Cambridge, MA, United States | **Clinical Groups** | **Pulmonology Group** |
| Hubert Chen | Genentech, San Francisco, CA, United States | **Clinical Groups** | **Pulmonology Group** |
| Rion Pendergrass | Genentech, San Francisco, CA, United States | **Clinical Groups** | **Pulmonology Group** |
| Natalie Bowers | Genentech, San Francisco, CA, United States | **Clinical Groups** | **Pulmonology Group** |
| Joanna Betts | GlaxoSmithKline, Brentford, United Kingdom | **Clinical Groups** | **Pulmonology Group** |
| Kirsi Auro | GlaxoSmithKline, Espoo, Finland | **Clinical Groups** | **Pulmonology Group** |
| Rajashree Mishra | GlaxoSmithKline, Brentford, United Kingdom | **Clinical Groups** | **Pulmonology Group** |
| Majd Mouded | Novartis, Basel, Switzerland | **Clinical Groups** | **Pulmonology Group** |
| Debby Ngo | Novartis, Basel, Switzerland | **Clinical Groups** | **Pulmonology Group** |
| Teemu Niiranen | Finnish Institute for Health and Welfare (THL), Helsinki, Finland | **Clinical Groups** | **Cardiometabolic Diseases Group** |
| Felix Vaura | Finnish Institute for Health and Welfare (THL), Helsinki, Finland | **Clinical Groups** | **Cardiometabolic Diseases Group** |

| | | | |
|---|---|---|---|
| Veikko Salomaa | Finnish Institute for Health and Welfare (THL), Helsinki, Finland | **Clinical Groups** | **Cardiometabolic Diseases Group** |
| Kaj Metsärinne | Hospital District of Southwest Finland, Turku, Finland | **Clinical Groups** | **Cardiometabolic Diseases Group** |
| Jenni Aittokallio | Hospital District of Southwest Finland, Turku, Finland | **Clinical Groups** | **Cardiometabolic Diseases Group** |
| Mika Kähönen | Pirkanmaa Hospital District, Tampere, Finland | **Clinical Groups** | **Cardiometabolic Diseases Group** |
| Jussi Hernesniemi | Pirkanmaa Hospital District, Tampere, Finland | **Clinical Groups** | **Cardiometabolic Diseases Group** |
| Daniel Gordin | Hospital District of Helsinki and Uusimaa, Helsinki, Finland | **Clinical Groups** | **Cardiometabolic Diseases Group** |
| Juha Sinisalo | Hospital District of Helsinki and Uusimaa, Helsinki, Finland | **Clinical Groups** | **Cardiometabolic Diseases Group** |
| Marja-Riitta Taskinen | Hospital District of Helsinki and Uusimaa, Helsinki, Finland | **Clinical Groups** | **Cardiometabolic Diseases Group** |
| Tiinamaija Tuomi | Hospital District of Helsinki and Uusimaa, Helsinki, Finland | **Clinical Groups** | **Cardiometabolic Diseases Group** |
| Timo Hiltunen | Hospital District of Helsinki and Uusimaa, Helsinki, Finland | **Clinical Groups** | **Cardiometabolic Diseases Group** |
| Jari Laukkanen | Central Finland Health Care District, Jyväskylä, Finland | **Clinical Groups** | **Cardiometabolic Diseases Group** |
| Amanda Elliott | Institute for Molecular Medicine Finland (FIMM), HiLIFE, University of Helsinki, Helsinki, Finland; Broad Institute, Cambridge, MA, USA and Massachusetts General Hospital, Boston, MA, USA | **Clinical Groups** | **Cardiometabolic Diseases Group** |
| Mary Pat Reeve | Institute for Molecular Medicine Finland (FIMM), HiLIFE, University of Helsinki, Helsinki, Finland | **Clinical Groups** | **Cardiometabolic Diseases Group** |
| Sanni Ruotsalainen | Institute for Molecular Medicine Finland (FIMM), HiLIFE, University of Helsinki, Helsinki, Finland | **Clinical Groups** | **Cardiometabolic Diseases Group** |

| | | | |
|---|---|---|---|
| Benjamin Challis | Astra Zeneca, Cambridge, United Kingdom | **Clinical Groups** | **Cardiometabolic Diseases Group** |
| Dirk Paul | Astra Zeneca, Cambridge, United Kingdom | **Clinical Groups** | **Cardiometabolic Diseases Group** |
| Julie Hunkapiller | Genentech, San Francisco, CA, United States | **Clinical Groups** | **Cardiometabolic Diseases Group** |
| Natalie Bowers | Genentech, San Francisco, CA, United States | **Clinical Groups** | **Cardiometabolic Diseases Group** |
| Rion Pendergrass | Genentech, San Francisco, CA, United States | **Clinical Groups** | **Cardiometabolic Diseases Group** |
| Audrey Chu | GlaxoSmithKline, Brentford, United Kingdom | **Clinical Groups** | **Cardiometabolic Diseases Group** |
| Kirsi Auro | GlaxoSmithKline, Espoo, Finland | **Clinical Groups** | **Cardiometabolic Diseases Group** |
| Dermot Reilly | Janssen Research & Development, LLC, Boston, MA, United States | **Clinical Groups** | **Cardiometabolic Diseases Group** |
| Mike Mendelson | Novartis, Boston, MA, United States | **Clinical Groups** | **Cardiometabolic Diseases Group** |
| Jaakko Parkkinen | Pfizer, New York, NY, United States | **Clinical Groups** | **Cardiometabolic Diseases Group** |
| Melissa Miller | Pfizer, New York, NY, United States | **Clinical Groups** | **Cardiometabolic Diseases Group** |
| Tuomo Meretoja | Hospital District of Helsinki and Uusimaa, Helsinki, Finland | **Clinical Groups** | **Oncology Group** |
| Heikki Joensuu | Hospital District of Helsinki and Uusimaa, Helsinki, Finland | **Clinical Groups** | **Oncology Group** |
| Olli Carpén | Hospital District of Helsinki and Uusimaa, Helsinki, Finland | **Clinical Groups** | **Oncology Group** |
| Johanna Mattson | Hospital District of Helsinki and Uusimaa, Helsinki, Finland | **Clinical Groups** | **Oncology Group** |
| Eveliina Salminen | Hospital District of Helsinki and Uusimaa, Helsinki, Finland | **Clinical Groups** | **Oncology Group** |

| | | | |
|---|---|---|---|
| Annika Auranen | Pirkanmaa Hospital District , Tampere, Finland | **Clinical Groups** | **Oncology Group** |
| Peeter Karihtala | Northern Ostrobothnia Hospital District, Oulu, Finland | **Clinical Groups** | **Oncology Group** |
| Päivi Auvinen | Northern Savo Hospital District, Kuopio, Finland | **Clinical Groups** | **Oncology Group** |
| Klaus Elenius | Hospital District of Southwest Finland, Turku, Finland | **Clinical Groups** | **Oncology Group** |
| Johanna Schleutker | Hospital District of Southwest Finland, Turku, Finland | **Clinical Groups** | **Oncology Group** |
| Esa Pitkänen | Institute for Molecular Medicine Finland (FIMM), HiLIFE, University of Helsinki, Helsinki, Finland | **Clinical Groups** | **Oncology Group** |
| Nina Mars | Institute for Molecular Medicine Finland (FIMM), HiLIFE, University of Helsinki, Helsinki, Finland | **Clinical Groups** | **Oncology Group** |
| Mark Daly | Institute for Molecular Medicine Finland (FIMM), HiLIFE, University of Helsinki, Helsinki, Finland; Broad Institute of MIT and Harvard; Massachusetts General Hospital | **Clinical Groups** | **Oncology Group** |
| Relja Popovic | Abbvie, Chicago, IL, United States | **Clinical Groups** | **Oncology Group** |
| Jeffrey Waring | Abbvie, Chicago, IL, United States | **Clinical Groups** | **Oncology Group** |
| Bridget Riley-Gillis | Abbvie, Chicago, IL, United States | **Clinical Groups** | **Oncology Group** |
| Anne Lehtonen | Abbvie, Chicago, IL, United States | **Clinical Groups** | **Oncology Group** |
| Jennifer Schutzman | Genentech, San Francisco, CA, United States | **Clinical Groups** | **Oncology Group** |
| Julie Hunkapiller | Genentech, San Francisco, CA, United States | **Clinical Groups** | **Oncology Group** |

| | | | |
|---|---|---|---|
| Natalie Bowers | Genentech, San Francisco, CA, United States | **Clinical Groups** | **Oncology Group** |
| Rion Pendergrass | Genentech, San Francisco, CA, United States | **Clinical Groups** | **Oncology Group** |
| Diptee Kulkarni | GlaxoSmithKline, Brentford, United Kingdom | **Clinical Groups** | **Oncology Group** |
| Kirsi Auro | GlaxoSmithKline, Espoo, Finland | **Clinical Groups** | **Oncology Group** |
| Alessandro Porello | Janssen Research & Development, LLC, Spring House, PA, United States | **Clinical Groups** | **Oncology Group** |
| Andrey Loboda | Merck, Kenilworth, NJ, United States | **Clinical Groups** | **Oncology Group** |
| Heli Lehtonen | Pfizer, New York, NY, United States | **Clinical Groups** | **Oncology Group** |
| Stefan McDonough | Pfizer, New York, NY, United States | **Clinical Groups** | **Oncology Group** |
| Sauli Vuoti | Janssen-Cilag Oy, Espoo, Finland | **Clinical Groups** | **Oncology Group** |
| Kai Kaarniranta | Northern Savo Hospital District, Kuopio, Finland | **Clinical Groups** | **Opthalmology Group** |
| Joni A Turunen | Helsinki University Hospital and University of Helsinki, Helsinki, Finland; Eye Genetics Group, Folkhälsan Research Center, Helsinki, Finland | **Clinical Groups** | **Opthalmology Group** |
| Terhi Ollila | Hospital District of Helsinki and Uusimaa, Helsinki, Finland | **Clinical Groups** | **Opthalmology Group** |
| Hannu Uusitalo | Pirkanmaa Hospital District, Tampere, Finland | **Clinical Groups** | **Opthalmology Group** |
| Juha Karjalainen | Institute for Molecular Medicine Finland (FIMM), HiLIFE, University of Helsinki, Helsinki, Finland | **Clinical Groups** | **Opthalmology Group** |

| | | | |
|---|---|---|---|
| Esa Pitkänen | Institute for Molecular Medicine Finland (FIMM), HiLIFE, University of Helsinki, Helsinki, Finland | **Clinical Groups** | **Opthalmology Group** |
| Mengzhen Liu | Abbvie, Chicago, IL, United States | **Clinical Groups** | **Opthalmology Group** |
| Heiko Runz | Biogen, Cambridge, MA, United States | **Clinical Groups** | **Opthalmology Group** |
| Stephanie Loomis | Biogen, Cambridge, MA, United States | **Clinical Groups** | **Opthalmology Group** |
| Erich Strauss | Genentech, San Francisco, CA, United States | **Clinical Groups** | **Opthalmology Group** |
| Natalie Bowers | Genentech, San Francisco, CA, United States | **Clinical Groups** | **Opthalmology Group** |
| Hao Chen | Genentech, San Francisco, CA, United States | **Clinical Groups** | **Opthalmology Group** |
| Rion Pendergrass | Genentech, San Francisco, CA, United States | **Clinical Groups** | **Opthalmology Group** |
| Kaisa Tasanen | Northern Ostrobothnia Hospital District, Oulu, Finland | **Clinical Groups** | **Dermatology Group** |
| Laura Huilaja | Northern Ostrobothnia Hospital District, Oulu, Finland | **Clinical Groups** | **Dermatology Group** |
| Katariina Hannula-Jouppi | Hospital District of Helsinki and Uusimaa, Helsinki, Finland | **Clinical Groups** | **Dermatology Group** |
| Teea Salmi | Pirkanmaa Hospital District, Tampere, Finland | **Clinical Groups** | **Dermatology Group** |
| Sirkku Peltonen | Hospital District of Southwest Finland, Turku, Finland | **Clinical Groups** | **Dermatology Group** |
| Leena Koulu | Hospital District of Southwest Finland, Turku, Finland | **Clinical Groups** | **Dermatology Group** |
| Nizar Smaoui | Abbvie, Chicago, IL, United States | **Clinical Groups** | **Dermatology Group** |
| Fedik Rahimov | Abbvie, Chicago, IL, United States | **Clinical Groups** | **Dermatology Group** |

| | | | |
|---|---|---|---|
| Anne Lehtonen | Abbvie, Chicago, IL, United States | **Clinical Groups** | **Dermatology Group** |
| David Choy | Genentech, San Francisco, CA, United States | **Clinical Groups** | **Dermatology Group** |
| Rion Pendergrass | Genentech, San Francisco, CA, United States | **Clinical Groups** | **Dermatology Group** |
| Dawn Waterworth | Janssen Research & Development, LLC, Spring House, PA, United States | **Clinical Groups** | **Dermatology Group** |
| Kirsi Kalpala | Pfizer, New York, NY, United States | **Clinical Groups** | **Dermatology Group** |
| Ying Wu | Pfizer, New York, NY, United States | **Clinical Groups** | **Dermatology Group** |
| Pirkko Pussinen | Hospital District of Helsinki and Uusimaa, Helsinki, Finland | **Clinical Groups** | **Odontology Group** |
| Aino Salminen | Hospital District of Helsinki and Uusimaa, Helsinki, Finland | **Clinical Groups** | **Odontology Group** |
| Tuula Salo | Hospital District of Helsinki and Uusimaa, Helsinki, Finland | **Clinical Groups** | **Odontology Group** |
| David Rice | Hospital District of Helsinki and Uusimaa, Helsinki, Finland | **Clinical Groups** | **Odontology Group** |
| Pekka Nieminen | Hospital District of Helsinki and Uusimaa, Helsinki, Finland | **Clinical Groups** | **Odontology Group** |
| Ulla Palotie | Hospital District of Helsinki and Uusimaa, Helsinki, Finland | **Clinical Groups** | **Odontology Group** |
| Maria Siponen | Northern Savo Hospital District, Kuopio, Finland | **Clinical Groups** | **Odontology Group** |
| Liisa Suominen | Northern Savo Hospital District, Kuopio, Finland | **Clinical Groups** | **Odontology Group** |
| Päivi Mäntylä | Northern Savo Hospital District, Kuopio, Finland | **Clinical Groups** | **Odontology Group** |
| Ulvi Gursoy | Hospital District of Southwest Finland, Turku, Finland | **Clinical Groups** | **Odontology Group** |

| | | | |
|---|---|---|---|
| Vuokko Anttonen | Northern Ostrobothnia Hospital District, Oulu, Finland | **Clinical Groups** | **Odontology Group** |
| Kirsi Sipilä | Research Unit of Oral Health Sciences Faculty of Medicine, University of Oulu, Oulu, Finland; Medical Research Center, Oulu, Oulu University Hospital and University of Oulu, Oulu, Finland | **Clinical Groups** | **Odontology Group** |
| Rion Pendergrass | Genentech, San Francisco, CA, United States | **Clinical Groups** | **Odontology Group** |
| Hannele Laivuori | Institute for Molecular Medicine Finland (FIMM), HiLIFE, University of Helsinki, Helsinki, Finland | **Clinical Groups** | **Women’s Health and Reproduction Group** |
| Venla Kurra | Pirkanmaa Hospital District, Tampere, Finland | **Clinical Groups** | **Women’s Health and Reproduction Group** |
| Laura Kotaniemi-Talonen | Pirkanmaa Hospital District, Tampere, Finland | **Clinical Groups** | **Women’s Health and Reproduction Group** |
| Oskari Heikinheimo | Hospital District of Helsinki and Uusimaa, Helsinki, Finland | **Clinical Groups** | **Women’s Health and Reproduction Group** |
| Ilkka Kalliala | Hospital District of Helsinki and Uusimaa, Helsinki, Finland | **Clinical Groups** | **Women’s Health and Reproduction Group** |
| Lauri Aaltonen | Hospital District of Helsinki and Uusimaa, Helsinki, Finland | **Clinical Groups** | **Women’s Health and Reproduction Group** |
| Varpu Jokimaa | Hospital District of Southwest Finland, Turku, Finland | **Clinical Groups** | **Women’s Health and Reproduction Group** |
| Johannes Kettunen | Northern Ostrobothnia Hospital District, Oulu, Finland | **Clinical Groups** | **Women’s Health and Reproduction Group** |
| Marja Vääräsmäki | Northern Ostrobothnia Hospital District, Oulu, Finland | **Clinical Groups** | **Women’s Health and Reproduction Group** |
| Outi Uimari | Northern Ostrobothnia Hospital District, Oulu, Finland | **Clinical Groups** | **Women’s Health and Reproduction Group** |
| Laure Morin-Papunen | Northern Ostrobothnia Hospital District, Oulu, Finland | **Clinical Groups** | **Women’s Health and Reproduction Group** |

| | | | |
|---|---|---|---|
| Maarit Niinimäki | Northern Ostrobothnia Hospital District, Oulu, Finland | **Clinical Groups** | **Women's Health and Reproduction Group** |
| Terhi Piltonen | Northern Ostrobothnia Hospital District, Oulu, Finland | **Clinical Groups** | **Women's Health and Reproduction Group** |
| Katja Kivinen | Institute for Molecular Medicine Finland (FIMM), HiLIFE, University of Helsinki, Helsinki, Finland | **Clinical Groups** | **Women's Health and Reproduction Group** |
| Elisabeth Widen | Institute for Molecular Medicine Finland (FIMM), HiLIFE, University of Helsinki, Helsinki, Finland | **Clinical Groups** | **Women's Health and Reproduction Group** |
| Taru Tukiainen | Institute for Molecular Medicine Finland (FIMM), HiLIFE, University of Helsinki, Helsinki, Finland | **Clinical Groups** | **Women's Health and Reproduction Group** |
| Mary Pat Reeve | Institute for Molecular Medicine Finland (FIMM), HiLIFE, University of Helsinki, Helsinki, Finland | **Clinical Groups** | **Women's Health and Reproduction Group** |
| Mark Daly | Institute for Molecular Medicine Finland (FIMM), HiLIFE, University of Helsinki, Helsinki, Finland; Broad Institute of MIT and Harvard; Massachusetts General Hospital | **Clinical Groups** | **Women's Health and Reproduction Group** |
| Niko Välimäki | University of Helsinki, Helsinki, Finland | **Clinical Groups** | **Women's Health and Reproduction Group** |
| Eija Laakkonen | University of Jyväskylä, Jyväskylä, Finland | **Clinical Groups** | **Women's Health and Reproduction Group** |
| Jaakko Tyrmi | University of Oulu, Oulu, Finland / University of Tampere, Tampere, Finland | **Clinical Groups** | **Women's Health and Reproduction Group** |
| Heidi Silven | University of Oulu, Oulu, Finland | **Clinical Groups** | **Women's Health and Reproduction Group** |
| Eeva Sliz | University of Oulu, Oulu, Finland | **Clinical Groups** | **Women's Health and Reproduction Group** |
| Riikka Arffman | University of Oulu, Oulu, Finland | **Clinical Groups** | **Women's Health and Reproduction Group** |

| | | | |
|---|---|---|---|
| Susanna Savukoski | University of Oulu, Oulu, Finland | **Clinical Groups** | **Women's Health and Reproduction Group** |
| Triin Laisk | Estonian biobank, Tartu, Estonia | **Clinical Groups** | **Women's Health and Reproduction Group** |
| Natalia Pujol | Estonian biobank, Tartu, Estonia | **Clinical Groups** | **Women's Health and Reproduction Group** |
| Mengzhen Liu | Abbvie, Chicago, IL, United States | **Clinical Groups** | **Women's Health and Reproduction Group** |
| Bridget Riley-Gillis | Abbvie, Chicago, IL, United States | **Clinical Groups** | **Women's Health and Reproduction Group** |
| Rion Pendergrass | Genentech, San Francisco, CA, United States | **Clinical Groups** | **Women's Health and Reproduction Group** |
| Janet Kumar | GlaxoSmithKline, Collegeville, PA, United States | **Clinical Groups** | **Women's Health and Reproduction Group** |
| Kirsi Auro | GlaxoSmithKline, Espoo, Finland | **Clinical Groups** | **Women's Health and Reproduction Group** |
| Iiris Hovatta | University of Helsinki, Finland | **Clinical Groups** | **Depression group** |
| Chia-Yen Chen | Biogen, Cambridge, MA, United States | **Clinical Groups** | **Depression group** |
| Erkki Isometsä | Hospital District of Helsinki and Uusimaa, Helsinki, Finland | **Clinical Groups** | **Depression group** |
| Hanna Ollila | Institute for Molecular Medicine Finland (FIMM), HiLIFE, University of Helsinki, Helsinki, Finland | **Clinical Groups** | **Depression group** |
| Jaana Suvisaari | Finnish Institute for Health and Welfare (THL), Helsinki, Finland | **Clinical Groups** | **Depression group** |
| Thomas Damm Als | Aarhus University, Denmark | **Clinical Groups** | **Depression group** |
| Antti Mäkitie | Department of Otorhinolaryngology - Head and Neck Surgery, University of Helsinki and Helsinki University Hospital, Helsinki, Finland | **Clinical Groups** | **ENT (ear, nose and throath) Group** |

| | | | |
|---|---|---|---|
| Argyro Bizaki-Vallaskangas | Pirkanmaa Hospital District, Tampere, Finland | **Clinical Groups** | **ENT (ear, nose and throath) Group** |
| Sanna Toppila-Salmi | University of Helsinki, Finland | **Clinical Groups** | **ENT (ear, nose and throath) Group** |
| Tytti Willberg | Hospital District of Southwest Finland, Turku, Finland | **Clinical Groups** | **ENT (ear, nose and throath) Group** |
| Elmo Saarentaus | Institute for Molecular Medicine Finland (FIMM), HiLIFE, University of Helsinki, Helsinki, Finland | **Clinical Groups** | **ENT (ear, nose and throath) Group** |
| Antti Aarnisalo | Hospital District of Helsinki and Uusimaa, Helsinki, Finland | **Clinical Groups** | **ENT (ear, nose and throath) Group** |
| Eveliina Salminen | Hospital District of Helsinki and Uusimaa, Helsinki, Finland | **Clinical Groups** | **ENT (ear, nose and throath) Group** |
| Elisa Rahikkala | Northern Ostrobothnia Hospital District, Oulu, Finland | **Clinical Groups** | **ENT (ear, nose and throath) Group** |
| Johannes Kettunen | Northern Ostrobothnia Hospital District, Oulu, Finland | **Clinical Groups** | **ENT (ear, nose and throath) Group** |
| Kristiina Aittomäki | Department of Medical Genetics, Helsinki University Central Hospital, Helsinki, Finland | **Clinical Groups** | **POI (premature ovarian failure) Group** |
| Fredrik Åberg | Transplantation and Liver Surgery Clinic, Helsinki University Hospital, Helsinki University, Helsinki, Finland | **Clinical Groups** | **LiverScore Group** |
| Mitja Kurki | Institute for Molecular Medicine Finland (FIMM), HiLIFE, University of Helsinki, Helsinki, Finland; Broad Institute, Cambridge, MA, United States | **FinnGen Analysis working group** | **FinnGen Analysis working group** |
| Samuli Ripatti | Institute for Molecular Medicine Finland (FIMM), HiLIFE, University of Helsinki, Helsinki, Finland | **FinnGen Analysis working group** | **FinnGen Analysis working group** |
| Mark Daly | Institute for Molecular Medicine, Finland (FIMM), HiLIFE, University of Helsinki, Helsinki, Finland; Broad Institute of MIT and Harvard; Massachusetts General Hospital | **FinnGen Analysis working group** | **FinnGen Analysis working group** |

| | | | |
|---|---|---|---|
| Juha Karjalainen | Institute for Molecular Medicine Finland (FIMM), HiLIFE, University of Helsinki, Helsinki, Finland | **FinnGen Analysis working group** | **FinnGen Analysis working group** |
| Aki Havulinna | Institute for Molecular Medicine Finland (FIMM), HiLIFE, University of Helsinki, Helsinki, Finland; Finnish Institute for Health and Welfare (THL), Helsinki, Finland | **FinnGen Analysis working group** | **FinnGen Analysis working group** |
| Juha Mehtonen | Institute for Molecular Medicine Finland (FIMM), HiLIFE, University of Helsinki, Helsinki, Finland | **FinnGen Analysis working group** | **FinnGen Analysis working group** |
| Priit Palta | Institute for Molecular Medicine Finland (FIMM), HiLIFE, University of Helsinki, Helsinki, Finland | **FinnGen Analysis working group** | **FinnGen Analysis working group** |
| Shabbeer Hassan | Institute for Molecular Medicine Finland (FIMM), HiLIFE, University of Helsinki, Helsinki, Finland | **FinnGen Analysis working group** | **FinnGen Analysis working group** |
| Pietro Della Briotta Parolo | Institute for Molecular Medicine Finland (FIMM), HiLIFE, University of Helsinki, Helsinki, Finland | **FinnGen Analysis working group** | **FinnGen Analysis working group** |
| Wei Zhou | Broad Institute, Cambridge, MA, United States | **FinnGen Analysis working group** | **FinnGen Analysis working group** |
| Mutaamba Maasha | Broad Institute, Cambridge, MA, United States | **FinnGen Analysis working group** | **FinnGen Analysis working group** |
| Shabbeer Hassan | Institute for Molecular Medicine Finland (FIMM), HiLIFE, University of Helsinki, Helsinki, Finland | **FinnGen Analysis working group** | **FinnGen Analysis working group** |
| Susanna Lemmelä | Institute for Molecular Medicine Finland (FIMM), HiLIFE, University of Helsinki, Helsinki, Finland | **FinnGen Analysis working group** | **FinnGen Analysis working group** |

| | | | |
|---|---|---|---|
| Manuel Rivas | University of Stanford, Stanford, CA, United States | **FinnGen Analysis working group** | **FinnGen Analysis working group** |
| Mari E. Niemi | Institute for Molecular Medicine Finland (FIMM), HiLIFE, University of Helsinki, Helsinki, Finland | **FinnGen Analysis working group** | **FinnGen Analysis working group** |
| Aarno Palotie | Institute for Molecular Medicine Finland (FIMM), HiLIFE, University of Helsinki, Helsinki, Finland | **FinnGen Analysis working group** | **FinnGen Analysis working group** |
| Aoxing Liu | Institute for Molecular Medicine Finland (FIMM), HiLIFE, University of Helsinki, Helsinki, Finland | **FinnGen Analysis working group** | **FinnGen Analysis working group** |
| Arto Lehisto | Institute for Molecular Medicine Finland (FIMM), HiLIFE, University of Helsinki, Helsinki, Finland | **FinnGen Analysis working group** | **FinnGen Analysis working group** |
| Andrea Ganna | Institute for Molecular Medicine Finland (FIMM), HiLIFE, University of Helsinki, Helsinki, Finland | **FinnGen Analysis working group** | **FinnGen Analysis working group** |
| Vincent Llorens | Institute for Molecular Medicine Finland (FIMM), HiLIFE, University of Helsinki, Helsinki, Finland | **FinnGen Analysis working group** | **FinnGen Analysis working group** |
| Hannele Laivuori | Institute for Molecular Medicine Finland (FIMM), HiLIFE, University of Helsinki, Helsinki, Finland | **FinnGen Analysis working group** | **FinnGen Analysis working group** |
| Taru Tukiainen | Institute for Molecular Medicine Finland (FIMM), HiLIFE, University of Helsinki, Helsinki, Finland | **FinnGen Analysis working group** | **FinnGen Analysis working group** |
| Mary Pat Reeve | Institute for Molecular Medicine Finland (FIMM), HiLIFE, University of Helsinki, Helsinki, Finland | **FinnGen Analysis working group** | **FinnGen Analysis working group** |
| Henrike Heyne | Institute for Molecular Medicine Finland (FIMM), HiLIFE, University of Helsinki, Helsinki, Finland | **FinnGen Analysis working group** | **FinnGen Analysis working group** |

| | | | |
|---|---|---|---|
| Nina Mars | Institute for Molecular Medicine Finland (FIMM), HiLIFE, University of Helsinki, Helsinki, Finland | **FinnGen Analysis working group** | **FinnGen Analysis working group** |
| Joel Rämö | Institute for Molecular Medicine Finland (FIMM), HiLIFE, University of Helsinki, Helsinki, Finland | **FinnGen Analysis working group** | **FinnGen Analysis working group** |
| Elmo Saarentaus | Institute for Molecular Medicine Finland (FIMM), HiLIFE, University of Helsinki, Helsinki, Finland | **FinnGen Analysis working group** | **FinnGen Analysis working group** |
| Hanna Ollila | Institute for Molecular Medicine Finland (FIMM), HiLIFE, University of Helsinki, Helsinki, Finland | **FinnGen Analysis working group** | **FinnGen Analysis working group** |
| Rodos Rodosthenous | Institute for Molecular Medicine Finland (FIMM), HiLIFE, University of Helsinki, Helsinki, Finland | **FinnGen Analysis working group** | **FinnGen Analysis working group** |
| Satu Strausz | Institute for Molecular Medicine Finland (FIMM), HiLIFE, University of Helsinki, Helsinki, Finland | **FinnGen Analysis working group** | **FinnGen Analysis working group** |
| Tuula Palotie | University of Helsinki and Hospital District of Helsinki and Uusimaa, Helsinki, Finland | **FinnGen Analysis working group** | **FinnGen Analysis working group** |
| Kimmo Palin | University of Helsinki, Helsinki, Finland | **FinnGen Analysis working group** | **FinnGen Analysis working group** |
| Javier Garcia-Tabuenca | University of Tampere, Tampere, Finland | **FinnGen Analysis working group** | **FinnGen Analysis working group** |
| Harri Siirtola | University of Tampere, Tampere, Finland | **FinnGen Analysis working group** | **FinnGen Analysis working group** |
| Tuomo Kiiskinen | Institute for Molecular Medicine Finland (FIMM), HiLIFE, University of Helsinki, Helsinki, Finland | **FinnGen Analysis working group** | **FinnGen Analysis working group** |

| | | | |
|---|---|---|---|
| Jiwoo Lee | Institute for Molecular Medicine Finland (FIMM), HiLIFE, University of Helsinki, Helsinki, Finland; Broad Institute, Cambridge, MA, United States | **FinnGen Analysis working group** | **FinnGen Analysis working group** |
| Kristin Tsuo | Institute for Molecular Medicine Finland (FIMM), HiLIFE, University of Helsinki, Helsinki, Finland; Broad Institute, Cambridge, MA, United States | **FinnGen Analysis working group** | **FinnGen Analysis working group** |
| Amanda Elliott | Institute for Molecular Medicine Finland (FIMM), HiLIFE, University of Helsinki, Helsinki, Finland; Broad Institute, Cambridge, MA, USA and Massachusetts General Hospital, Boston, MA, USA | **FinnGen Analysis working group** | **FinnGen Analysis working group** |
| Kati Kristiansson | THL Biobank / Finnish Institute for Health and Welfare (THL), Helsinki, Finland | **FinnGen Analysis working group** | **FinnGen Analysis working group** |
| Mikko Arvas | Finnish Red Cross Blood Service / Finnish Hematology Registry and Clinical Biobank, Helsinki, Finland | **FinnGen Analysis working group** | **FinnGen Analysis working group** |
| Kati Hyvärinen | Finnish Red Cross Blood Service, Helsinki, Finland | **FinnGen Analysis working group** | **FinnGen Analysis working group** |
| Jarmo Ritari | Finnish Red Cross Blood Service, Helsinki, Finland | **FinnGen Analysis working group** | **FinnGen Analysis working group** |
| Olli Carpén | Helsinki Biobank / Helsinki University and Hospital District of Helsinki and Uusimaa, Helsinki | **FinnGen Analysis working group** | **FinnGen Analysis working group** |
| Johannes Kettunen | Northern Finland Biobank Borealis / University of Oulu / Northern Ostrobothnia Hospital District, Oulu, Finland | **FinnGen Analysis working group** | **FinnGen Analysis working group** |

| | | | |
|---|---|---|---|
| Katri Pylkäs | University of Oulu, Oulu, Finland | **FinnGen Analysis working group** | **FinnGen Analysis working group** |
| Eeva Sliz | University of Oulu, Oulu, Finland | **FinnGen Analysis working group** | **FinnGen Analysis working group** |
| Minna Karjalainen | University of Oulu, Oulu, Finland | **FinnGen Analysis working group** | **FinnGen Analysis working group** |
| Tuomo Mantere | Northern Finland Biobank Borealis / University of Oulu / Northern Ostrobothnia Hospital District, Oulu, Finland | **FinnGen Analysis working group** | **FinnGen Analysis working group** |
| Eeva Kangasniemi | Finnish Clinical Biobank Tampere / University of Tampere / Pirkanmaa Hospital District, Tampere, Finland | **FinnGen Analysis working group** | **FinnGen Analysis working group** |
| Sami Heikkinen | University of Eastern Finland, Kuopio, Finland | **FinnGen Analysis working group** | **FinnGen Analysis working group** |
| Arto Mannermaa | Biobank of Eastern Finland / University of Eastern Finland / Northern Savo Hospital District, Kuopio, Finland | **FinnGen Analysis working group** | **FinnGen Analysis working group** |
| Eija Laakkonen | University of Jyväskylä, Jyväskylä, Finland | **FinnGen Analysis working group** | **FinnGen Analysis working group** |
| Nina Pitkänen | Auria Biobank / University of Turku / Hospital District of Southwest Finland, Turku, Finland | **FinnGen Analysis working group** | **FinnGen Analysis working group** |
| Samuel Lessard | Translational Sciences, Sanofi R&D, Framingham, MA, USA | **FinnGen Analysis working group** | **FinnGen Analysis working group** |
| Clément Chatelain | Translational Sciences, Sanofi R&D, Framingham, MA, USA | **FinnGen Analysis working group** | **FinnGen Analysis working group** |

| | | | |
|---|---|---|---|
| Perttu Terho | Auria Biobank / University of Turku / Hospital District of Southwest Finland, Turku, Finland | **Biobank directors** | **Biobank directors** |
| Sirpa Soini | THL Biobank / Finnish Institute for Health and Welfare (THL), Helsinki, Finland | **Biobank directors** | **Biobank directors** |
| Jukka Partanen | Finnish Red Cross Blood Service / Finnish Hematology Registry and Clinical Biobank, Helsinki, Finland | **Biobank directors** | **Biobank directors** |
| Eero Punkka | Helsinki Biobank / Helsinki University and Hospital District of Helsinki and Uusimaa, Helsinki | **Biobank directors** | **Biobank directors** |
| Raisa Serpi | Northern Finland Biobank Borealis / University of Oulu / Northern Ostrobothnia Hospital District, Oulu, Finland | **Biobank directors** | **Biobank directors** |
| Sanna Siltanen | Finnish Clinical Biobank Tampere / University of Tampere / Pirkanmaa Hospital District, Tampere, Finland | **Biobank directors** | **Biobank directors** |
| Veli-Matti Kosma | Biobank of Eastern Finland / University of Eastern Finland / Northern Savo Hospital District, Kuopio, Finland | **Biobank directors** | **Biobank directors** |
| Teijo Kuopio | Central Finland Biobank / University of Jyväskylä / Central Finland Health Care District, Jyväskylä, Finland | **Biobank directors** | **Biobank directors** |
| Anu Jalanko | Institute for Molecular Medicine Finland (FIMM), HiLIFE, University of Helsinki, Helsinki, Finland | **FinnGen Teams** | **Administration** |
| Huei-Yi Shen | Institute for Molecular Medicine Finland (FIMM), HiLIFE, University of Helsinki, Helsinki, Finland | **FinnGen Teams** | **Administration** |
| Risto Kajanne | Institute for Molecular Medicine Finland (FIMM), HiLIFE, University of Helsinki, Helsinki, Finland | **FinnGen Teams** | **Administration** |

| | | | |
|---|---|---|---|
| Mervi Aavikko | Institute for Molecular Medicine Finland (FIMM), HiLIFE, University of Helsinki, Helsinki, Finland | **FinnGen Teams** | **Administration** |
| Henna Palin | Finnish Clinical Biobank Tampere / University of Tampere / Pirkanmaa Hospital District, Tampere, Finland | **FinnGen Teams** | **Administration** |
| Malla-Maria Linna | Helsinki Biobank / Helsinki University and Hospital District of Helsinki and Uusimaa, Helsinki | **FinnGen Teams** | **Administration** |
| Mitja Kurki | Institute for Molecular Medicine Finland (FIMM), HiLIFE, University of Helsinki, Helsinki, Finland; Broad Institute, Cambridge, MA, United States | **FinnGen Teams** | **Analysis** |
| Juha Karjalainen | Institute for Molecular Medicine Finland (FIMM), HiLIFE, University of Helsinki, Helsinki, Finland | **FinnGen Teams** | **Analysis** |
| Pietro Della Briotta Parolo | Institute for Molecular Medicine Finland (FIMM), HiLIFE, University of Helsinki, Helsinki, Finland | **FinnGen Teams** | **Analysis** |
| Arto Lehisto | Institute for Molecular Medicine Finland (FIMM), HiLIFE, University of Helsinki, Helsinki, Finland | **FinnGen Teams** | **Analysis** |
| Juha Mehtonen | Institute for Molecular Medicine Finland (FIMM), HiLIFE, University of Helsinki, Helsinki, Finland | **FinnGen Teams** | **Analysis** |
| Wei Zhou | Broad Institute, Cambridge, MA, United States | **FinnGen Teams** | **Analysis** |
| Masahiro Kanai | Broad Institute, Cambridge, MA, United States | **FinnGen Teams** | **Analysis** |
| Mutaamba Maasha | Broad Institute, Cambridge, MA, United States | **FinnGen Teams** | **Analysis** |

| | | | |
|---|---|---|---|
| Hannele Laivuori | Institute for Molecular Medicine Finland (FIMM), HiLIFE, University of Helsinki, Helsinki, Finland | **FinnGen Teams** | **Clinical Endpoint Development** |
| Aki Havulinna | Institute for Molecular Medicine Finland (FIMM), HiLIFE, University of Helsinki, Helsinki, Finland; Finnish Institute for Health and Welfare (THL), Helsinki, Finland | **FinnGen Teams** | **Clinical Endpoint Development** |
| Susanna Lemmelä | Institute for Molecular Medicine Finland (FIMM), HiLIFE, University of Helsinki, Helsinki, Finland | **FinnGen Teams** | **Clinical Endpoint Development** |
| Tuomo Kiiskinen | Institute for Molecular Medicine Finland (FIMM), HiLIFE, University of Helsinki, Helsinki, Finland | **FinnGen Teams** | **Clinical Endpoint Development** |
| L. Elisa Lahtela | Institute for Molecular Medicine Finland (FIMM), HiLIFE, University of Helsinki, Helsinki, Finland | **FinnGen Teams** | **Clinical Endpoint Development** |
| Mari Kaunisto | Institute for Molecular Medicine Finland (FIMM), HiLIFE, University of Helsinki, Helsinki, Finland | **FinnGen Teams** | **Communication** |
| Elina Kilpeläinen | Institute for Molecular Medicine Finland (FIMM), HiLIFE, University of Helsinki, Helsinki, Finland | **FinnGen Teams** | **E-Science** |
| Timo P. Sipilä | Institute for Molecular Medicine Finland (FIMM), HiLIFE, University of Helsinki, Helsinki, Finland | **FinnGen Teams** | **E-Science** |
| Oluwaseun Alexander Dada | Institute for Molecular Medicine Finland (FIMM), HiLIFE, University of Helsinki, Helsinki, Finland | **FinnGen Teams** | **E-Science** |
| Awaisa Ghazal | Institute for Molecular Medicine Finland (FIMM), HiLIFE, University of Helsinki, Helsinki, Finland | **FinnGen Teams** | **E-Science** |

| | | | |
|---|---|---|---|
| Anastasia Kytölä | Institute for Molecular Medicine Finland (FIMM), HiLIFE, University of Helsinki, Helsinki, Finland | **FinnGen Teams** | **E-Science** |
| Rigbe Weldatsadik | Institute for Molecular Medicine Finland (FIMM), HiLIFE, University of Helsinki, Helsinki, Finland | **FinnGen Teams** | **E-Science** |
| Sanni Ruotsalainen | Institute for Molecular Medicine Finland (FIMM), HiLIFE, University of Helsinki, Helsinki, Finland | **FinnGen Teams** | **E-Science** |
| Kati Donner | Institute for Molecular Medicine Finland (FIMM), HiLIFE, University of Helsinki, Helsinki, Finland | **FinnGen Teams** | **Genotyping** |
| Timo P. Sipilä | Institute for Molecular Medicine Finland (FIMM), HiLIFE, University of Helsinki, Helsinki, Finland | **FinnGen Teams** | **Genotyping** |
| Anu Loukola | Helsinki Biobank / Helsinki University and Hospital District of Helsinki and Uusimaa, Helsinki | **FinnGen Teams** | **Sample Collection Coordination** |
| Päivi Laiho | THL Biobank / Finnish Institute for Health and Welfare (THL), Helsinki, Finland | **FinnGen Teams** | **Sample Logistics** |
| Tuuli Sistonen | THL Biobank / Finnish Institute for Health and Welfare (THL), Helsinki, Finland | **FinnGen Teams** | **Sample Logistics** |
| Essi Kaiharju | THL Biobank / Finnish Institute for Health and Welfare (THL), Helsinki, Finland | **FinnGen Teams** | **Sample Logistics** |
| Markku Laukkanen | THL Biobank / Finnish Institute for Health and Welfare (THL), Helsinki, Finland | **FinnGen Teams** | **Sample Logistics** |
| Elina Järvensivu | THL Biobank / Finnish Institute for Health and Welfare (THL), Helsinki, Finland | **FinnGen Teams** | **Sample Logistics** |
| Sini Lähteenmäki | THL Biobank / Finnish Institute for Health and Welfare (THL), Helsinki, Finland | **FinnGen Teams** | **Sample Logistics** |
| Lotta Männikkö | THL Biobank / Finnish Institute for Health and Welfare (THL), Helsinki, Finland | **FinnGen Teams** | **Sample Logistics** |

| | | | |
|---|---|---|---|
| Regis Wong | THL Biobank / Finnish Institute for Health and Welfare (THL), Helsinki, Finland | **FinnGen Teams** | **Sample Logistics** |
| Auli Toivola | THL Biobank / Finnish Institute for Health and Welfare (THL), Helsinki, Finland | **FinnGen Teams** | **Sample Logistics** |
| Minna Brunfeldt | THL Biobank / Finnish Institute for Health and Welfare (THL), Helsinki, Finland | **FinnGen Teams** | **Registry Data Operations** |
| Hannele Mattsson | THL Biobank / Finnish Institute for Health and Welfare (THL), Helsinki, Finland | **FinnGen Teams** | **Registry Data Operations** |
| Kati Kristiansson | THL Biobank / Finnish Institute for Health and Welfare (THL), Helsinki, Finland | **FinnGen Teams** | **Registry Data Operations** |
| Susanna Lemmelä | Institute for Molecular Medicine Finland (FIMM), HiLIFE, University of Helsinki, Helsinki, Finland | **FinnGen Teams** | **Registry Data Operations** |
| Sami Koskelainen | THL Biobank / Finnish Institute for Health and Welfare (THL), Helsinki, Finland | **FinnGen Teams** | **Registry Data Operations** |
| Tero Hiekkalinna | THL Biobank / Finnish Institute for Health and Welfare (THL), Helsinki, Finland | **FinnGen Teams** | **Registry Data Operations** |
| Teemu Paajanen | THL Biobank / Finnish Institute for Health and Welfare (THL), Helsinki, Finland | **FinnGen Teams** | **Registry Data Operations** |
| Priit Palta | Institute for Molecular Medicine Finland (FIMM), HiLIFE, University of Helsinki, Helsinki, Finland | **FinnGen Teams** | **Sequencing Informatics** |
| Kalle Pärn | Institute for Molecular Medicine Finland (FIMM), HiLIFE, University of Helsinki, Helsinki, Finland | **FinnGen Teams** | **Sequencing Informatics** |
| Mart Kals | Institute for Molecular Medicine Finland (FIMM), HiLIFE, University of Helsinki, Helsinki, Finland | **FinnGen Teams** | **Sequencing Informatics** |
| Shuang Luo | Institute for Molecular Medicine Finland (FIMM), HiLIFE, University of Helsinki, Helsinki, Finland | **FinnGen Teams** | **Sequencing Informatics** |
| Tarja Laitinen | Pirkanmaa Hospital District, Tampere, Finland | **FinnGen Teams** | **Trajectory** |

| | | | |
|---|---|---|---|
| Mary Pat Reeve | Institute for Molecular Medicine Finland (FIMM), HiLIFE, University of Helsinki, Helsinki, Finland | **FinnGen Teams** | **Trajectory** |
| Shanmukha Sampath Padmanabhuni | Institute for Molecular Medicine Finland (FIMM), HiLIFE, University of Helsinki, Helsinki, Finland | **FinnGen Teams** | **Trajectory** |
| Marianna Niemi | University of Tampere, Tampere, Finland | **FinnGen Teams** | **Trajectory** |
| Harri Siirtola | University of Tampere, Tampere, Finland | **FinnGen Teams** | **Trajectory** |
| Javier Gracia-Tabuenca | University of Tampere, Tampere, Finland | **FinnGen Teams** | **Trajectory** |
| Mika Helminen | University of Tampere, Tampere, Finland | **FinnGen Teams** | **Trajectory** |
| Tiina Luukkaala | University of Tampere, Tampere, Finland | **FinnGen Teams** | **Trajectory** |
| Iida Vähätalo | University of Tampere, Tampere, Finland | **FinnGen Teams** | **Trajectory** |
| Jyrki Pitkänen | Institute for Molecular Medicine Finland (FIMM), HiLIFE, University of Helsinki, Helsinki, Finland | **FinnGen Teams** | **Data protection officer** |
| Marco Hautalahti | Finnish Biobank Cooperative - FINBB | **FinnGen Teams** | **FINBB - Finnish biobank cooperative** |
| Johanna Mäkelä | Finnish Biobank Cooperative - FINBB | **FinnGen Teams** | **FINBB - Finnish biobank cooperative** |
| Sarah Smith | Finnish Biobank Cooperative - FINBB | **FinnGen Teams** | **FINBB - Finnish biobank cooperative** |
| Tom Southerington | Finnish Biobank Cooperative - FINBB | **FinnGen Teams** | **FINBB - Finnish biobank cooperative** |

**Million Veteran Program**

MVP Program Office

- Sumitra Muralidhar, Ph.D., Program Director

US Department of Veterans Affairs, 810 Vermont Avenue NW, Washington, DC 20420

- Jennifer Moser, Ph.D., Associate Director, Scientific Programs

US Department of Veterans Affairs, 810 Vermont Avenue NW, Washington, DC 20420

- Jennifer E. Deen, B.S., Associate Director, Cohort & Public Relations

US Department of Veterans Affairs, 810 Vermont Avenue NW, Washington, DC 20420

MVP Executive Committee

- Co-Chair: Philip S. Tsao, Ph.D.

VA Palo Alto Health Care System, 3801 Miranda Avenue, Palo Alto, CA 94304

- Co-Chair: Sumitra Muralidhar, Ph.D.

US Department of Veterans Affairs, 810 Vermont Avenue NW, Washington, DC 20420

- J. Michael Gaziano, M.D., M.P.H.

VA Boston Healthcare System, 150 S. Huntington Avenue, Boston, MA 02130

- Elizabeth Hauser, Ph.D.

Durham VA Medical Center, 508 Fulton Street, Durham, NC 27705

- Amy Kilbourne, Ph.D., M.P.H.

VA HSR&D, 2215 Fuller Road, Ann Arbor, MI 48105

- Shiuh-Wen Luoh, M.D., Ph.D.

VA Portland Health Care System, 3710 SW US Veterans Hospital Rd, Portland, OR 97239

- Michael Matheny, M.D., M.S., M.P.H.

VA Tennessee Valley Healthcare System, 1310 24$^{th}$ Ave. South, Nashville, TN 37212

- Dave Oslin, M.D.

Philadelphia VA Medical Center, 3900 Woodland Avenue, Philadelphia, PA 19104

MVP Co-Principal Investigators

- J. Michael Gaziano, M.D., M.P.H.

VA Boston Healthcare System, 150 S. Huntington Avenue, Boston, MA 02130

- Philip S. Tsao, Ph.D.

VA Palo Alto Health Care System, 3801 Miranda Avenue, Palo Alto, CA 94304

MVP Core Operations

- Lori Churby, B.S., Director, MVP Regulatory Affairs

VA Palo Alto Health Care System, 3801 Miranda Avenue, Palo Alto, CA 94304

- Stacey B. Whitbourne, Ph.D., Director, MVP Cohort Management

VA Boston Healthcare System, 150 S. Huntington Avenue, Boston, MA 02130

- Jessica V. Brewer, M.P.H., Director, MVP Recruitment & Enrollment

VA Boston Healthcare System, 150 S. Huntington Avenue, Boston, MA 02130

- Shahpoor (Alex) Shayan, M.S., Director, MVP Recruitment and Enrollment Informatics

VA Boston Healthcare System, 150 S. Huntington Avenue, Boston, MA 02130

- Luis E. Selva, Ph.D., Executive Director, MVP Biorepositories

VA Boston Healthcare System, 150 S. Huntington Avenue, Boston, MA 02130

- Saiju Pyarajan Ph.D., Director, Data and Computational Sciences

VA Boston Healthcare System, 150 S. Huntington Avenue, Boston, MA 02130

- Kelly Cho, M.P.H, Ph.D., Director, MVP Phenomics Data Core

VA Boston Healthcare System, 150 S. Huntington Avenue, Boston, MA 02130

- Scott L. DuVall, Ph.D., Director, VA Informatics and Computing Infrastructure (VINCI)

VA Salt Lake City Health Care System, 500 Foothill Drive, Salt Lake City, UT 84148

- Mary T. Brophy M.D., M.P.H., Director, VA Central Biorepository

VA Boston Healthcare System, 150 S. Huntington Avenue, Boston, MA 02130

- MVP Coordinating Centers
    - MVP Coordinating Center, Boston - J. Michael Gaziano, M.D., M.P.H.

VA Boston Healthcare System, 150 S. Huntington Avenue, Boston, MA 02130

- MVP Coordinating Center, Palo Alto – Philip S. Tsao, Ph.D.

VA Palo Alto Health Care System, 3801 Miranda Avenue, Palo Alto, CA 94304

- MVP Information Center, Canandaigua – Brady Stephens, M.S.

Canandaigua VA Medical Center, 400 Fort Hill Avenue, Canandaigua, NY 14424

- Cooperative Studies Program Clinical Research Pharmacy Coordinating Center, Albuquerque – Todd Connor, Pharm.D.; Dean P. Argyres, B.S., M.S.

New Mexico VA Health Care System, 1501 San Pedro Drive SE, Albuquerque, NM 87108

MVP Publications and Presentations Committee

- Co-Chair: Tim Assimes, M.D.
VA Palo Alto Health Care System, 3801 Miranda Avenue, Palo Alto, CA 94304
- Co-Chair: Adriana Hung, M.D.
VA Tennessee Valley Healthcare System, 1310 24th Ave. South, Nashville, TN 37212
- Co-Chair: Henry Kranzler, M.D.
Philadelphia VA Medical Center, 3900 Woodland Avenue, Philadelphia, PA 19104

MVP Local Site Investigators

- Samuel Aguayo, M.D., Phoenix VA Health Care System
650 E. Indian School Road, Phoenix, AZ 85012
- Sunil Ahuja, M.D., South Texas Veterans Health Care System
7400 Merton Minter Boulevard, San Antonio, TX 78229
- Kathrina Alexander, M.D., Veterans Health Care System of the Ozarks
1100 North College Avenue, Fayetteville, AR 72703
- Xiao M. Androulakis, M.D., Columbia VA Health Care System
6439 Garners Ferry Road, Columbia, SC 29209
- Prakash Balasubramanian, M.D., William S. Middleton Memorial Veterans Hospital
2500 Overlook Terrace, Madison, WI 53705
- Zuhair Ballas, M.D., Iowa City VA Health Care System
601 Highway 6 West, Iowa City, IA 52246-2208
- Jean Beckham, Ph.D., Durham VA Medical Center
508 Fulton Street, Durham, NC 27705
- Sujata Bhushan, M.D., VA North Texas Health Care System
4500 S. Lancaster Road, Dallas, TX 75216
- Edward Boyko, M.D., VA Puget Sound Health Care System
1660 S. Columbian Way, Seattle, WA 98108-1597
- David Cohen, M.D., Portland VA Medical Center
3710 SW U.S. Veterans Hospital Road, Portland, OR 97239
- Louis Dellitalia, M.D., Birmingham VA Medical Center
700 S. 19th Street, Birmingham AL 35233

- L. Christine Faulk, M.D., Robert J. Dole VA Medical Center

5500 East Kellogg Drive, Wichita, KS 67218-1607

- Joseph Fayad, M.D., VA Southern Nevada Healthcare System

6900 North Pecos Road, North Las Vegas, NV 89086

- Daryl Fujii, Ph.D., VA Pacific Islands Health Care System

459 Patterson Rd, Honolulu, HI 96819

- Saib Gappy, M.D., John D. Dingell VA Medical Center

4646 John R Street, Detroit, MI 48201

- Frank Gesek, Ph.D., White River Junction VA Medical Center

163 Veterans Drive, White River Junction, VT 05009

- Jennifer Greco, M.D., Sioux Falls VA Health Care System

2501 W 22nd Street, Sioux Falls, SD 57105

- Michael Godschalk, M.D., Richmond VA Medical Center

1201 Broad Rock Blvd., Richmond, VA 23249

- Todd W. Gress, M.D., Ph.D., Hershel “Woody” Williams VA Medical Center

1540 Spring Valley Drive, Huntington, WV 25704

- Samir Gupta, M.D., M.S.C.S., VA San Diego Healthcare System

3350 La Jolla Village Drive, San Diego, CA 92161

- Salvador Gutierrez, M.D., Edward Hines, Jr. VA Medical Center

5000 South 5th Avenue, Hines, IL 60141

- John Harley, M.D., Ph.D., Cincinnati VA Medical Center

3200 Vine Street, Cincinnati, OH 45220

- Kimberly Hammer, Ph.D., Fargo VA Health Care System

2101 N. Elm, Fargo, ND 58102

- Mark Hamner, M.D., Ralph H. Johnson VA Medical Center

109 Bee Street, Mental Health Research, Charleston, SC 29401

- Adriana Hung, M.D., M.P.H., VA Tennessee Valley Healthcare System

1310 24th Avenue, South Nashville, TN 37212

- Robin Hurley, M.D., W.G. (Bill) Hefner VA Medical Center

1601 Brenner Ave, Salisbury, NC 28144

- Pran Iruvanti, D.O., Ph.D., Hampton VA Medical Center

100 Emancipation Drive, Hampton, VA 23667

- Frank Jacono, M.D., VA Northeast Ohio Healthcare System

10701 East Boulevard, Cleveland, OH 44106
- Darshana Jhala, M.D., Philadelphia VA Medical Center

3900 Woodland Avenue, Philadelphia, PA 19104
- Scott Kinlay, M.B.B.S., Ph.D., VA Boston Healthcare System

150 S. Huntington Avenue, Boston, MA 02130
- Jon Klein, M.D., Ph.D., Louisville VA Medical Center

800 Zorn Avenue, Louisville, KY 40206
- Michael Landry, Ph.D., Southeast Louisiana Veterans Health Care System

2400 Canal Street, New Orleans, LA 70119
- Peter Liang, M.D., M.P.H., VA New York Harbor Healthcare System

423 East 23rd Street, New York, NY 10010
- Suthat Liangpunsakul, M.D., M.P.H., Richard Roudebush VA Medical Center

1481 West 10th Street, Indianapolis, IN 46202
- Jack Lichy, M.D., Ph.D., Washington DC VA Medical Center

50 Irving St, Washington, D. C. 20422
- C. Scott Mahan, M.D., Charles George VA Medical Center

1100 Tunnel Road, Asheville, NC 28805
- Ronnie Marrache, M.D., VA Maine Healthcare System

1 VA Center, Augusta, ME 04330
- Stephen Mastorides, M.D., James A. Haley Veterans' Hospital

13000 Bruce B. Downs Blvd, Tampa, FL 33612
- Elisabeth Mates M.D., Ph.D., VA Sierra Nevada Health Care System

975 Kirman Avenue, Reno, NV 89502
- Kristin Mattocks, Ph.D., M.P.H., Central Western Massachusetts Healthcare System

421 North Main Street, Leeds, MA 01053
- Paul Meyer, M.D., Ph.D., Southern Arizona VA Health Care System

3601 S 6th Avenue, Tucson, AZ 85723
- Jonathan Moorman, M.D., Ph.D., James H. Quillen VA Medical Center

Corner of Lamont & Veterans Way, Mountain Home, TN 37684
- Timothy Morgan, M.D., VA Long Beach Healthcare System

5901 East 7th Street Long Beach, CA 90822
- Maureen Murdoch, M.D., M.P.H., Minneapolis VA Health Care System

One Veterans Drive, Minneapolis, MN 55417

- James Norton, Ph.D., VA Health Care Upstate New York

113 Holland Avenue, Albany, NY 12208

- Olaoluwa Okusaga, M.D., Michael E. DeBakey VA Medical Center

2002 Holcombe Blvd, Houston, TX 77030

- Kris Ann Oursler, M.D., Salem VA Medical Center

1970 Roanoke Blvd, Salem, VA 24153

- Ana Palacio, M.D., M.P.H., Miami VA Health Care System

1201 NW 16th Street, 11 GRC, Miami FL 33125

- Samuel Poon, M.D., Manchester VA Medical Center

718 Smyth Road, Manchester, NH 03104

- Emily Potter, Pharm.D., VA Eastern Kansas Health Care System

4101 S 4th Street Trafficway, Leavenworth, KS 66048

- Michael Rauchman, M.D., St. Louis VA Health Care System

915 North Grand Blvd, St. Louis, MO 63106

- Richard Servatius, Ph.D., Syracuse VA Medical Center

800 Irving Avenue, Syracuse, NY 13210

- Satish Sharma, M.D., Providence VA Medical Center

830 Chalkstone Avenue, Providence, RI 02908

- River Smith, Ph.D., Eastern Oklahoma VA Health Care System

1011 Honor Heights Drive, Muskogee, OK 74401

- Peruvemba Sriram, M.D., N. FL/S. GA Veterans Health System

1601 SW Archer Road, Gainesville, FL 32608

- Patrick Strollo, Jr., M.D., VA Pittsburgh Health Care System

University Drive, Pittsburgh, PA 15240

- Neeraj Tandon, M.D., Overton Brooks VA Medical Center

510 East Stoner Ave, Shreveport, LA 71101

- Philip Tsao, Ph.D., VA Palo Alto Health Care System

3801 Miranda Avenue, Palo Alto, CA 94304-1290

- Gerardo Villareal, M.D., New Mexico VA Health Care System

1501 San Pedro Drive, S.E. Albuquerque, NM 87108

- Agnes Wallbom, M.D., M.S., VA Greater Los Angeles Health Care System

11301 Wilshire Blvd, Los Angeles, CA 90073

- Jessica Walsh, M.D., VA Salt Lake City Health Care System

500 Foothill Drive, Salt Lake City, UT 84148
- John Wells, Ph.D., Edith Nourse Rogers Memorial Veterans Hospital
200 Springs Road, Bedford, MA 01730
- Jeffrey Whittle, M.D., M.P.H., Clement J. Zablocki VA Medical Center
5000 West National Avenue, Milwaukee, WI 53295
- Mary Whooley, M.D., San Francisco VA Health Care System
4150 Clement Street, San Francisco, CA 94121
- Allison E. Williams, N.D., Ph.D., R.N, Bay Pines VA Healthcare System
10,000 Bay Pines Blvd Bay Pines, FL 33744
- Peter Wilson, M.D., Atlanta VA Medical Center
1670 Clairmont Road, Decatur, GA 30033
- Junzhe Xu, M.D., VA Western New York Healthcare System
3495 Bailey Avenue, Buffalo, NY 14215-1199
- Shing Shing Yeh, Ph.D., M.D., Northport VA Medical Center
79 Middleville Road, Northport, NY 11768

**Geisinger MyCode**

Regeneron Genetics Center Banner Author List and Contribution Statements

All authors/contributors are listed in alphabetical order.

<u>RGC Management and Leadership Team</u>
Goncalo Abecasis, Ph.D., Aris Baras, M.D., Michael Cantor, M.D., Giovanni Coppola, M.D., Aris Economides, Ph.D., Luca A. Lotta, M.D., Ph.D., John D. Overton, Ph.D., Jeffrey G. Reid, Ph.D., Alan Shuldiner, M.D.

Contribution: All authors contributed to securing funding, study design and oversight. All authors reviewed the final version of the manuscript.

<u>Sequencing and Lab Operations</u>

Christina Beechert, Caitlin Forsythe, M.S., Erin D. Fuller, Zhenhua Gu, M.S., Michael Lattari, Alexander Lopez, M.S., John D. Overton, Ph.D., Thomas D. Schleicher, M.S., Maria Sotiropoulos Padilla, M.S., Louis Widom, Sarah E. Wolf, M.S., Manasi Pradhan, M.S., Kia Manoochehri, Ricardo H. Ulloa.

Contribution: C.B., C.F., A.L., and J.D.O. performed and are responsible for sample genotyping. C.B, C.F., E.D.F., M.L., M.S.P., L.W., S.E.W., A.L., and J.D.O. performed and are responsible for exome sequencing. T.D.S., Z.G., A.L., and J.D.O. conceived and are responsible for laboratory automation. M.P., K.M., R.U., and J.D.O are responsible for sample tracking and the library information management system.

Genome Informatics

Xiaodong Bai, Ph.D., Suganthi Balasubramanian, Ph.D., Andrew Blumenfeld, Boris Boutkov, Ph.D., Gisu Eom, Lukas Habegger, Ph.D., Alicia Hawes, B.S., Shareef Khalid, Olga Krasheninina, M.S., Rouel Lanche, Adam J. Mansfield, B.A., Evan K. Maxwell, Ph.D., Mrunali Nafde, Sean O'Keeffe, M.S., Max Orelus, Razvan Panea, Ph.D., Tommy Polanco, B.A., Ayesha Rasool, M.S., Jeffrey G. Reid, Ph.D., William Salerno, Ph.D., Jeffrey C. Staples, Ph.D.

Contribution: X.B., A.H., O.K., A.M., S.O., R.P., T.P., A.R., W.S. and J.G.R. performed and are responsible for the compute logistics, analysis and infrastructure needed to produce exome and genotype data. G.E., M.O., M.N. and J.G.R. provided compute infrastructure development and operational support. S.B., S.K., and J.G.R. provide variant and gene annotations and their functional interpretation of variants. E.M., J.S., R.L., B.B., A.B., L.H., J.G.R. conceived and are responsible for creating, developing, and deploying analysis platforms and computational methods for analyzing genomic data.

Research Program Management

Marcus B. Jones, Ph.D., Lyndon J. Mitnaul, Ph.D.

Contribution: All authors contributed to the management and coordination of all research activities, planning and execution. All authors contributed to the review process for the final version of the manuscript.
